# Integrated cell type-specific analysis of blood and gut identifies matching eQTL for 140 IBD risk loci and entrectinib as possible repurposing candidate

**DOI:** 10.1101/2024.10.14.24315443

**Authors:** Hélène Perée, Viacheslav A Petrov, Yumie Tokunaga, Alexander Kvasz, Sophie Vieujean, Sarah Regimont, Myriam Mni, Marie Wéry, Samira Azarzar, Sophie Jacques, Nicolas Fouillien, Latifa Karim, Manon Deckers, Emilie Detry, Alice Mayer, Raafat Stephan, Keith Harshman, Yasutaka Mizoro, Catherine Reenaers, Catherine Van Kemseke, Odile Warling, Virginie Labille, Sophie Kropp, Maxime Poncin, Anne Catherine Moreau, Benoit Servais, Jean-Philippe Joly, SYSCID Consortium, BRIDGE Consortium, Wouter Coppieters, Emmanouil Dermitzakis, Edouard Louis, Michel Georges, Haruko Takeda, Souad Rahmouni

## Abstract

Genes whose expression is affected in a consistent manner by GWAS-identified risk variants and the disease process, constitute preferred drug targets. We herein combine integrated cis-eQTL analysis in 27 blood cell populations and 43 intestinal cell types of the ileum, colon and rectum, and information on gene expression in patients, to search for putative drug targets for inflammatory bowel disease (IBD). We detect >95K cis-eQTL that affect >13K e-genes and cluster in >24K regulatory modules (RM). We uncover matching RM for 140 risk loci, implicating >300 e-genes not previously connected to IBD, and find 152 IBD-matching e-genes whose expression is perturbed in the blood or gut of patients. We identify entrectinib, a small molecule inhibiting the NRLP3 inflammasome by binding NEK7, as a promising repurposing candidate for IBD.

## Introduction

The growing prevalence of common complex diseases (CCD) threatens the sustainability of health care systems world-wide. There is a need for more effective prevention and treatment [Busse *et al*., 2010; Schumacher *et al*., 2016; Jakab *et al*., 2018].

Predisposition to most CCD has a considerable inherited component [Polubriaginoff *et al*., 2018]. Accordingly, GWAS with case-control cohorts in the tens to hundreds of thousands of individuals have nearly systematically uncovered tens to hundreds of risk loci that explain up to ∼50% of the heritability [Visscher *et al*., 2017; Abdelloui *et al*., 2023]. Identifying the genes that are perturbed by the risk variants in these loci is considered a key goal, as these constitute preferred drug targets for the pharmaceutical industry [King *et al*., 2019; Burgess *et al*., 2023; Trajonaska *et al*., 2023; Minikel *et al*., 2024].

A common feature of CCD is that the majority of associations are driven by regulatory variants as opposed to coding variants. For example, amongst the more than 200 risk loci now mapped for inflammatory bowel disease (IBD), only 14 encompass ORF-altering variants in the corresponding credible sets (i.e., the set of variants in high linkage disequilibrium (LD) that have a high posterior probability to be causal), thereby rather convincingly pinpointing causative genes and hence putative drug targets [Sazonovs *et al*., 2023]. The prevailing hypothesis is that for the remaining loci, regulatory variants perturb the expression of causative genes in *cis*. The challenges are to identify the causative regulatory variants in the credible sets, and – most importantly - the genes they perturb in *cis*. A common approach to achieve the latter is by means of eQTL analyses performed in disease relevant cell types collected from healthy individuals. Indeed, it is reasonable to assume that: (i) the effect of most regulatory variants will manifest by genotype-specific differences in steady state messenger levels (i.e., eQTL effects), and (ii) the corresponding eQTL effects, caused by mostly common variants, are detectable in healthy individuals as well. If a *cis*-eQTL underpins a GWAS-identified risk locus it will exhibit an association pattern (the vector of association log(1/p) values of all the SNPs in the locus with the gene’s expression levels, henceforth referred to as EAP for <u>e</u>xpression <u>a</u>ssociation <u>p</u>attern) that is very similar to the association pattern of the risk locus (henceforth referred to as DAP for <u>d</u>isease <u>a</u>ssociation <u>p</u>attern), provided that the GWAS and eQTL studies were conducted in cohorts of same ethnicity (and hence same LD structure). A number of colocalization methods have been developed to quantify the resemblance between DAP and EAP [f.i. Giambartolomei *et al*., 2014; Zhu *et al*., 2016; Hormozdiari *et al*., 2016; Momozawa *et al*., 2018].

Several eQTL datasets have been assembled towards that goal, admittedly still heavily biased in favor of Northern European ancestry. The most prominent of those is the GTEx cohort that provides eQTL information for 52 tissues in up to 838 subjects [GTEx Consortium, 2020]. A common observation from such colocalization experiments is that “matching” eQTL are only found for ∼25% of risk loci, raising questions about the molecular underpinnings of the still majority of risk loci [Umans *et al*., 2021]. For example, we previously identified matching eQTL in 63 of 200 analyzed IBD risk loci using a catalog of 23,650 eQTL identified using transcriptome data of six circulating immune cell types and intestinal biopsies at three locations (CEDAR-1 dataset) [Momozawa *et al*., 2018].

A plausible explanation of this still high proportion of “orphan” risk loci, is that the corresponding eQTL remain to be discovered. This could be because the relevant cell populations were underrepresented in the (often heterogeneous) samples studied to date, because the manifestation of the eQTL is context dependent (f.i. after initiation of the disease process), or because the eQTL discovered thus far (with small sample sizes when compared to disease GWAS) are distinct from the ones that drive CCD [Mostafavi *et al*., 2023].

In this work, we followed up on the first hypothesis, i.e., the relevant cell populations are underrepresented in the samples studied to date, in the context of IBD. Towards that goal we established the CEDAR-2 eQTL dataset based on (i) bulk transcriptome data from 27 circulating immune cell populations for 200 healthy individuals, and (ii) single-cell RNA (scRNA) transcriptome data for intestinal biopsies collected at three locations (terminal ileum, transverse colon and rectum) from 60 healthy individuals. Using this dataset, we herein report the identification of matching *cis*-eQTL for 140 of 206 examined IBD risk loci, identify more than 300 novel candidate causal genes, and pinpoint opportunities for drug repurposing.

## Results

### 60,113 cis-eQTL affecting 11,874 eQTL genes in 27 circulating immune cell populations cluster in 22,067 cis-acting regulatory modules

We collected peripheral blood from 251 healthy individuals of both sexes (STable 1). We genotyped all individuals with Illumina’s OmniExpress array interrogating ∼700K SNPs, and augmented genotype data to ∼6.3 million (M) variants by imputation. For each individual, in addition to collecting peripheral blood mononuclear cells (PBMC), we sorted 26 distinct immune cell populations from whole blood by fluorescent activated or magnetic cell sorting (FACS or MACS) (SFig. 1; STable 2). We produced next generation sequencing (NGS) libraries for each of the 27 cellular fractions of each individual (5,292 mRNA SMART-Seq HT libraries in total), and generated an average of ∼12.6M, 2 x 150 bp (3,180 libraries) or ∼13M, 2 x 50 bp (2,112 libraries) paired-end reads per library on Illumina instruments (STable 3). Transcriptome data were used to check the assignment of libraries to individuals and cell types, and errors corrected (SFig. 2A-B). The numbers of genes detected averaged 16,615 per cell type (range: 12,776 – 19,042) (STable 4). Hierarchical clustering of cell types based on average gene expression levels generated a dendrogram largely consistent with hematopoietic ontogeny (SFig. 2C).

We performed *cis*-eQTL analyses using a custom-made pipeline including QTLtools [Delaneau *et al*., 2017], regressing gene expression level on alternate allele dosage and including an average of 24.4 expression principal components (PC) (range: 13 - 36) in the model (STable 4; M&M). We identified 60,113 eQTL (within cell type FDR ≤ 0.05), influencing the expression of 11,874 eQTL genes (e-genes) (STable 5). The number of eQTL detected per cell type (average: 2,226; range: 608 – 3,448) was largely determined by the number of samples available for analysis (*r*^2^=0.67) (SFig. 3A). The number of eQTL detected in PBMC was larger than expected given sample numbers, consistent with PBMC encompassing multiple cell types. The number of eQTL detected in plasmocytes was lower than expected given sample numbers. Controlling for the variable abundance of immunoglobulin transcripts did not increase the number of detected plasmocyte eQTL (SFig. 3B). On average, *cis*-eQTL explained 19.7% (range: 3.2% - 83.6%) of variance in gene expression level (SFig. 3C). Significant secondary *cis* signals (range: 2 - 5) were detected for 6% of *cis*-eQTL, when repeating the eQTL analysis conditional on the previous signals (SFig. 3D).

eQTL affecting the same gene in multiple cell types were merged if sharing a similar association pattern (EAP) as evaluated using theta (|*θ*| ≥ 0.6) [Momozawa *et al*., 2018], yielding 23,831 distinct gene-specific modules (STable 6). Modules were augmented with 5,840 tier-2 eQTL that would nevertheless match (|*θ*| ≥ 0.6) at least one significant eQTL in the module (see M&M). Modules can be characterized by a vector of 0 and 1 that indicates in which cell type(s) the module is active. The 31 most common vectors were dominated by 17,439 modules (i.e., 73.2%) that are active in only one of each of the 27 cell types, and included 117 modules that are – *a contrario* - active in all 27 cell types (Fig. 1A). The overdispersion of the number of active cell types (i.e., excess of modules active either in very few or many cell types) was highly significant (p< 0.001) (see M&M). Sharing of modules by cell types was largely determined by their ontogenic proximity (SFig. 3E).

**Figure 1:**
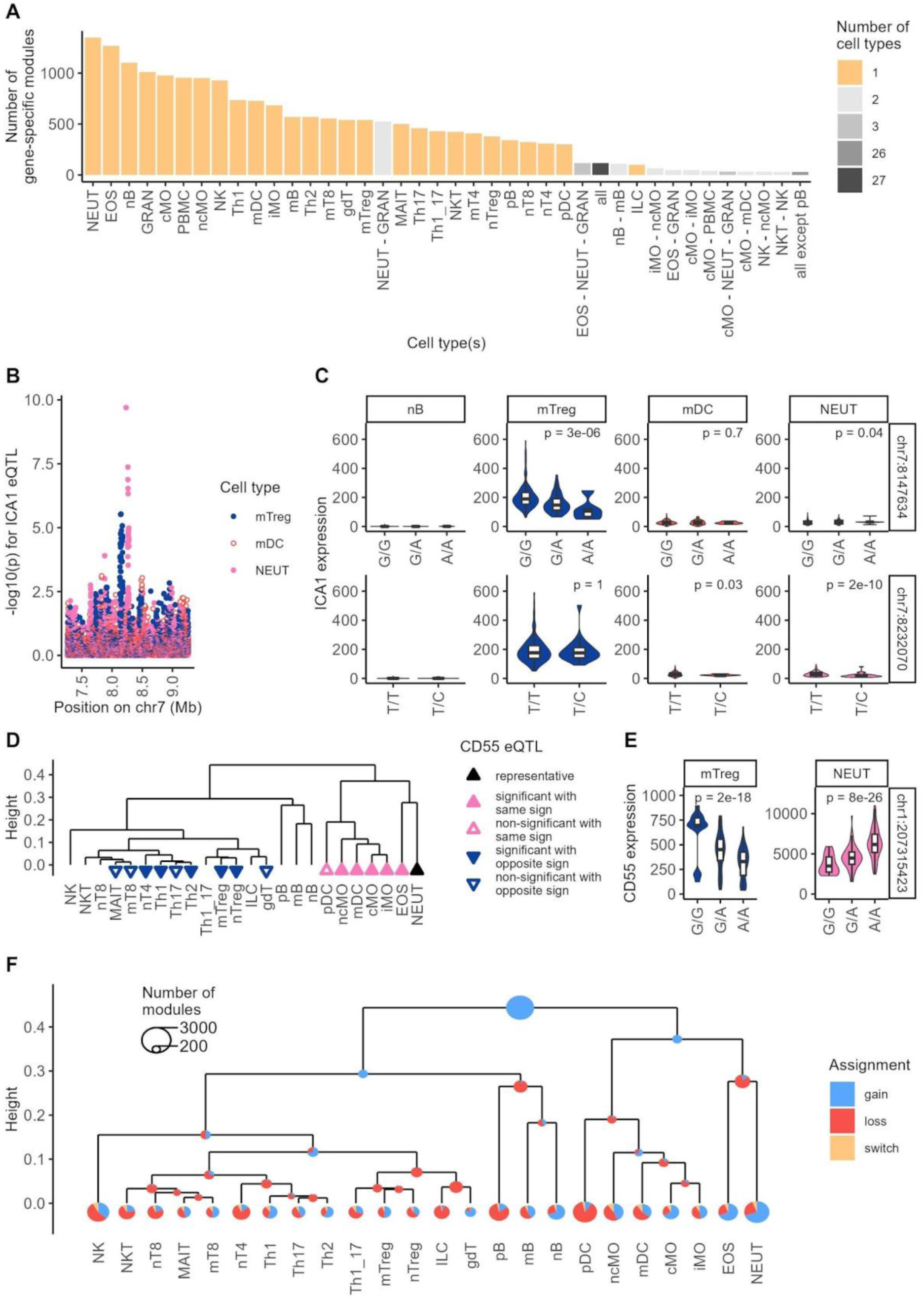
**(A)** Gene-specific modules grouped by cell type combinations in which they are active, and ordered according to the frequency of occurrence (40 most frequent combinations out of 3,572 observed). The most common modules are dominated cell type-specific ones, i.e., only active in one cell type (27 yellow bars). Also, amongst the top 40 combinations, are those corresponding to modules that are active in all (i.e., ubiquitous eQTL; black bar). The remaining combinations that are shown correspond to modules that are active in 2 or 3 closely related cell types (hence still very cell type-specific). Cell type abbreviations are as in STable 2. **(B-C)** Example of a gene (*ICA1*) that is controlled by two distinct modules operating respectively in memory regulatory T cells (mTreg, module #12,606, blue in EAP and violin plots) and neutrophils (NEUT, module #16,344, pink in EAP and violin plots). Neither of these is active in myeloid dendritic cells (mDC, orange EAP and violin plot) despite the fact that the gene is expressed at relatively high levels in this cell type (scenario 2). The gene is expressed at very low levels in naïve B cells (nB) in which consequently neither module is active either (scenario 1). Individuals are sorted by genotype at the lead SNPs for modules #12,605 (upper row) and #16,344 (lower row). **(D-E)** Example of a gene-specific eQTL module (*CD55*, module #678) that is active in 16 of the 24 shown cell types, including all myeloid cell types, and nine of 12 types of T-cells (filled triangles: significant eQTL; empty triangles (tier-2): non-significant eQTL but matching (|*θ*| ≥ 0.6) at least one of the significant eQTL in the module). However, the sign of the eQTL (effect of the alternate allele) is opposite in myeloid cells (pink, upward pointing triangles) and T lymphocytes (blue, downward pointing triangles). The reference sign is determined by the most significant, “representative” eQTL in the module (black triangle). Violin plots show the distribution of *CD55* expression (DESeq2 normalized expressions) in memory regulatory T cells (left, blue) and neutrophils (right, pink) for individuals sorted by genotype at the lead variant for the consensus EAP. **(F)** Gains of gene-specific modules were assigned to the “most recent common ancestor” (MRCA) node of all nodes/leaves in which the module is active (blue segments). Losses of gene-specific modules were assigned to the MRCA node of all descendent (of a node to which a module was assigned) nodes/leaves in which the module was not active (red segments). If the loss of a module coincided with the gain of a module for the same gene, we assumed that a module switch occurred (yellow segments). The diameter of the circles is indicative of the corresponding numbers, *per* legend.

A module can be active in cell type A but not in cell type B because: (i) the gene is expressed in cell type A but not in cell type B, (ii) the gene is expressed in both cell types but the eQTL is only active in cell type A, or (iii) the gene is in distinct modules in cell type A and B (i.e., different EAP in cell types A and B). Expression of the gene was below detection levels in cell type B (i.e., first scenario) in 17.7% of cases. Analysis of the modules indicated that the third scenario (module switch) accounts for 8.5% of cases. We devised an interaction test to evaluate the importance of scenario 2 (see M&M) in the remaining 73.8% of the cases. We estimated the proportion of alternative hypothesis (*π*_1_) following Storey *et al*. [2003] at 97.3%, hence supporting the fact that the eQTL was indeed not active in cell type B in 71.8% of cases despite the fact that the gene was expressed in cell type B (Fig. 1B-C; SFig. 3F). For modules active in more than one cell type, the direction of the effects was the same across cell types in 96.1% of the cases, yet there were 250 cases for which the sign of the effect switched between cell types (Fig. 1D-E; STable 6). The proportion of shared eQTL yet with opposite sign increased with ontogenic distance between considered cell types as expected (*p*_logit_ = 4×10^-33^; STable 7). Modules were assigned to nodes and leaves in the ontogenic dendrogram using the patterns of module sharing across cell types (vectors of 0’s and 1’s) (see M&M), suggesting extensive remodeling of eQTL activity during hematopoiesis, including the presumed common loss of progenitor cell eQTL and gain of new eQTL by differentiated cells, primarily involving different sets of genes (as the proportion of module switches was limited) (Fig. 1F). Genes with *cis*-eQTL assigned towards the root (presumed to be active in progenitor cells, n=2,776) were enriched in GO terms: antigen processing and presentation, amino-acid metabolism, organic acid and small molecule metabolic processes, and cell motility, while genes with *cis*-eQTL assigned towards the leaves (n=9,201) did not show enrichment in any GO terms (STable 8).

We then merged gene-specific modules with matching EAP “across genes”, yielding a total of 22,067 *cis*-acting regulatory modules. Modules were augmented with 7,402 tier-2 eQTL matching at least one significant *cis*-eQTL in the module, recruiting 759 extra genes (M&M; STable 9). We observed 2,470 modules (11.2%) comprising more than one gene. On average such multigenic modules encompassed 2.9 genes (range: 2 - 32) and were active in 9.3 cell types (SFig. 3G). Gene-specific modules active in multiple (but not all) cell types had more chance to join a multigenic module than gene-specific modules active in only one cell type (SFig. 3H). eQTL effects with opposite sign (negative *θ*) were observed for 48.8% of multigenic modules. For the 5,856 modules encompassing more than one EAP (whether from the same or different genes), we combined the constituent association patterns in a consensus EAP representing the module (see M&M). We developed a web browser to visualize the activity of the regulatory modules across the genome and cell types (SFig. 4; https://tools.giga.uliege.be/cedar/publihpq).

### Single-cell RNA Seq analysis of intestinal biopsies reveals 35,010 eQTL affecting 3,007 genes and clustering in 3,337 modules

We collected biopsies in the terminal ileum (IL), transverse colon (TC), and rectum (RE) (locations) of 60 healthy individuals. Epithelium and lamina propria (fractions) were separated, location- and fraction-specific cell suspensions hash-tagged, and subjected to scRNA-Seq using a 10X Chromium platform and Illumina sequencers. We obtained quality-filtered sequence data for a total of 293,801 cells from 57 individuals (5,154 cells per individual on average). The number of reads per cell averaged 48,628, the number of unique molecular identifiers (UMI) per cell 7,422, and the number of genes detected per cell 2,035 (STable 10). Cells were assigned to one of nine sets corresponding to anatomical location (IL, TC, RE) and cell category (epithelial, immune, stromal) (see M&M). K-means clustering of the cells, by set, yielded a total of 276 clusters. Samples were merged using Harmony [Korsunsky *et al*., 2019], and a hierarchical tree (of clusters) constructed using the Euclidean distances between the clusters’ centroids in Harmony space. Leaves (corresponding to clusters in the original nine cell sets) and nodes in the tree were assigned to 13 epithelial, 14 lymphoid, 6 myeloid and 10 stromal cell types (43 cell-types in total) using cell-type specific gene signatures from the literature [Smillie *et al*., 2019; Franzen *et al*., 2019; Hao *et al*., 2021; Burclaff *et al*., 2022; Ishikawa *et al*., 2022; Hickey *et al*., 2023; Kong *et al*., 2023; Krzak *et al*., 2023] (Fig. 2; SFig. 5&6; STable 11). Numbers of cells were relatively evenly distributed across the three anatomical locations (Fig. 2B). Epithelial cells were more abundant than lymphoid, myeloid and stromal fractions combined (Fig. 2C). Strikingly, myeloid, lymphoid and endothelial cells from the three locations overlapped well in UMAP space, while absorptive and secretory epithelial cells as well as fibroblast from distinct locations did not, supporting larger effects of location on the transcriptome for the latter (Fig. 2E). Paneth cells were exclusively observed in ileal samples as expected, while most other cell types were present in the three locations.

**Figure 2:**
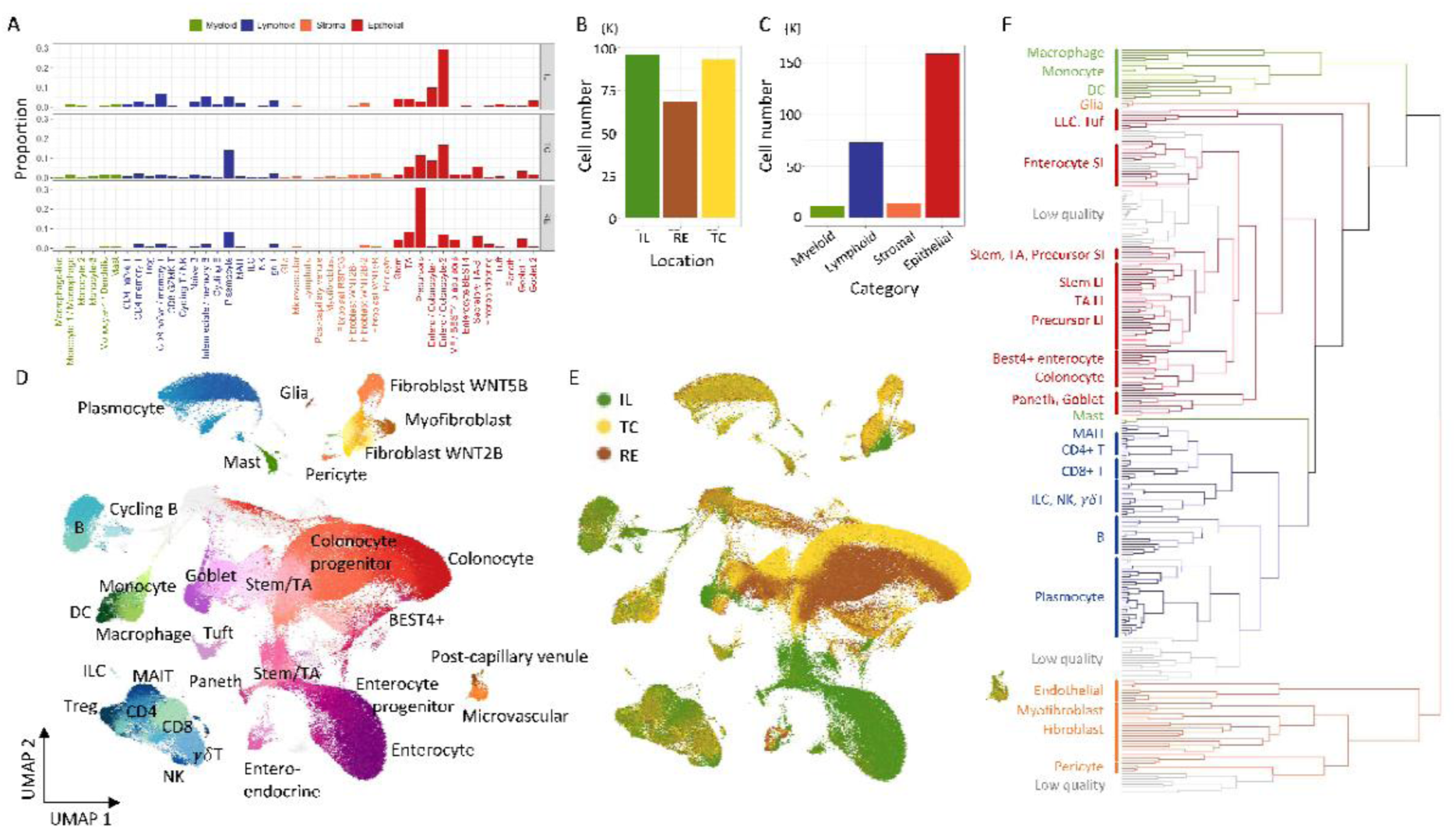
**(A)** Proportion of cells from ileal (IL), colonic (TC), and rectal (RE) biopsies assigned to 6 myeloid cell types (green), 14 lymphoid cell types (blue), 10 stromal cell types (orange) and 13 epithelial cell types (red). **(B)** Number of recovered quality-controlled (QC-ed) cells by anatomical location. **(C)** Number of recovered QC-ed cells by cell category. Colors are as in A. **(D)** UMAP of 293,801 QC-ed cells labeled by cell category (myeloid, lymphoid, stromal, epithelial) and cell-type within category. **(E)** Same UMAP as in D, labeled by anatomical location (colors as in B). **(F)** Hierarchical tree of 276 cell clusters (with 275 nodes). Four cell categories (myeloid, lymphoid, stromal, epithelial) are color-coded as in A and C. The main cell types within categories are labeled.

Epithelial, stromal, myeloid and lymphoid cell types clustered in the tree as expected (except for glia and mast cells) (Fig. 2F). The segregation of ileal, colonic and rectal clusters occurred mostly at terminal branches of the tree, with the exception of absorptive epithelium, in agreement with the overlap in UMAP space (SFig. 6J).

We then conducted eQTL analyses separately by leaf and node across the tree (551 analyses in total). eQTL analyses were performed using the same custom-made pipeline including QTLtools, using a pseudo-bulk approach and leaf/node-specific PEER factors [Stegle *et al*., 2010] to correct for hidden confounders (including variable cell-type proportions) (SFig. 5). We detected a total of 35,010 *cis*-eQTL (within leaf/node FDR ≤ 0.05) affecting 3,007 e-genes (STable 12). The number of eQTL detected per leaf/node was largely determined by the number of cells in the leaf/node with, however, a higher yield per cell for epithelial than for the other cell categories. It plateaued at ∼1,100 presumably limited by sample size (57 individuals) (Fig. 3A). A second independent effect was detected for 259 *cis*-eQTL, and a third for two (STable 12). As for the circulating immune populations, we merged *cis*-eQTL affecting the same gene in 3,345 gene-specific modules when sharing similar EAP (|*θ*| ≥ 0.6), and augmented modules with 22,904 tier-2 eQTL that would nevertheless match at least one significant eQTL in the module (see M&M) (STable 13). We assigned modules to the most recent common ancestor (MRCA) of all active nodes/leaves. Modules mapping near the tree’s root were allocated to pairs of cell categories when possible. Most of the modules mapped towards the root of the tree, as expected as this is where the number of cells per node is largest and hence detection power is highest (Fig. 3B). However, the numbers of leaves/nodes in which a module was active was over-dispersed: modules tended to be either active in fewer leaves/nodes or in more leaves/nodes than expected assuming random assortment (p < 0.001; see M&M) (Fig. 3C), which is reminiscent of the observations in circulating immune cell populations (Fig. 1A). This suggests that numerous cell-type specific eQTL also exist in the gut. Accordingly, we observed 524 modules that were only active in one of the 43 cell-types, of which 429 were also location-specific. The latter were mainly eQTL that were active either in enterocytes or their precursors from the small intestine (SI = IL) or in colonocytes or their precursors from the large intestine (LI = TC + RE), respectively (Fig. 3D). We developed a 3D application to visualize the activity of eQTL on a UMAP, vividly illustrating the location- and cell-type-specific activity of some modules (Fig. 3E). For gene-specific modules active in more than one leaf/node, the sign of the eQTL effect was the same in all leaves/nodes for 99.2% of the modules. For 27 genes, however, the effect differed depending on the leaf/node. In a number of instances, this clearly corresponded to a distinct effect depending on cell type (Fig. 3F; STable 13).

**Figure 3:**
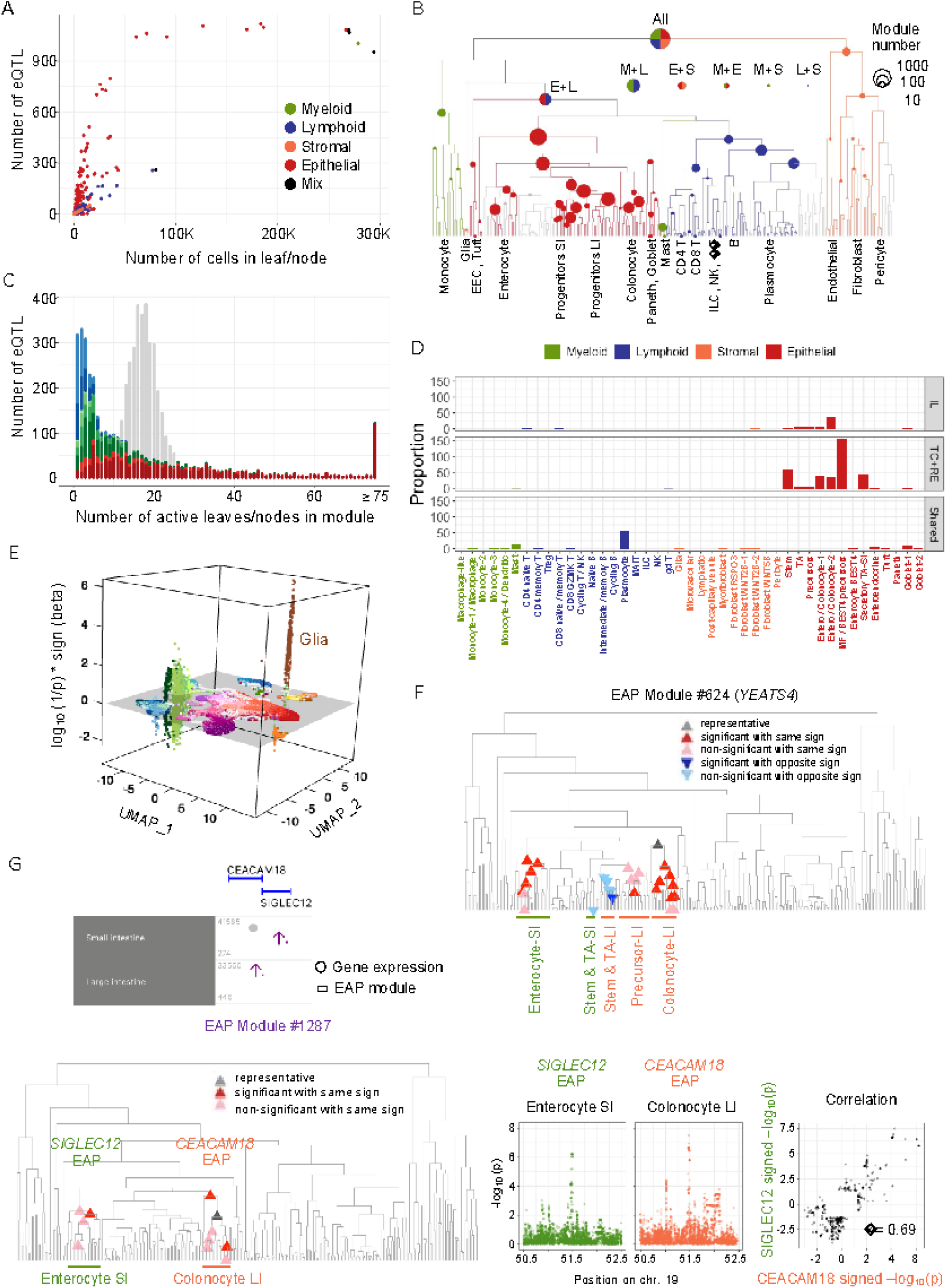
**(A)** Number of eQTL detected (within leaf/node FDR ≤ 0.05) as a function of the number of cells in the corresponding leaf/node. Leaves/nodes are colored by cell category (myeloid: green, lymphoid: blue, stromal: orange, epithelial: red, mix (=multiple categories): black). **(B)** Assignment of 3,345 gene-specific regulatory modules to the MRCA of all active leaves/nodes. Leaves/nodes are colored by cell-type category as in A. The surface of the circles is proportionate to the log of the number of modules assigned to the corresponding leaf/node. Modules initially assigned near the tree’s root were assigned to pairs of cell categories when possible, corresponding to the bisected circles ranked by size. **(C)** X-axis: number of active leaves/nodes in modules. Y-axis: number of observations. The modules are color-coded according to the leaf/node to which they were assigned. Modules assigned to the root are in dark red, modules assigned to a pair of cell-type categories are in red, modules assigned to one cell-type category are in green (shades of green (from dark to light) correspond to increasing levels of cell-type specificity in the category), modules assigned to a specific anatomical location are shown in blue. The grey distribution was obtained by randomly permuting activity status across modules, yet keeping the number of significant eQTL per leaf/node as for the real data (see M&M). **(D)** Number of modules that are specific for one of the 43 most granular cell-types, whether location specific (IL versus TC+RE) or shared across locations. Cell categories are labeled as before. **(E)** Example of a highly cell type-specific eQTL effect (*GFRA2* in glia (brown)). The x- and y-axes correspond to the UMAP 1 & 2 axes, while the z-axis measures the strength of the association (log(1/p) multiplied by the sign of *β*. **(F)** Example of a gene-specific regulatory module (gene: *YEATS4*) for which the sign of the eQTL effects differs between cell types. The module is primarily active in absorptive intestinal epithelium in small (SI = IL, green) and large (LI = TC + RE, orange) intestine. The sign of the eQTL effect switches upon transition from stem/TA cells to precursor enterocytes in both SI and LI. **(G)** Example of two adjacent genes that are controlled by the same *cis*-acting regulatory module yet in different cell types: *SIGLEC12* in enterocytes of the small intestine (SI: green) and *CEACAM18* in enterocytes of the large intestine (LI: orange). The positions on the tree where the RM regulates the corresponding genes are shown as triangles (left panel). The corresponding EAPs and theta-plot are shown (right panels).

We then merged intestinal gene-specific modules characterized by similar EAP as measured by *θ*. These across-gene modules were augmented with 24,333 tier-2 eQTLs (see M&M) recruiting 715 extra genes. This yielded 666 modules encompassing EAP from more than one gene (21.6%), and 2,415 modules that remained monogenic. The number of genes in multigenic modules averaged 2.8, ranging from 2 to 16 (STable 14). The proportion of multigenic modules encompassing eQTL effects with opposite sign was 53%. We observed a number of instances where distinct genes were controlled by the same variants (i.e., were assigned to the same regulatory module) but in distinct cell types (Fig. 3G). All intestinal eQTL/module information in their genomic context is browsable using the same website as for the blood module information (SFig. 7; https://tools.giga.uliege.be/cedar/publihpq).

### Merging blood and intestinal cis-eQTL modules reveals eQTLs that are specific for gut-resident immune cells

We then merged EAP in regulatory modules using the same approach as before, but this time for the two datasets combined, i.e., blood and biopsies (STable 15 and 16). This yielded 24,745 across-gene modules, of which 8,472 were active in more than one cell type. Of the latter, 2,172 were found to be active in both blood and biopsies (Fig. 4A). Amongst those, there was a significant excess of modules that were active in multiple cell types in both blood and biopsies (p < 10^-5^), as well as modules that were lymphoid-specific in both blood and biopsies (p = 7 x 10^-4^). Modules that were active in multiple cell types in the blood had more chance to be detected in biopsies than cell-type-specific blood modules (p < 10^-5^), while modules that were enterocyte-specific in biopsies had less chance to be detected in blood (p < 10^-5^), all as expected (Fig. 4B; Stable 17).

**Figure 4:**
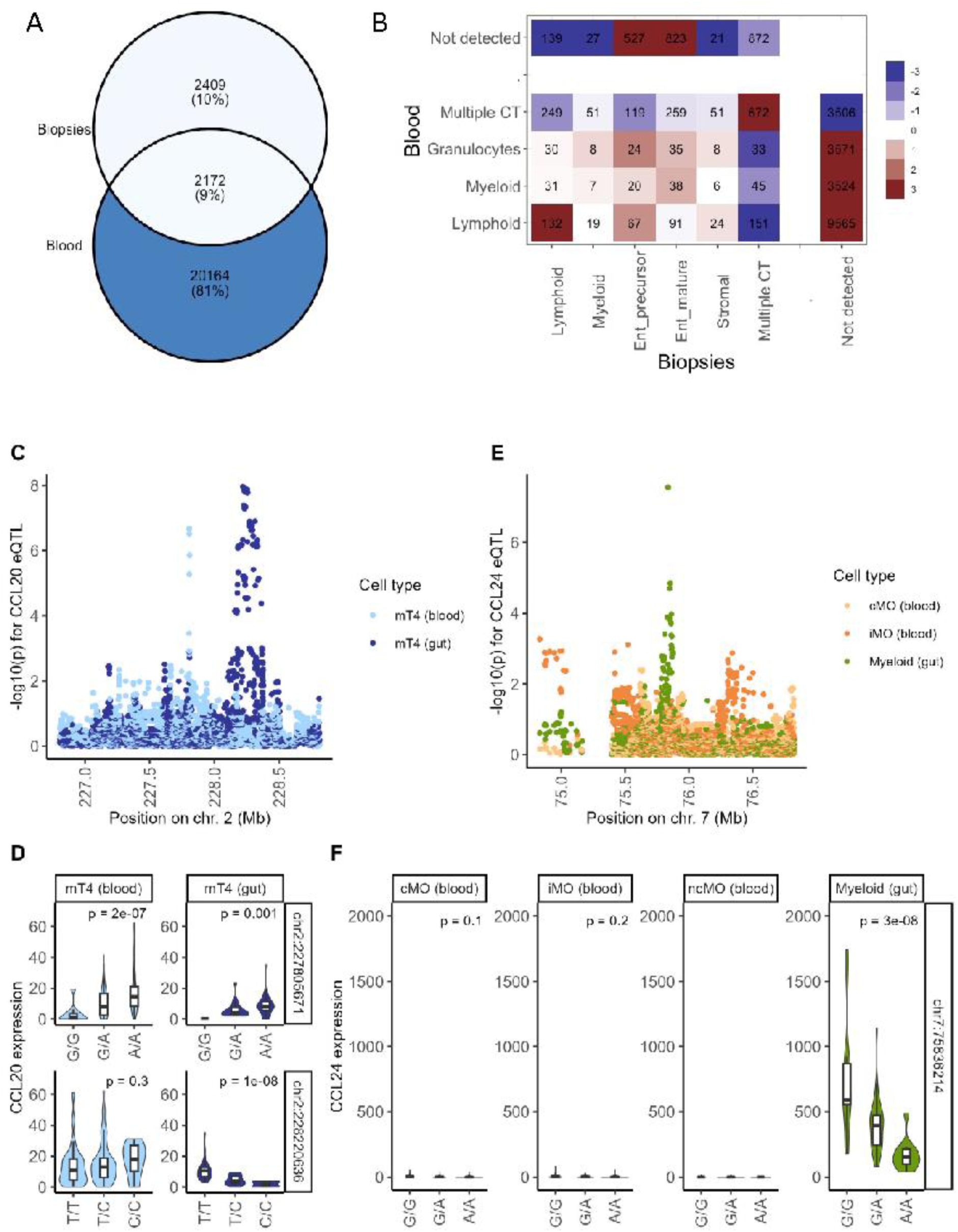
Merging regulatory modules across blood and gut samples. **(A)** When merging all blood (n=60,113) and gut (n=35,010) eQTL jointly in regulatory modules, we obtained a total of 24,745 across-gene modules, including 8,472 that are active in several cell types of which 2,172 (9%) encompassed eQTL from blood and gut. **(B)** Modules were assigned to cell types, separately for blood and gut, as described before. Blood cell types were grouped in lymphoid, monocytes/dendritic cells (myeloid), granulocytes or multiple of these cell types (i.e., active in several of the other categories). Intestinal cell types were grouped in lymphoid, myeloid, enterocyte precursors, mature enterocytes, stromal or multiple of these cell types. “Not detected” indicates that the module is not active in the corresponding sample type (blood or gut). The numbers in the tiles correspond to the number of observations for the corresponding combinations. The colors correspond to -log(*p*) of an empirical test of independence (i.e., to what extent do the observed numbers deviate from expectation assuming that the proportions of the different categories in blood and gut are independent). Red: excess. Blue: depletion. **(C)** Example of a gene (*CCL20*, C-C Motif Chemokine Ligand 20) that is expressed in circulating as well as gut-resident memory CD4 T lymphocytes (mT4). However, the gene is subject to two clearly distinct eQTL in these two compartments (light blue: blood mT4 EAP, dark blue: gut mT4 EAP). Of note the EAP observed in circulating cells matches a DAP for UC (Table 21). The corresponding EAP is also detectable in one intestinal mT4 leaf, which could very well correspond to blood present in the biopsy. **(D)** Violin plot showing the expression levels of *CCL20* in blood mT4 (left panels in light blue) and gut-resident mT4 (right panels in dark blue) for individuals sorted by genotype for the top SNP of the light blue blood EAP (upper panels) and the top SNP of the dark blue gut EAP (lower panels). **(E)** Example of a gene (*CCL24*) that is strongly expressed in gut-resident myeloid cells (including monocytes and dendritic cells) but nearly undetectable in the equivalent circulating cells (shown for the three types of monocytes: conventional (cMO), intermediate (iMO), and non-conventional (ncMO). *CCL24* is subject to an eQTL that is detectable in gut-resident myeloid cells (green EAP) but not detectable in circulating monocytes (as the gene is virtually not expressed) (yellowish EAP). **(F)** Violin plots showing the expression of the *CCL24* gene in circulating monocytes (three panels on the left), and in gut-resident myeloid cells (panel on the right) for individuals sorted by genotype for the top SNP of the gut EAP in (E).

We mined the corresponding gene-specific catalog for modules that were labeled lymphoid- or myeloid-specific in biopsies (two cell type categories that are also present in blood), but were reported as “Not detected” in blood. We reasoned that such a list might be enriched in *cis*-eQTL that would not be active in circulating immune cell population(s), but would reveal themselves in the same immune cell population(s) once becoming gut-resident. Hundred-thirty-one modules matched this pattern, of which 57 were dropped after visual inspection of the corresponding EAP using the CEDAR2 website. Of the remaining candidates: (i) 8 appeared to be mastocyte-specific eQTL, a myeloid cell type not present in blood, (ii) 39 corresponded to genes that were considered to be expressed at too low level in the corresponding circulating blood population to warrant eQTL analysis (and hence likely differentially expressed between cognate circulating and resident cells, and reminiscent of scenario 1 above), and (iii) 26 appeared to be subject to gut-specific eQTL not active in the cognate circulating population despite the genes being detectable (hence reminiscent of scenario 2 above) (STable 18). Thus, it appears that there not only exist many cell-type-specific eQTL, but that – for a given cell type – eQTL can be context specific, f.i. manifest in some anatomical compartments (resident) but not in others (circulating). *CCL20* and *CCL24* constitute two interesting such examples (Fig. 4C-F).

### Identifying new cis-eQTL driving inherited predisposition to IBD

We then mined our database of blood and intestinal *cis*-eQTL for EAP-matching disease-association patterns (DAP) for inflammatory bowel disease (IBD), with the aim to identify novel candidate causative genes and hence putative drug targets. We defined the boundaries of 206 risk loci reported by Lange *et al*. [2017], encompassing 173 IBD loci (considering CD and UC patients jointly in GWAS), 157 CD loci and 125 UC loci, by visual examination of the local association patterns obtained with (Europeans-only) ∼25K cases and ∼35K controls from the International IBD Genetics Consortium (IIBDGC) (STable 19). Thirty-two risk loci that encompass composite peaks were further subdivided in sub-risk loci, to be confronted separately to *cis*-eQTL in addition to the cognate complete risk locus. It is indeed conceivable that larger, multi-peak DAP reflect the compound effects of multiple *cis*-eQTL either of the same or different genes in the same or different cell types. Splitting the corresponding risk loci may reveal distinct matching eQTL. Colocalization analyses were conducted using the *θ* metric, as described in Momozawa *et al*. [2018]. To define suitable thresholds for significance, we performed parallel analyses using permuted genotype data, separately for blood and biopsies (genome-wide *cis*-eQTL analysis in all cell types, leaves and nodes). Confronting results obtained with the real versus permuted data allowed us to compute an FDR for each DAP-EAP pair as a function of the value of |*θ*| (≥ 0.6), its *p*-value, as well as the *p*-value (adjusted for *cis*-window) for the eQTL (see M&M). This yielded matching DAP-EAP with FDR ≤ 0.05 for 379 genes in 119 risk loci (tier-1), or 556 genes in 140 risk loci when considering matching DAP-EAP with FDR ≤ 0.10 (tier-1+2) (Fig. 5A; Stable 20-22). No credible extra DAP-EAP matches were detected when using the modules’ consensus EAP. Of note, and to the best of our knowledge, no eQTL-based connection with IBD has been previously reported for 366 of the 556 genes (STable 22). Matching EAP were detected in both blood and gut for 77 (55%) risk loci, only in blood for 33 (24%), and only in the gut for 30 (21%). Thus, the scRNA-Seq data yielded a comparable number of DAP-EAP matches despite its limited sample size. For risk loci with matching DAP-EAP in both blood and gut, the e-genes involved were generally different. The number of e-genes with matching EAP averaged 4 per risk locus, ranging from 1 to 34 (i.e., for the 140 risk loci with at least one match). Local gene density explained ∼25% of the differences in number of matching e-genes per risk locus. There was a strong correlation between the number of matching e-genes in blood and gut for a given risk locus, despite the fact that the genes involved mostly differed (SFig. 8C and 8D). In circulating immune cells, regulatory modules active in all cell types contributed disproportionately to DAP-EAP matches, reminiscent of a previous report [Momozawa *et al*., 2018]. Concomitantly, in biopsies, modules assigned to the root of the tree accounted for the largest proportion of DAP-EAP matches (SFig. 8H and 8I). We note the modest enrichment (1.3-fold) of modules active only in circulating natural killer (NK) cells. Only 12.5% of the 472 (= 556 – 84 IBD-only genes) DAP matching e-genes were shared between the two diseases. This proportion only increased to 31.8% when restricting the analysis to the 25 (of 140) risk loci associated with both CD and UC. This possibly underscores the distinct molecular determinism of the two pathologies (SFig. 8 F and 8G).

**Figure 5:**
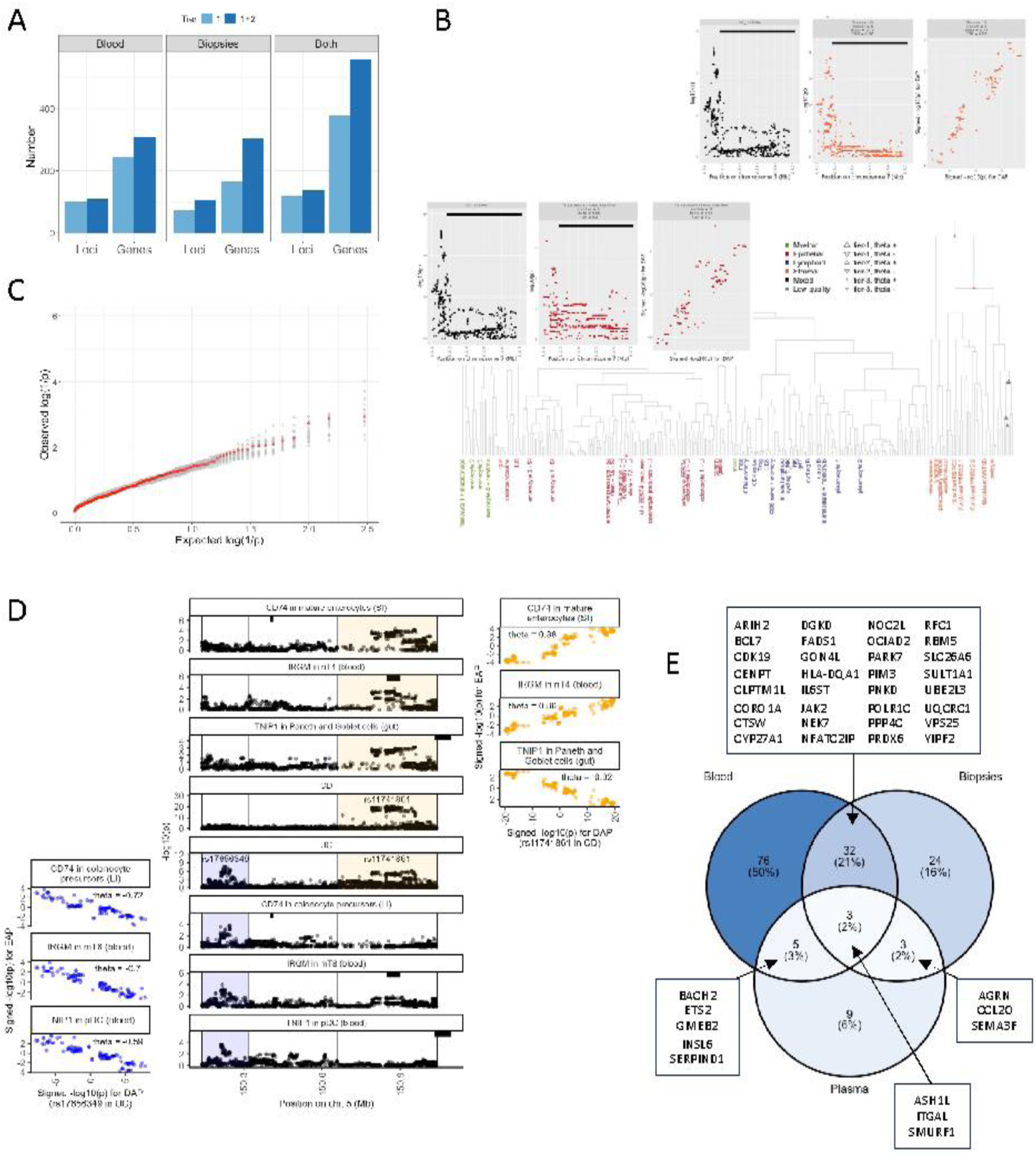
**(A)** Numbers of IBD risk loci and genes identified with matching DAP-EAP in the 27 circulating immune cell populations (blood), in the 43 intestinal cell populations (biopsies), and when combining both datasets (Both). Tier 1 corresponds to matching DAP-EAP with |*θ*| ≥ 0.6 and FDR ≤ 0.05. Tier 2 corresponds to matching DAP-EAP with |*θ*| ≥ 0.6 and 0.05 < FDR ≤ 0.10. **(B)** EAP of the cystic fibrosis *CFTR* gene matching the DAP of a UC risk locus on chromosome 7 (rs38904) in secretory TA precursor cells (red) and intestinal stromal cells (orange). The nodes/leaves with DAP-matching EAP are marked by triangles (large: FDR < 0.10, small: FDR > 0.10). Insets: (left) DAP for UC, (middle) EAP for *CFTR*, (right) *θ* plot, (red) secretory TA precursor cells, (orange) stromal cells. The boundaries of the CFTR gene are marked by the thick horizontal black line. The positive sign of *θ* indicates that downregulation of the *CFTR* gene in these cell types may be protective, which is corroborating the recent finding that *CFTR* loss-of-function mutations protect against CD [Yu *et al*., 2024]. **(C)** QQ plot generated with the burden test p-values obtained for 298 of our 556 DAP matching e-genes by Sazonovs *et al*. [2022] (red dots). Grey dots correspond to QQ plots obtained with randomly sampled sets of 298 genes from the list of genes with data in Sazonovs *et al*. [2022]. **(D)** Central panel: DAP for CD and UC in two adjacent risk loci on chromosome 5 (rs17656349 shaded in blue, and rs11741861 shaded in yellow), as well as (below) EAP matching the rs17656349 UC DAP for *CD74* (colonocyte precursors in large intestine), *IRGM* (circulating memory CD8), and *TNIP1* (circulating plasmacytoid DC), and (above) EAP matching the rs11741861 CD/UC DAP for *CD74* (mature enterocytes in small intestine), *IRGM* (circulating naïve CD4), and *TNIP1* (paneth and goblet cells). The genomic position of the corresponding genes are marked by black horizontal bars. The corresponding theta plots are shown on the left (rs17656349 in blue) and right (rs11741861 in yellow), respectively. **(E)** DAP matching e-genes whose expression levels are affected by the IBD disease process in a direction that is consistent with the effect of risk variants (i.e., both risk variant and disease increase expression, or both risk variant and disease decrease expression) in one or more of 27 circulating immune populations (Blood), in intestinal biopsies (Biopsies, Kong *et al*., 2023), or in plasma (Plasma, Eldjarn *et al*., 2023).

Reactome analysis conducted with the complete gene list highlighted four pathways: interferon gamma signaling (found entities: *CIITA*, *IRF5*, *IRF6*, *PTPN2* and *MHC class I* and *II* genes), interleukin-6 signaling (*IL6ST*, *IL6R* and *JAK2*), chemokines and their receptors (*CXCR1, CXCR2, CCR2, CCR6, CXCL2, CXCL5, CCL20*), and RUNX regulated immune response and cell migration (*ITGA4, ITGAL*) (Stable 23). We scanned the literature, for the 481 protein coding genes out of the list of 556, for functional evidence (other than association or differential expression-based) regarding epithelial barrier function, innate or adaptive immunity that would be considered as support for causality if generated as follow-up of the GWAS and eQTL colocalization. We found such support for 216 of the 556 genes (Table 1; Stable 24 and 25). This included four genes associated with monogenic forms of human inflammatory bowel disease (*CARMIL2*, *PMVK*, *TMEM50B* and *TNIP1*), and 69 genes that upon perturbation affect susceptibility to colitis in a rodent model (Stable 25). The number of genes with incriminating functional evidence averaged 1.46 per risk locus (across the 140 risk loci), with a maximum of 15 for the rs3197999 locus on chromosome 3 (48.48-50.23 Mb). There were multiple risk loci with more than one strongly supported candidate gene. For example, the chr1:rs3180018 locus harbors the DAP-matching *IL6R* e-gene, member of the inflammatory cytokine pathway highlighted by the Reactome analysis, but also the DAP-matching *PMVK* gene causing IBD-like manifestations in a compound heterozygote [Yildiz *et al*., 2023]. Along similar lines, the chr5:rs17656349 locus harbors the DAP-matching *IRGM* gene, regulating autophagy and response to various pathogen-associated molecular patterns (PAMPs), and also *TNIP1* coding for the “*TNFAIP3* interacting protein 1”, knowing that *de novo* mutations in *TNFAIP3* are associated with juvenile IBD [Zou *et al*., 2020; Tanigushi *et al*., 2021]. Similarly, the chr16:rs28449958 locus encompasses *CARMIL2* causing very early onset IBD [Roncagalli *et al*., 2016; Magg *et al*., 2019; Bosa *et al*., 2021], and also *SMPD3*, known to regulate TNF-*α* response in macrophages and B cells, and to influence the severity of DSS-induced colitis when modulated in mice [Liu *et al*., 2017; Al-Rashed *et al*., 2020; Li *et al*., 2024]. There was no correlation between the number of genes with supporting functional evidence in a risk locus and its odds ratio on disease.

**Table 1:**
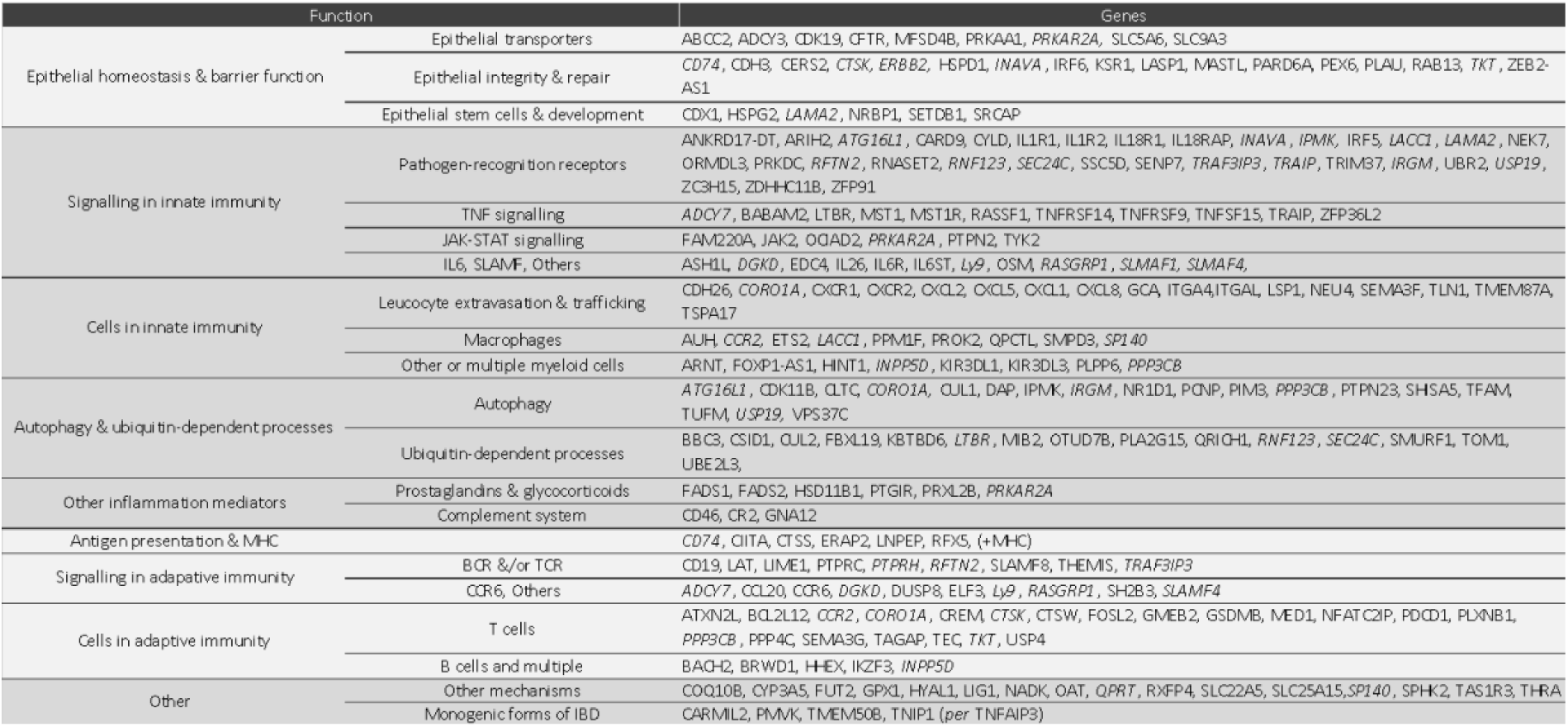
List of 211 e-genes with DAP-matching EAP and published evidence (documented in STable 25 and Suppl. Material) for an effect on one or more functions in one or more cell types participating in epithelial barrier, innate and adaptive immunity, or other IBD-relevant mechanisms. Genes affecting multiple categories of functions are italicized.

Particularly noteworthy is the observation that variants increasing the expression of the cystic fibrosis-causing *CFTR* gene in stromal and/or epithelial precursor cells increase the risk for UC while possibly decreasing the risk for CD (Fig. 5B). This may be related to recent reports that loss-of-function coding variants in *CFTR* protect against CD [Yu *et al*., 2024]. In addition, we found that variants affecting the expression of *PRKAA1* (shown to phosphorylate and modulate *CFTR* activity [Hallows *et al*., 2003]), *CDK19* (shown to control the *CFTR* pathway in the intestinal epithelium of mice [Prieto *et al*., 2022]), *ADCY3* and *PRKAR2A* (both linked to *β*2 adrenergic-dependent *CFTR* expression [Belinky *et al*., 2015]) also affect IBD susceptibility, although these effects were often detected in cells other than the intestinal epithelium. Of note, we observed that variants that decrease *SLC9A3* expression in enterocytes increase UC risk. Loss-of-function mutations in the *SLC9A3* sodium-proton antiporter cause congenital diarrhea 3 and 8 [Dimitrov *et al*., 2019].

Recently, the whole exome of ∼30,000 IBD patients was sequenced and rare-variant burden tests (MAF < 0.001) conducted for 11,978 genes [Sazonovs *et al*., 2023]. The distribution of burden test *p*-values for 298 of our DAP-matching e-genes with sequence information did not depart from expectations under the null (Fig. 5C). Nevertheless, our list of 556 includes *TAGAP*, one of nine genes with a single associated rare coding variant (as opposed to multiple variants considered jointly in a burden test) identified in this study: a rare missense variant (E147K) protecting against CD (OR: 0.786). *TAGAP* has an EAP in enterocyte progenitors that matches the DAP of a CD risk locus on chromosome 6 (rs212388) with positive *θ* (0.79, FDR = 0.04), hence compatible with the protective effect of the missense variant. Of note, borderline DAP-EAP hits were observed for two other genes in Sazanovs’ list of nine, namely *CCR7* (memory CD4 in gut) and *RELA* (plasmocytes in gut). However, for those two genes the positive sign of *θ* did, *a priori*, not match the increased risk associated with the reported missense variants. One gene, *ATG4C*, was further incriminated in this study based on a mutational burden attributed to three missense variants (suggestive signal). *ATG4C* was not part of our list, but *ATG16L2*, a paralogue of the *ATG16L1* autophagy gene not previously incriminated in IBD, yielded a borderline signal with a convincing *θ* of 0.77 despite a modest eQTL signal (*p*_*window adj*_ = 0.27).

We noticed 10 instances where distinct, cell type-specific EAP from the same gene match DAP from IBD risk loci that were considered different albeit adjacent (*QPRT, EIF2B4, TNIP1, PRXL2B, SLC35E2B, IRGM, CDK11B, CD74, SLC25A15, ZNF589*). For example, an EAP for *IRGM* in memory CD8 cells matches the UC DAP in risk locus chr5:rs17656349, while a distinct *IRGM* EAP in naïve CD4 cells matches the CD DAP in the adjacent chr5:rs11741861 locus. In the same risk locus, a *CD74* EAP in colonocyte precursors of the large intestine matches the UC DAP in the chr5:rs17656349 risk locus, while a distinct *CD74* EAP in mature enterocytes of the small intestine matches the CD DAP in the adjacent chr5:rs11741861 locus (Fig. 5D). Similar observations were made for multiple sub-risk loci as defined above (STable 22). Thus, complex “composite” disease association patterns may reflect the effect of distinct risk variants that perturb the expression of the same gene in different cell types, with possibly distinct effects on disease. Also, the DAP for CD and UC, even when overlapping and considered as the same risk locus, may differ and match distinct EAP. For example, the CD DAP in risk locus chr6:rs1819333 matches an EAP for *RNASET2* that is detected in nearly all circulating immune populations, while the distinct UC DAP in the same risk locus matches a *CCR6* EAP active only in NKT cells.

We also observed at least 10 e-genes with DAP-matching EAPs in multiple cell types, for which the sign of *θ* differed between cell types either for the same (CD: *FADS1, IL18R1*; UC: *TOM1*; IBD: *PRXL2B, UBE2L3*) or for different forms of IBD (*CD74, IL1R2, IRGM, PRXL2B, CD244 (=SLAMF4), SLC25A15*) (f.i. Fig. 4D). This calls for caution when defining the desired effect of a drug (activator or inhibitor) based on the sign of *θ*.

The CEDAR2 web-site [https://tools.giga.uliege.be/cedar/publihpq] allows for convenient visual inspection of the matching DAP-EAP patterns underpinning our analyses as well as downloading of the data.

### Entrectinib, a small molecule NEK7 inhibitor, is a repurposing candidate for IBD

It is not obvious that pharmacological targeting of causative genes (that upon perturbation by common regulatory variants increase the chance to develop the disease), will succeed in reversing the disease process once initiated, i.e., be curative. Additional prioritization of candidate genes whose expression is also perturbed by the disease process itself may be useful. To that end we collected blood from 55 active CD patients, performed RNA-Seq on the same 27 fractionated circulating immune cell populations, and performed differential expression analysis between the two cohorts (cases vs controls) by cell type (STable 26). We additionally consulted lists of genes that were shown, from scRNA-Seq data, to be differentially expressed between intestinal biopsies of IBD cases and controls [Kong *et al*., 2023], as well as lists of proteins that were differentially abundant in plasma of IBD cases and controls [Eldjarn *et al*., 2023]. The expression of 109 of our 556 e-genes differed in cases in a manner that was consistent with the sign of *θ* (i.e., both risk variant and disease increase expression, or both risk variant and disease decrease expression) in one of the three data sets (circulating immune populations, biopsies or plasma), of 40 e-genes in two of the three data sets, and of three e-genes in the three data sets (circulating immune populations and biopsies and plasma) (Fig. 5E).

Hundred eighty-one drugs targeting 180 e-genes are or have been used/tested to treat IBD [Mountjoy *et al*., 2021; Vieujean *et al*., 2024] (STable 27). Eleven of these overlapped with our list of 556 e-genes: *CXCR2* (elubrixin: abandoned for lack of efficacy)*, IL6R* (TJ301: phase 2 ongoing)*, IL6ST* (olamkicept: phase 2 completed with positive results), *IMPDH2* (mycophenolate: abandoned for undocumented reasons)*, ITGA4* (multiple in phase 2 and two approved including vedolizumab)*, ITGAL* (efalizumab: abandoned for safety issues including multifocal leukoencephalopathy), *JAK2* (multiple with positive results after phase 2 for peficitinib)*, NDUFAF1* (metformin: phase 2 ongoing), *PDCD1* (rosnilimab: phase 2 ongoing)*, TEC* (ritlecitinib: phase 2 completed with positive results for CD and UC) and *TNFSF15* (several: phase 2 ongoing) (STable 22). For seven of these (*IL6R, IL6ST, ITGA4*, *ITGAL, JAK2*, *IMPDH2*, *PDCD1*), the effect (presumed activator vs inhibitor) of at least some of the drugs was in agreement with the sign of *θ*. For six (*CXCR2, IL6ST, IL6R, ITGAL, JAK2, NDUFAF1*), the effect of disease on expression/abundance level was compatible with the effect of the drug (STable 22). We further identified another 40 genes in our list of candidates, targeted by known drugs (in phase 1 or higher) that – to the best of our knowledge – have not been tested in the context of IBD (STable 22). For 16 of these the activity of at least one drug (likely inhibitor or activator) was consistent with the sign of *θ*. The corresponding drugs were in phase 1 for two (*<u>PIM3</u>, <u>RPS6KB1</u>*), phase 2 for six (*<u>ATP2A1</u>, CDK11B, <u>ERAP2</u>, HLA-DRB1, KIR2DL1, PPP5C*), phase 3 for one (*INPP5D*), and approved for at least one disease other than IBD for seven (*<u>CFTR</u>, CYP3A5, IL18R1, IL18RAP, LAMA2, <u>NEK7</u>* and *PTGIR*). For 6 of the 16 genes (underlined), expression was predominantly affected by the disease process in a manner consistent with *θ*, at least in one of the three datasets. Detailed examination of the drugs that were in phase 3 or higher (Supplemental Material 1), identifies entrectinib (ENB) as a possible repurposing candidate for IBD. Entrectinib (ENB) is a potent tyrosine multikinase small-molecule inhibitor that targets the NTRK, ROS1 and ALK oncogenes, approved by the FDA for the treatment of various tumors [Liu et al., 2018]. It was recently shown to bind to arginine 121 of NEK7, thereby inhibiting its interaction with NLRP3 [Jin et al., 2023]. Downregulation of *NEK7* by intraperitoneal injection of lentiviruses expressing anti-*NEK7* shRNAs was shown to attenuate DSS-induced colitis [Chen et al., 2019]. In mouse models, ENB effectively reduced symptoms of NLRP3 inflammasome-related diseases (other than colitis) [Jin et al., 2023]. ENB has a high safety profile and is well tolerated by almost all patients without cumulative toxicity [Jiang et al., 2022]. Genetic variants that increase *NEK7* expression in naïve B cells increase the risk for IBD (*θ*=0.94; FDR=0.02), while *NEK7* expression is increased in a majority of circulating immune cells of active CD patients, and in non-inflamed colonic epithelium of CD patients when compared to controls [Kong et al., 2023]. On the downside, there is some evidence, albeit non-significant, that the same genetic variants decrease NEK7 expression in some gut-resident immune cells (https://tools.giga.uliege.be/cedar/publihpq), while *NEK7* expression is up-regulated in some circulating immune cells of active CD patients (f.i. eosinophils, Stable 22).

## Discussion

### Regulatory modules: sensible (e)QTL analysis framework?

It is generally admitted that most CCD risk variants act by perturbing the expression of causative genes in one or more disease-relevant cell types, and that it should be possible to pick-up many of these regulatory effects as *cis*-eQTL in these cell types. *Cis*-eQTL effects are pervasive and it is therefore not sufficient to identify a *cis*-eQTL overlapping a risk locus to assume that it affects risk. By definition, causal *cis*-eQTL are determined by the same variant(s) that affect disease risk. As a consequence, the pattern of association between regional variants and gene expression (EAP) should be the same as that for the disease (DAP). This is certainly the case if disease and gene expression are measured in the same individuals. It is also the case if disease and gene expression are measured in distinct cohorts, provided that they share local LD structure. The DAP-EAP similarity applies to the causal variant(s) *per se*, but also to passenger variants whose association with disease and gene expression is indirect, reflecting their LD with the causative variant(s). The expression of a given gene may be controlled by distinct sets of regulatory variants in distinct cell types. This will yield distinct EAP for the same gene, even if the respective sets of regulatory variants partially overlap. The DAP will only match the gene’s EAP in the disease-relevant cell type. It is conceivable that the expression of a causal gene in more than one cell type (with distinct EAP) influences disease risk. If the significant variants for the corresponding EAP are sufficiently distant, multiple DAP-matching EAP may be found for the same gene in different cell types. We have observed several such instances in this work, for adjacent sub-risk loci or even adjacent risk loci considered separate thus far (f.i. Fig. 5D & STable 22). We have also observed instances where distinct EAP (corresponding to different cell types) for the same gene match the DAP for different diseases (in this case CD and UC). If the significant variants of the distinct EAP overlap, the DAP may not match either EAP. More advanced approaches would be needed to dissect such cases, converging towards fine-mapping of multiple independent variant effects. This, however, requires larger sample sizes than what is presently available for multi-tissular eQTL studies.

Causal *cis*-eQTL are the first links in the chain connecting risk variants with disease, the final outcome. The same regulatory module-based approach can in theory be used to uncover the intermediate molecular links between risk variants and disease, provided that the abundance or state of the corresponding molecular species can be quantified. The corresponding *trans-* QTL (whether expression QTL or any other quantifiable molecular phenotype) should be characterized by DAP-matching EAP. CCD are highly polygenic, influenced by hundreds of risk loci or more. It is likely that at least some pathways linking variants with disease converge prior to disease outcome. Thus, some intermediate molecular phenotypes will have matching EAP with multiple DAP (distinct risk loci). This should allow reconstruction of the topology of the pathways linking the multiple risk variants with the disease.

### Cell-type specificity of regulatory modules operating in immune cell populations

One striking observation of this work, is that 73.2% of blood regulatory modules were found to be active in only one of the 27 studied immune cell populations. Most of the time (100-8.5=91.5%) the corresponding genes did not appear to be under marked genetic *cis*-control in the 26 other cell types (i.e., no module switch). Hence, a large proportion of eQTL appear to be very cell-type specific at the chosen level of granularity, at least in blood. One could argue that this is a power issue: the eQTLs may have existed in some other cell type, but remained under the radar of statistical significance because of insufficient sample size. We therefore expanded our search for matching EAP to non-significant eQTL, i.e., we allowed for tier-2 eQTL to enter into the modules if their EAP matched significant ones with |*θ*| ≥ 0.6 and a combination of *p*-values of match and eQTL ensuring an FDR ≤ 0.05 (see Results and M&M). This had virtually no effect on the proportion of modules that were active in only one cell type (from 76% to 73.2%). We therefore think that the observed eQTL cell-type specificity is genuine. These findings are reminiscent of those reported in circulating immune cells by Schmiedel *et al*. [2018] and Ota *et al*., [2021], and across multiple tissues by the GTEx consortium [2020].

The eQTL specificity may even be more pronounced for at least some immune cell populations, being in addition context-dependent. Indeed, the availability of scRNA-Seq data for intestinal biopsies allowed us to compare eQTL activity for the same cell type yet circulating in blood on the one hand, and resident in the intestine on the other hand. To detect such compartment-specific eQTL, we focused our attention on modules that were significantly active in (i) lymphocytes, or (ii) macrophages/monocytes/dendritic cells isolated from the intestinal biopsies, but not in the equivalent cell types isolated from blood. We didn’t add the mirror comparison, i.e., modules active in blood but not in gut, because we assumed that we had more (statistical) power to detect eQTL in blood than in gut. The absence of a detectable eQTL effect in the gut could more often be a trivial power issue. We detected several instances supporting the existence of such compartment-dependent eQTL (STable 18). Part of these seem to involve induction of gene expression upon entering the intestinal compartment (scenario 1), others seem to be independent of gene expression level but a genuine conditional effect of the regulatory variants (scenario 2). We illustrate both scenarios using *CCL24* in monocytes/macrophages and *CCL20* in T lymphocytes, two CC-motif chemokines with chemotactic and antimicrobial activity whose genes show to have higher expression and be under specific genetic *cis*-control in the gut (Fig. 4C and 4D).

### Using inferred cell-type ontogeny to effectively map eQTL using scRNA-Seq data

Obviously, the observed degree of eQTL cell type specificity will depend on the chosen cell type granularity. This becomes particularly pertinent when working with single cell (ultimate granularity) RNA-Seq data: if one splits a cell cluster in sub-clusters, what was an eQTL specific for the cluster may now become shared by the sub-clusters (and hence apparently less cell type specific). Deciding at what cluster resolution to perform eQTL analysis will also have a considerable impact on eQTL detection: considering two clusters that share an eQTL separately rather than together, may decrease the detection power if the number of cells in each cluster is power limiting. The optimal cell partitioning strategy to detect a given eQTL/module will depend on where (i.e., in which cell types) the eQTL/module is active. If an eQTL is active in all cell types, the best strategy is to consider all cells jointly. If an eQTL is specific to a very small subset of cells, merging these cells with others that do not express the eQTL will reduce detection power. To address these issues in a generic way, we decided to construct a hierarchical tree of cell clusters based on the similarity of their transcriptomes. We assumed and demonstrated that this tree largely reflects cellular ontogeny. We also assumed that regulatory modules are turned on at specific developmental stages (i.e., nodes or leaves of the tree) and affect at least part of downstream branches along variable length. Accordingly, we performed eQTL analysis separately for all leaves and nodes of the tree. This effectively guides and limits the cell pooling options in an ontogenic framework, yet allows informed exploration of many possible scenarios. Of note, the proposed method disconnects eQTL mapping from cell type annotation.

The eQTL detection step was followed by the merging of similar EAP into modules, and the visualization of where a given module is active along the hierarchical tree (see f.i. Fig. 3F). Assuming that the module has been turned on once along the ontogenic tree, the signal should be the strongest for the node corresponding to the ontogenic stage where it was turned on. The signal should become weaker as one moves towards the root as the cells in which the module is active are progressively diluted by cells in which it is not. It should also become weaker as one moves towards the leaves as the number of cells for analysis decreases. Thus, one could assign the module to the segment in the tree where the detection signal is maximal. In reality we didn’t see the predicted smooth decrease of signal strength up and down-wards (in the tree) from a point of maximum significance, presumably because signal strength is affected by a multitude of other factors. We therefore chose to assign the module to the most recent common ancestor (MRCA) of all active leaves/nodes, which may result in positioning the modules too much towards the root.

The number of nodes/leaves in which modules were active showed a clear sign of overdispersion with many modules being either active in fewer nodes/leaves than expected by chance, or active in more nodes/leaves than expected by chance. As for blood, the excess of modules active in fewer than expected nodes was unlikely due to statistical power issues as augmenting modules with matching yet less significant eQTL didn’t affect this pattern. We believe that this indicates that many regulatory modules are cell type specific in the gut as previously observed for the FACS/MACS sorted circulating immune cell populations. In particular, enterocytes and stromal cells from the small intestine differ considerably from those of the large intestine, both with regards to transcriptome (Fig. 2E) and eQTL activity (Fig. 3D).

### Increasing cell-type granularity uncovers new IBD-driving regulatory modules

The initial premise of this work was that a large fraction of risk loci remained “orphan” thus far, because the disease driving-eQTL were active in cell types that were absent or underrepresented in previous eQTL datasets. The observation that a large fraction of eQTL/modules indeed appear to be highly cell type and even context specific supports this hypothesis. Accordingly, this work in essence doubles the number of IBD risk loci with DAP-matching eQTL to 140, i.e., ∼70% of the 206 studied risk loci. For approximately half of risk loci (55%), DAP-matching EAP were detected in both blood and gut. For the remaining half, there were slightly more matches in blood (24%) than in the gut (21%). Thus, scRNA-Seq-based eQTL detection in the gut made a considerable contribution to DAP-EAP matching despite the fact that we analyzed samples from only 57 individuals (yet in three locations). It seems reasonable to assume that increasing intestinal scRNA-Seq sample size will uncover DAP-matching EAP for additional risk loci, and meta-analyses towards that goal are in progress.

An additional factor that has contributed to the marked increase in the number of IBD risk loci with DAP-matching EAP is the splitting of risk loci into sub-risk loci. This was done for 32 of the 206 studied risk loci, because we assumed that “multimodal” DAP might be the sum of multiple independent EAP. This strategy allowed us to detect an extra 28 DAP-matching e-genes, covering an additional nine IBD risk loci.

It has been argued that it may be more effective to perform eQTL analyses in fewer, easily collectable sample types (f.i. whole blood) but from many more individuals, than to increase sample types yet remain limited in the number of individuals, to uncover more DAP-matching EAP. To verify this, we used PBMC eQTL summary statistics from ∼30,000 healthy individuals of European descent [Võsa *et al*., 2021], and searched for DAP-matching EAP using a slightly more permissive procedure as the one used with our own expression data. Using Võsa’s data, we identified DAP-matching EAP for 39 loci involving 59 genes. Using our own PBMC information (i.e., 187 individuals), we identified DAP-matching EAP for 32 IBD risk loci involving 42 genes, with 12 overlapping loci and three overlapping genes. Using our full blood data set (27 immune cell populations), we identified DAP-matching EAP for 110 loci involving 310 genes, of which 36 loci and 27 genes overlapping with Võsa. Using our complete data set (blood and gut), DAP-matching EAP for 140 loci involving 556 genes, of which 38 loci and 30 genes overlapping with Võsa. Thus, the Võsa data enabled the identification of matching EAP for one IBD risk locus (rs1479918), and 29 e-genes that were missed with our data. However, we identified matching EAP for 102 IBD risk loci, and 526 e-genes that were missed with Võsa’s data. These findings suggest that a large number of additional matches are uncovered when performing eQTL analyses in isolated cell types.

### Are the e-genes controlled by IBD-driving regulatory modules causal?

The initial assumption, when eQTL studies to identify causative genes in risk loci were initiated, was that DAP-matching EAP would occur rather exceptionally, yet - when detected - would provide strong evidence for gene causality. It now appears that DAP-matching EAP are rather common, and that for many risk loci, DAP-matching EAP are found for multiple genes (STable 21&24). Moreover, we find in this work that when, for a given risk locus, DAP-matching EAP are found in both blood and gut, the genes involved are largely different. Are all of these genes, one way or the other, causally involved in disease risk, only some, or none? In other words, what is the proportion of “red herring” eQTL [Connally *et al*., 2022] amongst DAP-matching EAP? All scenarios are plausible, and each one likely applies to at least some risk loci.

It seems possible that - because *cis*-acting regulatory elements are often shared by multiple genes [f.i. Thurman *et al*., 2012] – many regulatory variants will affect the expression of neighboring genes including some that have no bearing on disease risk. The relatively modest signals obtained by pathway enrichment analyses (STable 23) supports the assertion that a sizable proportion of DAP-matching e-genes are not directly influencing disease risk. It is tempting to assume that DAP-matching information will be more specific for risk loci with fewer matching e-genes, and this information is available from STable 21&24.

On the other hand, when scanning the literature for functional evidence supporting the causal involvement of genes in our candidate list, we were struck by the large number of DAP-matching e-genes with such evidence (212 out of the 483 examined coding genes). Several risk loci harbor more than one gene with quite enticing support (see examples, in results, of loci with strong functional candidates that additionally harbor less well-known genes underpinning early onset, Mendelian forms of IBD). Thus, it seems likely that – at least for some risk loci – the risk variants are affecting the expression of multiple genes that jointly affect the risk to develop IBD. In other words, the notion of polygenicity may not be limited to the fact that disease risk is affected by multiple loci in the genome, but additionally that (at least some) risk loci harbor multiple causative genes controlled by the same or distinct regulatory modules. An additional level of complexity may result from the fact that a single gene may affect disease outcome through multiple pathways, operating for instance in different cell types, triggered by the same or by different regulatory modules. For example, *INAVA* was shown to play a pivotal role in PRR-induced signaling, cytokine secretion and bacterial clearance in peripheral macrophages and intestinal myeloid cells [Yan *et al*., 2017], yet at the same time to regulate the stability of adherens junctions of intestinal epithelial cells (where we see the best DAP-match) [Mohanan *et al*., 2018]. Also, *ATG16L1* is increasingly understood to affect disease outcome through autophagy-dependent but also -independent mechanisms [Hamoui *et al*., 2022], in agreement with our observation of a DAP-match in eosinophils but also colonocyte precursors. The suggestion that specific risk loci may affect disease outcome through multiple genes and pathways is also well illustrated by the *CCR6*/*RNASET2* and *IRGM/CD74/TNIP1* gene sets.

Is it possible that for some risk loci, none of the DAP-matching e-genes are causal? Positional cloning has been pursued very successfully during the last 30 years, under the assumption that causative mutations always affect the function of a causal gene in *cis*. A key assumption of the omnigenic model [Boyle *et al*., 2017; Liu *et al*., 2019] is that variants can affect phenotype without having any causal *cis*-effect on the expression of neighboring genes. That doesn’t mean that one will not see *cis*-eQTL effects on neighboring genes, but rather that these do not influence the phenotype: a sobering thought for positional cloners.

Of note, we didn’t see an effect of the number of DAP-matching genes, with or without reported function, per risk locus on the magnitude of its effect on disease (measured by the odds ratio (OR)). This is not surprising given that many factors will affect OR, but a positive result would have been in support of the “multigenic” nature of individual risk loci whether being causal *per se* or not (i.e., omnigenic model).

### Are multi-genic and multi-tissular regulatory modules underpinning pleiotropy?

The number of DAP-matching e-genes is remarkably high for some risk loci, often despite any obvious functional theme or coherence. This suggests that risk variants may hit master *cis*-regulators that control the expression of large chromosome domains and many genes therein, irrespective of function. We reasoned that such variants might, because of the number of affected genes, influence multiple traits, i.e., be pleiotropic. Of interest, under this scenario, the mechanism accounting for pleiotropy would only involve the variants, not the e-genes affected in *cis*, as causative genes would differ between traits. It is well established that as many as 90% of GWAS-identified risk loci affect multiple traits, that can even belong to distinct “trait domains”. We searched for a correlation between the number of DAP-matching (IBD) e-genes and the number of trait domains pleiotropically affected by the corresponding risk loci as reported in Watanabe *et al*. [2019] (SFig. 8C). There was no obvious relation between these two statistics. Thus, at first glance, it does not seem that risk loci with high numbers of DAP-matching e-genes make a disproportionate contribution to pleiotropy.

Regulatory modules that are active in all cell types, make a disproportionate contribution to the risk loci with DAP-matching EAP, particularly in blood (SFig. 8H and 8I). We made a similar observation for IBD risk loci with the less-granular CEDAR1 data set [Momozawa *et al*., 2018]. This is possibly due to the fact that detecting the DAP-match is less dependent on analyzing the correct (i.e., disease-relevant) cell type for these risk loci.

### Why are DAP-matching e-genes not showing an excess burden of coding variants in patients?

As discussed before, being a DAP-matching e-gene does not prove that it is causally involved in disease risk. There are two formal tests of gene causality. The first is the reciprocal hemizygosity test, which is very difficult to apply in mammals, let alone humans [Steinmetz *et al*., 2002; Stern et al., 2014]. The second is the enrichment of disruptive coding variants in cases. For example, this test was used to demonstrate the causality of *NOD2* in CD [Hugot *et al*., 2011; Lesage *et al*., 2002]. Therefore, a logical next step after the identification of DAP-matching e-genes, is to sequence the corresponding genes in large case-control cohorts and to perform rare variant-based burden tests [f.i. Momozawa *et al*., 2018]. As a matter of fact, this approach has now been applied genome-wide (i.e., without preselection of target genes using eQTL information) for several common complex diseases on very large cohorts including IBD [Sazanovs *et al*., 2022]. Although some new causative genes have been identified using this approach (including some that overlap with DAP-matching candidates, including from this study), the yield has been, arguably, somewhat disappointing given the magnitude of the effort. The same applies to most diseases for which this approach has been used [f.i. Flannick *et al*., 2019]. We herein have used Sazanovs’ resequencing data to look for the distribution of burden test *p*-values for 298 candidate e-genes out of our list of 556. There was no evidence from a departure from expectation under the null (Fig. 5C). Does that mean that none of our DAP-matching e-genes are affecting IBD risk? Although we cannot completely exclude that possibility, alternative explanations exist. One is a power issue: sequencing 35,000 cases is a lot but may not be sufficient, especially for small genes, and given the fact that predicting the effect of coding variants other than stop-gains remains difficult. The other is that coding variants in the corresponding genes do not cause IBD but possibly other diseases. A fundamental difference between coding and regulatory variants is that coding variants affect the function of the gene equally in all tissues where the gene is used, while regulatory variants likely affect the function of the gene in a restricted set of tissues. It is increasingly apparent that most genes are utilized in many tissues and cell types, as testified by the many synonyms existing for most gene names. As indicated by their denomination, diseases such as IBD are organ-restricted in their manifestations. It is very possible therefore that to be risk variants for IBD the effects have to be organ restricted (gut and immune cells), and that – for many genes - this does not apply to coding variants. The target space to resequence may therefore have to be redirected to (cell type-specific) *cis*-acting regulatory elements, yet these remain difficult to identify, the effects of variants on their functionality difficult to predict, and the corresponding genome space limited (hence affecting power).

### Genetic support for new repurposing opportunities in IBD

We identify entrectinib as a promising repurposing candidate for CD. Entrectnib (ENB) is a small molecule that blocks several tyrosine kinases including oncogenic ones. It has been approved by the FDA in 2019 for the treatment of several solid tumors. It is administered orally and has proven safe and well tolerated even at high doses and prolonged administration. More recently, a screening of FDA-approved kinase inhibitors, showed that ENB specifically blocks the NRLP3 inflammasome. This was shown to result from the reversible binding of ENB to R121 of the NEK7, thereby inhibiting NRLP3 activation. Paradoxically, ENB does not affect NEK7’s kinase activity, which increases the effect’s specificity. *In vivo* tests show that shRNA-mediated downregulation of NEK7 protects mice against DSS-induced colitis, while ENB was shown to protect mice against various NRLP3-dependent inflammatory conditions [Chen et al., 2019; Jin et al., 2023]. Thus, there is considerable prior functional and preclinical in vivo evidence supporting the use of ENB to treat IBD. In here, we show that CD risk variants increase NEK7 transcript levels in circulating naïve B cells, with very strong support for “colocalization” (*θ*_*IBD*_=0.94). We also show that expression levels of *NEK7* are affected in multiple circulating immune populations of active CD patients, and – using the data from Kong *et al.,* [2023] – are increased in the colonic epithelium of IBD patients. In addition to supporting a contribution of *NEK7* expression levels in influencing predisposition to IBD, this supports NEK7’s role in the disease process *per se*, and hence a more likely curative effect of ENB. We note, however, that the IBD risk variants appears to decrease *NEK7* expression in some gut-resident cells of healthy individuals, while *NEK7* expression is decreased in some circulating immune populations, and these findings deserve further scrutiny.

We further identify at least two instances, amongst the targets with approved drugs, where IBD risk variants, rather counterintuitively, increase the expression levels of what are assumed to be positive mediators of inflammation, in particular *IL18R1* and *IL18RAP* on the one hand, and *PTGIR* on the other (Supplemental material). Thus, “hypomorphic” IL18 and prostacyclin pathways may increase the risk to develop IBD, despite the fact that these pathways participate actively in the inflammatory process once initiated. These observations obviously call for caution when intending to use activators of the corresponding pathways to treat active IBD patients. It suggests, however, that treatment of IBD patients in active phase versus relapse should be differentiated. It is conceivable that activation of the IL18 and prostacyclin pathways might help to prevent relapse, particularly after disease remission has been achieved following surgery.

Possibly the most important outcome from this work is the suggestion that the notion of polygenicity extends *within* risk loci, and that causative genes in risk loci may influence disease through their effects on multiple cell types. It supports the notion that the genetic determinism of CCD is “quasi-infinitesimal”. This raises the question as to whether targeting individual components of this genetic architecture to treat the disease is the most effective strategy. As mentioned before, it seems unlikely that the effect of the many underpinning risk variants on disease are independent. They must perturb a series of pathways that progressively converge into a limited number of “highways”, that ultimately determine disease outcome. Such “highways” would involve the “core genes” as defined in the omnigenic model [Boyle *et al*., 2017]. It should, in theory, be possible to genetically reconstruct the topology of the corresponding network. Indeed, just as the disease is associated with multiple risk loci (multiple DAP) “in *trans*”, components of the upper part of the network (the “highways”) are also expected to be associated with multiple risk loci as *trans-*QTL. On the other hand, components of the lower parts of the tree will be associated with fewer risk loci, and ultimately, i.e., at the bottom of the network, only with one (for at least some, as a *cis*-eQTL). It seems that the components of the upper parts of the network (i.e., the “highways”) would make for better drug targets than those at the bottom of the tree. Of note, the abundance of the components of the upper part of the network are expected to be most strongly correlated with polygenic risk scores for the corresponding disease and – if conveniently measurable - may constitute biomarkers capturing both genetic and environmental effects.

## Data Availability

All data produced in the present study are available upon reasonable request to the authors

## Methods

### Identifying cis-eQTL modules in bulk RNA-Seq data of 27 sorted circulating immune cell populations

#### Sample collection

We collected 40 ml of venous blood (EDTA) from 251 healthy European subjects at the academic hospital of the University of Liège (CHU) between October 2018 and November 2022. Written informed consent was obtained prior to donation in agreement with the recommendations of the declaration of Helsinki for experiments involving human subjects. The experimental protocol was approved by the Ethics committee of the CHU Liège (reference number: 2017/214). Data collected from the electronic medical records (EMR) included birth date, age at sampling, ancestry, sex, weight, height, smoking and alcohol history, declared ethnicity, family history of disease, surgical and medication history, blood type when available and known allergies. The following hematological parameters were measured: counts of white blood cells, neutrophils, lymphocytes, monocytes, eosinophils, basophils, platelets, red blood cells, hemoglobin concentration, hematocrit, mean cell volume, mean cell hemoglobin, mean cell hemoglobin concentration, red cell distribution width and mean platelet volume (STable 1).

#### SNP genotyping

Genomic DNA was isolated from frozen EDTA-blood using the NucleoMag Blood 200 μL kit (Macherey-Nagel) on a KingFisher robot (Thermo Fisher Scientific). Individuals were genotyped for 713,606 SNPs using Illumina’s Human OmniExpress BeadChips, an iScan system and the Genome Studio software following the guidelines of the manufacturer. We confirmed the European ancestry of the participants by PCA (using the HapMap population as reference) as well as the absence of duplicated or related individuals(pi hat > 0.185). All individuals had less than 3% of missing genotypes. We excluded variants with call rate ≤ 0.95 or deviating from Hardy–Weinberg equilibrium (p ≤ 3 x 10^−4^) using plink (v1.9), leaving 689,223 quality-controlled variants. After lifting over to GRCh38 using Picard LiftoverVcf (v2.7.1), we phased and imputed to whole genome using the TOPMed Imputation Server (v1.6.6) and the TOPMed r2 reference panel. We removed variants with imputation score (R²) ≤ 0.7 or minor allele frequency (MAF) ≤ 0.05 using bcftools (v1.11), leaving genotypes at 6,299,998 QC-ed variants.

#### Cell sorting

Granulocytes were isolated from blood using the EasySep™ Direct Human Pan-Granulocyte Isolation Kit (StemCell Technologies #19659) within one hour after collection. Neutrophils and eosinophils were then isolated from the recovered granulocytes using the EasySep™ Human Neutrophil Isolation Kit (StellCell Technologies #17957) and EasySep™ Human Eosinophil Isolation Kit (StemCell Technologies #17956), respectively. Cells were recovered in cell homogenization buffer provided with the Maxwell 16 LEV simply RNA Tissue Kit (Promega #AS1280) or the AllPrep DNA/RNA Micro Kit (Qiagen #80284), and immediately frozen at - 80°C until use.

Peripheral blood mononuclear cells (PBMC) were isolated from fresh blood using SepMate™-50 (IVD) collection tubes (StemCell Technologies #85460) and Lymphoprep™ density gradient medium (StemCell Technologies #07861). After two washing steps, PBMC were stained with two panels of antibodies for 30 minutes at 4°C (STable 28). Cells suspended in PBS were then filtered using a 100 µm CellTrics filter (Sysmex #04004-2328). Cell sorting was performed on a FACS Aria III instrument (BD Biosciences) calibrated using CS&T beads (BD Biosciences). Fluorescence compensations were performed using CompBeads (BD Biosciences #552843). After exclusion of debris and doublets, we targeted monocytes (classical, non-classical and intermediate), T lymphocytes (including *γδ*, mucosal-associated invariant T cells (MAIT) cells, naive and memory regulatory T cells, naive and memory CD8+ T cells, naive and memory CD4+ T cells, Th1, Th2, Th17 and Th1/17 helper T cells), B lymphocytes (naive, memory and plasmocytes), dendritic cells (plasmacytoid and myeloid), natural killer cells (NK and NKT) and innate lymphoid cells (ILC). Panels of antibodies used, definition of sorted cells and gating strategies are described in SFig. 1 and STable 28. Purity of the sorted cell populations ranged from 78% to 99%. Up to 20,000 cells of each cell population were sorted directly on the cell homogenization buffer and frozen immediately at -80°C until use.

#### Bulk RNA sequencing

Total RNA was purified from sorted cells using the Maxwell 16 LEV simplyRNA Tissue Kit (Promega #AS1280) on a Maxwell 16 instrument (Promega) or manually using the AllPrep DNA/RNA Micro Kit (Qiagen #80284) with the QIAshredder (Qiagen #79656), according to their respective manufacturer’s instructions. RNA quantity was determined for all samples using the Quant-iT RiboGreen RNA Assay Kit (ThermoFisher Scientific #R11490), while the RNA quality has been evaluated for a subset of samples using the RNA 6000 Pico Kit on a 2100 Bioanalyzer instrument (Agilent #5067-1513). Reagents for cDNA and library preparation were dispensed with a Mantis Liquid Handler (Formulatrix) using half the recommended volumes. Full-length cDNA was generated from 1 ng of total RNA using the SMART-Seq HT Kit (Takara #634436), which poly-A selects mRNAs. Obtained cDNA quantity and quality were determined for all samples using the Quant-iT PicoGreen dsDNA Assay Kit (ThermoFisher Scientific #P7589), and for some using the High Sensitivity DNA kit on a 2100 Bioanalyzer instrument (Agilent #5067-4626), respectively. Uniquely indexed libraries were constructed from 300 pg of cDNA using the Nextera XT DNA Library Preparation Kit (Illumina #FC-131-1096) and custom-made 24 forward and 16 reverse primers. The libraries’ quantity was assessed for all samples by qPCR using a KAPA Library Quantification Kit (Roche#07960140001), while the libraries’ quality was checked for all samples using a QIAxcel Advanced technology (Qiagen). The libraries were pooled and sequenced on a NovaSeq 6000 instrument (Illumina), to an average read depth of 11 ± 5 million paired-end reads per sample (University of Geneva core facility (Geneva): 2×50 bp (2,112 samples); GIGA Genomics platform (Liège): 2×150 bp (3,180 samples)). We performed RNA-seq for 5,292 samples, corresponding to 27 cell types from 196 individuals.

#### Read mapping and quantification

Demultiplexing and FASTQ conversion were performed using bcl2fastq (v2.20). Read quality was assessed with FastQC (v0.12.1) and multiQC (v0.9). Reads were mapped to the GRCh38 (Ensembl release 105) human genome build using STAR (v2.7.1a). The STAR re-implementation of the WASP algorithm was used with a custom VCF file containing both reference and alternative alleles for all SNPs. The alignments that did not pass WASP filtering or that overlapped indels were removed from the resulting BAM files using samtools (v1.9). Alignment metrics were collected using Picard CollectRnaSeqMetrics (v2.7.1) (STable 3). Matching of genomic and transcriptome genotypes was evaluated with QTLtools mbv (v1.3.1) (SFig. 2A). Unstranded gene counts were generated with HTSeq (v0.6.1p1). If samples were split across two sequencing lanes, we summed up the respective counts. To detect mislabeled samples, reads counts were normalized using the variance stabilizing transformation from the DESeq2 R package and a t-SNE analysis was conducted using the 500 most variable genes (SFig. 2). STable 3 reports the characteristics of the 5,030 RNA-seq libraries that passed the quality control procedure.

#### Transcriptome-based hierarchical clustering of circulating immune cell types

A dendrogram was constructed based on the RNA-seq data, using 1-|Spearman’s correlation| between average (across all individuals) gene expression levels, using the “average”, “ward.d” and “ward.d2” methods. For each method, we built 32 dendrograms with from 250 to 8000 (by steps of 250) genes with best F statistic (cell type effect) from ANOVA. Within each method, we assessed the reliability of each dendrogram in a way inspired by the bootstrap procedure: for each node in the dendrogram, we computed the proportion among the 31 other trees that shared the same split. Within each method, we selected the dendrogram(s) with the highest sum of “bootstrap-like” values. Memory CD4 T cells, PBMC and granulocytes were ignored in this analysis as they encompass multiple cell types.

#### *Cis*-eQTL analyses

For each blood cell type, we filtered out the genes with less than 5 counts in more than 80% of samples, normalized the raw read counts using the DESeq2 R package and residualized them for age, sex, RNA extraction method, proportion of reads in each sequencing batch, top 3 genotype principal components (PCs) and top 13-36 expression PCs to maximize the number of *cis*-eQTLs. STable 4 reports the number of samples, genes and top expression PCs for each cell type. We removed variants with MAF ≤ 0.05 in the retained samples using bcftools (v1.9) for each cell type. We performed eQTL mapping using QTLtools in a 2 Mb window centered at the transcription start site and with the integrated rank normal transformation of the phenotypes. The *p*-values were corrected for multiple testing within each window by permutation (10,000 permutations) and within each cell type using the false discovery rate (FDR). eQTL with “within cell type FDR” ≤ 0.05 were considered significant.

#### Proportion of expression variance explained by *cis*-eQTLs

The proportion of expression variance was computed as 2*pqβ*^2^, where *p* and *q* are the allelic frequencies of reference and alternate allele, respectively, and *β* is the slope of the regression line of standardized gene expression (mean: 0, variance: 1) on dosage of the alternate allele, i.e., the allele substitution effect. This gave near identical results as QTLtools (r-squared values).

#### Agglomerating *cis*-eQTL in gene-specific and across-genes *cis*-acting regulatory modules (RM)

To assess if the expression levels of a given gene are affected by the same regulatory variants in a given pair of tissues, we compared the corresponding eQTL association patterns (EAP) using the *θ* metric devised in Momozawa *et al*. [2018]. Two EAP were compared if at least one of them was significant (FDR ≤ 0.05). We only used variants that had a *p*-value below 0.05 in at least one of the two EAP to compute *θ*. Two EAP were assumed to be part of the same *cis*-acting regulatory module (RM) if |*θ*| ≥ 0.6 (cfr. Momozawa *et al*. [2018]), and if the window-adjusted *p*-value of the eQTL (for non-significant eQTL) was ≤ 0.12, as these parameter values were shown to yield an agglomeration FDR ≤ 0.05 in a permutation test (see DAP-EAP matching section, hereafter). To better describe the connectivity of the modules, we retrospectively also computed *θ* for pairs of non-significant EAP if they were assigned to the same module. We first constructed gene-specific modules, i.e., we only confronted EAP from the same gene yet from different cell types. In a second stage, we constructed across-gene modules by evaluating the similarity of the EAP of different genes in the same or in different tissues, using the union of the overlapping 2 Mb windows, and using the same threshold values as above. We defined a representative EAP for each module as the EAP with the lowest adjusted *p*-value for *β*. For all EAP in a module, the sign of *β* was compared to that of the representative EAP. If the modules encompassed more than one EAP, we performed a meta-analysis to combine the constituent EAP in a consensus EAP representing the module. For each variant of the complete window (2-4 Mb), we converted nominal *p*-values to *z*-score which we squared and summed across all EAP in the module. The corresponding sum was assumed to have a chi-squared distribution with degrees of freedom equal to the number of EAP in the module. When arithmetic underflow was reached for the *p*-values, the −*log*_10_(*p*) values were predicted from the *z*-scores using a local polynomial regression. RMs were then curated to remove the connections between non-similar EAP. The similarity between the EAP and their consensus EAP was assessed using *θ*. If |*θ*| < 0.6, the EAP were excluded from the module and the consensus EAP reconstructed. Similarity between excluded EAP was tested to allow them to form distinct “sub-RM”.

#### Quantifying the statistical significance of the overdispersion of module activity across cell types

We quantified the dispersion for the real data, as the variance of the sum of active cell types across modules. We then performed “permutations”, in the sense that – for a given cell type – the activity states (1’s and 0’s) were permuted between modules. This was repeated for all cell types, and the dispersion of that permutation computed as the variance of the sum of “active” cell types across modules. The statistical significance of the real dispersion was then defined as the proportion of permutations that yielded as large or larger variance than the real data.

#### Assigning modules to nodes and leaves in the ontogenic dendrogram

Gene-specific modules were assigned to nodes and leaves of the best supported ontogenic tree (SFig. 2C). Modules that encompassed only one cell type were assigned to the leaves of the tree while modules that encompassed more than one cell type were assigned to the node corresponding to the most recent common ancestor (MRCA) of those cell types. Descendants of such MRCA nodes that were not encompassed by the module were assumed to have lost the eQTL effect. If these were part of another regulatory module for the same gene, the “loss” was converted to a “switch”. Memory CD4 T cells, PBMC and granulocytes were ignored in this analysis (cfr. above).

#### Testing for an excess sharing of RM between cell types

We followed Momozawa *et al*. [2018] to test whether specific cell types were sharing *cis*-eQTL more often than expected by chance (expected for ontogenically related cell types). In our analysis framework, this would manifest itself by the fact that the corresponding cell types would be co-included in the same RM more often than expected by chance. The number of *cis*-eQTL detected by cell type differs, and this has to be taken into account when measuring enrichment. We assumed that if sharing of eQTL was equally likely for any pair of cell types (accounting for differing numbers of eQTL per cell type), the proportion of sharing events between cell type *i* and *j* should correspond to the proportion of eQTL detected in cell type *j* (*n*_*jT*_) out of all eQTL detected in all cell types other than *i* 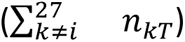. Imagine that cell type *i* is characterized by a total of *n*_*iS*_ sharing events, where 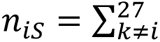 *n*_*ik*_, and *n*_*ik*_ is the observed number of sharing events between cell type *i* and *k*. We determined the probability to obtain *n*_*ij*_ or more “successes” under the null, by sampling *n_is_* events with a probability of success of 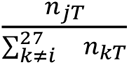 by simulation (*n* = 5,000). Of note, this process yields two *p*-values for every pair *i*, *j*: one obtained when considering *i* as reference, the other when considering *j* as reference. As in Momozawa *et al*., we performed the analysis 26 times: first considering RM with no more than two cell-types (hence eQTL that are shared by two cell types only), then considering RM with no more than three cell-types, etc., until considering RM encompassing all cell types.

#### Probing the causes of the cell type-specificity of gene-specific RM

Why is a *cis*-eQTL for gene “X” detected in cell type *a* but not *b*? We distinguished three possible scenarios: (i) Module switch: gene “X” is subject to a *cis*-eQTL effect in both cell type *a* and *b*, but the variants involved are distinct (i.e. dissimilar EAP), and the two *cis*-eQTL are assigned to different RM, (ii) Lack of expression cell type *b*: gene “X” is expressed at too low levels in cell type *b* to allow for the detection of a *cis*-eQTL effect, and (iii) Conditional eQTL effects: gene “X” is expressed at sufficient levels in cell type *b*, but there is no evidence for a *cis*-eQTL effect (significant variants x cell type interaction effect). To test the statistical significance of the third scenario, we first measured the *cis*-eQTL effect at the top variant for cell type *a*, in cell types *a* and *b*, yielding *β*_*a*_ and *β*_*b*_. We then generated bootstrap samples from cell type *a*, and computed 1,000 *β*_*s*_’s. The *p*-value of the variant x cell type interaction was determined as the number of *β*_*s*_-values that would be equal or lower than *β*_*b*_ if *β*_*a*_ was positive, or equal or higher than *β*_*b*_ if *β*_*a*_ was negative.

### Identifying cis-eQTL modules in single-cell RNA-Seq data from intestinal biopsies at three anatomical locations

#### Sample collection

Gut biopsies were obtained from 60 healthy adults (27 females and 33 males, average age was 54 years ranging from 23 to 75) that were visiting the university hospital of the University of Liège as part of a screening campaign for colon cancer between June 2019 and December 2021. Written informed consent was obtained prior to donation in agreement with the recommendations of the declaration of Helsinki for experiments involving human subjects. The experimental protocol was approved by the Ethics committee of CHU Liège (reference number: 2017/214). Two to four biopsy “bites” were collected from rectum (RE) and transverse colon (TC) for all participants while biopsies from the terminal ileum (IL) were obtained for 52. Biopsies were collected in 40 ml of RPMI-1640 culture medium (Lonza Bioscience, 12-167F) supplemented with 2 mM L-Glutamine (Thermo Fisher Scientific, 25030024) and 10% FBS (Sigma, F7524) on ice, and processed freshly within one hour from the time of colonoscopy. Data collected from the electronic medical record (EMR) were the same as for the individuals providing blood samples (STable 1).

#### SNP genotyping

Was conducted using the same procedure as for the circulating immune cell population samples (see above). We kept 686,493 SNPs interrogated by Illumina’s OmniExpress array after quality control, while genotypes at 6,352,658 variant positions were kept after imputation and quality control.

#### Single-cell RNA sequencing

Biopsies from the three locations (IL, TC, RE) were processed in parallel using so-called two-step protocols [Smillie *et al*., 2019; Kong *et al*., 2023] with some modifications. <u>Fractionation of epithelial (EC) and lamina propria (LP) cell layers:</u> The biopsies delivered in the transport media were collected by passing the media through a 100 μm cell strainer (Pluriselect Life Science, 43-50100-50), and transferred to a 50 ml EZFlip tube (Thermo Fisher Scientific, 10571663) with 25 ml of pre-warmed Epithelial Strip Buffer consisting of HBSS (Thermo Fisher Scientific, 14170088), 5 mM EDTA (Thermo Fisher Scientific, AM9260G), 15 mM HEPES (Lonza Bioscience, 17-737E) and 5% FBS and a magnetic stirring bar. The sample was agitated with gentle stirring (130 rpm) for 10 min at 37°C by placing the tube upside down on a magnetic stirrer (Thermo Fisher Scientific, 50088009) in a 37°C incubator. After adding DTT to a final concentration of 1 mM (VWR, 443852A), the sample was incubated for another 10 min with agitation at 37°C. The sample was taken out of the incubator and shaken by hand vigorously for 10 – 15 seconds. It was passed through a 100 μm cell strainer to fractionate EC in the flow-through in a new 50 ml canonical tube, while the LP remained on the cell strainer. The LP sample on the strainer was rinsed with Washing Buffer (HBSS supplemented with 1 mM EDTA and 1% FBS) and kept on ice in a 6-well containing Wash Buffer. <u>Dissociation of EC:</u> The tube with the EC fraction was filled with ice-cold Washing Buffer up to 50 ml and centrifuged at 500 rcf for 5 min at 4°C. After carefully removing the supernatant, the sample was transferred to a 1.5 ml siliconized microtube (Sigma, T4816) using 100 μl of TrypLE Express enzyme solution (Thermo Fisher Scientific, 12604-013). The sample was then mixed ten times using a 200 μl tip and incubated in a water bath at 37°C for 5 min. The reaction was stopped by adding 1 ml of ice-cold Wash Buffer. EC were collected by centrifugation at 500 rcf for 5 min at 4°C, and resuspended in 100 μl of PBS (Lonza Bioscience, 17-516F) with 10% FBS. Cell concentration and viability was estimated by staining 10 μl of cell suspension with an equal volume of 0.4% Trypan blue solution (Lonza Bioscience, 17-942E) using either TC20 (Bio-Rad) or Countess 3 (Thermo Fisher Scientific) automated cell counters. We obtained an average of 4.1E+05, 2.6E+05 and 1.7E+05 EC cells with viability of 58%, 47% and 47% for IL, TC and RE, respectively. <u>Dissociation of LP cells:</u> The LP remaining on the cell strainer was transferred to a gentleMACS C tube (Miltenyi Biotec, 130-093-237) using 15 ml of pre-warmed Enzyme Solution consisting of HBSS supplemented with 2.5 mg of Liberase TL (Roche, 05401020001), 7.5 U of DNase I (Thermo Fisher Scientific, EN052) and 2% FBS. The LP tissue was dissociated using a gentleMACS Octo Dissociator (Miltenyi Biotec) with “37C_m_LPDK” program (∼ 25 min). Large debris were filtered out by passing the cell suspension through a 100 μm cell strainer into a new 50 ml tube. The tube was filled with ice-cold Washing Buffer up to 50 ml and spun at 500 rcf for 5 min at 4°C. After carefully removing the supernatant, the sample was treated with TrypLE Express enzyme and resuspended in 100 μl of PBS with 10% FBS as described above. We obtained an average 9.1E+05, 8.6E+05 and 5.5E+05 LP cells with viability of 74%, 78% and 77% for IL, TC and RE, respectively. <u>Cell hashing:</u> To reduce technical batch effects and costs of droplet-based scRNA-Seq [Stoeckius *et al*. 2018], we labeled each fraction of cells with distinct oligo-tagged antibodies and performed droplet formation using all fractions from a donor together in a single well of a 10X Genomics Chromium system. As we generally obtained larger numbers of cells from LP than EC, the LP cell suspension was divided into two or three tubes, while EC was kept in one tube (total 10 tubes). The total volume of the cell suspension per tube was adjusted to 90 μl using PBS with 10% FBS. The cell suspensions were first incubated with 5 μl of Human TruStain FcX Fc receptor blocking solution (BioLegend, 422301) for 10 min at 4°C, then mixed with 2 μl of 10 times diluted unique TotalSeq-B anti-human Hashtag antibodies (Biolegend; see also STable 10) and incubated at 4°C for 30 min. The cells were washed twice by adding 1 ml of PBS with 10% FBS and centrifuged at 400 rcf for 5 min at 4°C. The cells were re-suspended in 100 μl of PBS with 10% FBS and cell density and viability was estimated as described above. Equalized numbers of cells (average of 70,000 cells per tube) were pooled into one tube. In addition, 20,000 non-labeled cells were added in the pool, to provide base lines of hashtag reads when demultiplexing sequencing data. The cell pool was washed once more and re-suspended in 100 ∼ 400 μl of PBS with 10% FBS. After filtering through a 70 μm filter followed by a 40 μm cell strainer (Thermo Fisher Scientific, 22363548 and 22363547), cell density and viability were measured as described above. <u>Single cell RNA-Seq:</u> 37,000 cells (range: 15,000 ∼ 40,000) were loaded into one well of 10X Genomics droplet-based scRNA-Seq system and libraries were constructed by following the manufacturer’s protocol “Chromium Next GEM Single Cell 3’ Reagent Kits with Feature Barcoding technology for Cell Surface Protein, v3.0 or v3.1”. The libraries were sequenced for 546 million paired-end fragments on average for cDNA and 96 million fragments for Hashtags using either Illumina NextSeq 500 or NovaSeq 6000. The number of recovered cells was 12,436 on average (ranging 5,208 ∼ 44,126)(STable 10). <u>Variations on the main protocol:</u> 89 of 172 samples were treated using the procedure described above (“protocol 4”). 28 Samples were treated using “protocol 1” with the following specificities. After dissociating biopsies into cell suspensions, EC cell fractions were sorted by FACS (BD Biosciences, FACSAria III Cell Sorter) to enrich living EC and intraepithelial lymphocytes (IEL) using anti-human CD326 (Biolegend, 324226), CD45 (Biolegend, 304037), CD19 (Biolegend, 363034) and CD11b antibodies (Biolegend, 301348) along with Zombie Green Fixable Viability Dye (Biolegend, 423112). In parallel, LP cell fractions were sorted for living lymphocytes (LP-LC) and myeloid cells (LP-MC) by staining with anti-human CD326, CD45 (Biolegend, 304032) and CD11b antibodies and Zombie Green Fixable Viability Dye. The 12 fractions of sorted cells (EC, IEC, LP-LC and LP-MC for 3 locations) were labeled using distinct TotalSeq-A anti-human Hashtag antibodies (Biolegend) and all cell fractions loaded together on a single well of 10X Genomics Chromium system. 36 samples were treated using “protocol 2” with the following specificities: living cells from EC and LP fractions of cells were enriched using EasySep Dead Cell Removal Annexin V kit (Stemcell technologies, 17899). 19 samples were treated using “protocol 3”, with following specificities: living cells from EC and LP fractions of cells were enriched using FACS by staining cells with Zombie Green Fixable Viability Dye. <u>General precautions:</u> Samples were manipulated on ice unless described and using low retention filter tips (e.g., RAININ, 30389213). Cell strainers were dipped in FBS before use. Samples retained on a cell strainer were transferred by flipping the cell strainer on a collection tube and flushing it with a buffer.

#### Preprocessing of scRNA-Seq data

Raw sequencing data were preprocessed with the Cellranger software version 7.1.0 with standard parameters and GRCh38 (Ensembl release 103) human genome build as reference. Processed counts were further analyzed in R (version 4.3.1) within the Seurat tools ecosystem (Seurat version 4.1.3 [Hao *et al*., 2021]). For each sample, mRNA read counts and hashtag barcode read counts were loaded to R and quality-checked. We excluded (i) hashtag barcodes with < 300 reads per individual, (ii) cells with ≥ 50% mitochondrial reads, (iii) cells with < 200 different genes expressed, (iv) cells with < 5 hashtag barcode reads. Demultiplexing of cells was done using two different algorithms implemented in Seurat: HTODemux [Stoeckius *et al*., 2018] (positive.quantile 0.999, clusterization function - kmeans) and MULTIseqDemux [McGinnis *et al*., 2019] with automated threshold finding. Cells identified as singlets of the same tissue type by the two methods were kept for further analysis. To meet the RAM usage requirements, variable features selection was done in five sample batches with the “mean.var.plot" algorithm (3,000 features per batch). We retained the intersection between the five batches. We then integrated the sample batches in the space defined by the first 50 principal components using Harmony [Korsunsky *et al*., 2019]. The UMAP plots shown throughout the manuscript (f.i. Fig. 2D and 2E) are in this coordinate system. Yet, to better differentiate cell types and improve the clustering, we further split the data in two stages. We first used Hashtag information to separate cells by anatomical location (IL, TC, RE). Within each anatomical location we repeated the variable feature selection and integration process. In each of these three sub-datasets, we identified cell clusters with the Louvain algorithm. Based on marker gene expression, we assigned clusters to three groups: immune (expressing *PTPRC*), epithelial (expressing *EPCAM*), and endothelial plus other cells (expressing *VWF*, *PECAM1*, *CDH5* or not included in previous groups). Within each one of these nine sub-groups, we again repeated variable features selection, integration and clustering (Louvain clustering algorithm with resolution parameter 1.5). This yielded a total of 276 clusters across the nine data sets (SFig. 5).

#### Constructing a hierarchical tree of cell clusters

We computed, for each of the 276 location- and cell-type specific cell clusters (obtained as described above), the mean coordinate vector in the space of the first 50 Harmony coordinates. Then we computed the Euclidean distance between these vectors followed by hierarchical clustering (function hclust, stats R package, “complete” agglomeration method). The final dendrogram was constructed with the dendextend [Galili, 2015] R package.

#### *Cis*-eQTL analysis

We performed eQTL analysis for each leaf and node in the hierarchical tree, provided that the median cells per patient was ≥ 5 and that number of patients with cells in the leaf/node was ≥ 30. This left 401 leaves/nodes for eQTL analysis. Within each analyzed leaf or node, all cells were treated as pseudo-bulk, i.e., as if all reads were derived from one mega-cell. Resulting gene expression data were normalized using DESeq2, residualized for age, sex and five genotype PCs, and corrected for hidden confounders utilizing the probabilistic estimation of expression residuals (PEER) algorithm [Stegle *et al*., 2010]. Association between gene expression and alternate allele dosage was conducted for each SNP in a 2Mb-window centered on the gene’s transcription start site using QTLtools [Delaneau *et al*., 2017]. Nominal *p*-values were corrected for the realization of multiple tests in this *cis*-window by permutation, yielding a window-adjusted *p*-value for one lead SNP for every gene x leaf/node combination. Window-adjusted lead SNP p-values for all genes in a leaf/node were jointly used to compute a *q*-value using Storey & Tibshirani [2003].

#### Agglomerating *cis*-eQTL in gene-specific and across-genes *cis*-acting regulatory modules (RM)

The construction of *cis*-acting regulatory modules (RM) was done in a similar way as for the circulating immune cell populations, across all 276 leaves and 275 nodes of the hierarchical tree. We first build gene-specific and then across-gene modules. For gene-specific modules, we computed *θ* [Momozawa *et al*., 2018] between EAP obtained, for that gene, in the different nodes/leaves. *θ* was computed in the 2Mb *cis*-window centered on the gene’s transcription start site (TSS) (the window used for *cis*-eQTL analysis). For across-gene modules, we additionally computed *θ* between EAP of different genes, provided that their 2Mb *cis*-windows overlapped. *θ* was then computed for the union between the two 2Mb *cis*-windows. For both gene-specific and across-gene windows, we only considered EAP pairs for which at least one corresponded to a significant eQTL (within leaf/node FDR ≤ 0.05). We only considered SNPs with eQTL nominal *p*-value ≤ 0.05 for at least one of the two EAP. EAP (from the same or different genes) were merged in the same module if |*θ*| ≥ 0.6, and (in the case one of the EAP pairs did not correspond to a significant eQTL) a *p*-value of the eQTL corrected for the multiple SNPs tested in the window (by permutation, see above) ≤ 0.012, as this yielded mergers with FDR ≤ 0.05 (see DAP-EAP part, hereafter). To be part of a module, an EAP had to satisfy these criteria with at least one significant member of the module. Once a module was assembled (all member EAP determined), *θ* was computed between non-significant members of the module to evaluate the tightness of the module (ideally one hopes for |*θ*| ≥ 0.6 between all members; observed: “Proportion of links” in STable 13 and Stable 14). Links between pairs of EAP within a module were given a sign (positive or negative) depending of the value of *θ*. Constituent EAP were given a positive sign if their *θ* with the module’s representative EAP (the one with the most significant eQTL) was positive, a negative sign otherwise. For modules that comprised more than one EAP, we computed a consensus EAP (2-4 Mb window) by “meta-analysis”. *P*-values for all SNPs in the window were converted to z-scores, z-scores (for a given SNP) squared and summed over all members of the module. The resulting sum was converted back to a *p*-value assuming that it had a chi-squared distribution with numbers of degrees of freedom equal to the number of EAP in the module. The EAP of all constituent eQTL were confronted to the consensus EAP in the full window. An EAP was only maintained in the module if its |*θ*|-value with the consensus was ≥ 0.6. Otherwise, it was ejected from the module. Ejected EAP were confronted to each other and given the possibility to assemble in new “sub-modules”.

#### Cell type annotation and cell type to hierarchical tree map

Cell type annotation was largely done by visually inspection of the expression profiles of 49 cell type-specific gene signatures obtained from the literature [Smillie *et al*., 2019; Franzen *et al*., 2019; Burclaff *et al*., 2022; Ishikawa *et al*., 2022; Hickey *et al*., 2023; Kong *et al*., 2023; Krzak *et al*., 2023] using the Seurat AddModuleScore function [Tirosh *et al*., 2016], and the Azimuth human PBMC reference mapping program for immune cells [Hao *et al*., 2021], on our global UMAP (i.e., all 293,801 QC-ed cells) (Fig. 2D). In essence, the distribution of a cell type-specific gene-signature on the UMAP was confronted with the distribution of the cells from each node/leaf of our hierarchical tree, looking for best matches. A given cell-type was assigned to the best matching node (and hence all descendent nodes/leaves). We further distinguished location-specific (i.e., ileum, colon and rectum) nodes and leaves within cell type-specific sections of the tree. The workflow and outcome of this analysis are shown in SFig. 8 and Stable 11.

#### Assigning regulatory modules to intestinal cell types

Regulatory modules encompass one or more EAP that can be active in one or more nodes/leaves of the hierarchical tree. If all nodes/leaves in which a module is active belong to the same cell type (see previous section), the module was assigned to that cell type (and possibly anatomical location within cell type). If nodes/leaves belonging to a module correspond to multiple cell types, the module was assigned to one of 29 supergroups, listed in STable 11&17. As an example, if a module was active in CD4 and CD8, it was assigned to the T lymphocyte supergroup.

#### Exploring the distribution of the number of nodes/leaves in which regulatory modules are active

As for blood, each module is characterized by a vector of 0’s and 1’s informing us about the nodes/leaves in which the module is active. In the case of the intestinal scRNA-Seq data, the length of the vector is 155 leaves + 246 nodes = 401 elements. The length of the vector is less than the sum of the total number of leaves and nodes in the module, because some nodes/leaves didn’t have any active module in them. The sum of 1’s in a module vector, i.e., the total number of leaves/nodes in which the module is active, is what we are looking at in Fig. 3C. More specifically, we are looking at the distribution of this sum across all modules. Conversely, the activity of a leaf/node can be summarized by a vector of 3,345 (gene-specific modules) or 3,081 (across-gene modules) 0’s and 1’s, indicating which module is active in the corresponding leaf/node. To verify whether the observed distribution differs from the expected one, assuming that the activity of a module in a given leaf/node is independent of its activity in other leaves/nodes, we assigned the 0’s and 1’s of a given leaf/node randomly to the modules, i.e., we randomly permuted module id within leaf/node. We then summed the number of 1’s across the modules and examined the distribution of this sum across modules (gray bars in Fig. 3C). We know that the activities of a module in adjacent nodes/leaves in the tree are not independent, as these have cells in common. To properly evaluate the statistical significance of the apparent “overdispersion” of module activity (too many modules either active in few nodes/leaves, or in many nodes/leaves), we restricted to the analysis to “parent” nodes of the 49 distinct cell types, selected such that no cell could be part of more than one such cell type (i.e., none of the selected nodes is ancestor of any other one). We quantified the dispersion for the real data, as the variance of the sum of active nodes across modules. We then performed “permutations”, in the sense that – for a given node – the activity states (1’s and 0’s) were permuted between modules. This was repeated for all nodes, and the dispersion of that permutation computed as the variance of the sum of “active” nodes across modules. The statistical significance of the real dispersion was then defined as the proportion of permutations that yielded as large or larger variance than the real data.

### 3D plots of *cis*-eQTL activity

We developed an application to visualize the activity of an eQTL of interest on a 3D UMAP plot. Briefly, for each cell in the dataset, we identified the 100 nearest neighbors in the space defined by the 50 first expression principal components using the Annoy algorithm [https://github.com/spotify/annoy] implemented in the Seurat Findneighbors function. We eliminated cell-centered neighborhoods encompassing cells from only one individual, less than five cells with non-null expression for the e-gene of interest, and MAF < 0.05 for the eQTL’s top SNP amongst the individuals with cells in the neighborhood. We then performed eQTL analysis in the remaining neighborhoods, one at a time, using a mixed model [Bates *et al*., 2015; Kuznetsova *et al*., 2017] including allele dosage (for the eQTL’s top SNP) as fixed regression, and individual as random effect. For each neighborhood we then multiplied -log(*p*-value) of the eQTL effect by the sign of the regression coefficient (*β*), assigned it to the cell defining the neighborhood, and plotted it as the third, *z* dimension of a 3D plot, at the x-y coordinate position corresponding to the position of the reference cell in 2D UMAP space.

### Merging blood and intestinal cis-eQTL modules reveals eQTLs that are specific for gut-resident immune cells

#### Module construction

Constructing modules integrating blood and intestinal data followed the same procedure as for the blood- and gut-specific modules. Requirements for an EAP to join a module were the same as before, i.e., |*θ*| ≥ 0.6 for both blood and gut EAP, *p*_*θ*_ ≤ 0.05 for both blood and gut, *p*_*eQTL,window adj*_ ≤ 0.12 for blood and ≤ 0.012 for gut. These thresholds ensured an FDR ≤ 0.05 in the corresponding DAP-EAP confrontations (see hereafter).

#### Test of independence of cell type annotation in blood and gut

Modules were assigned to cell types separately for blood cell type populations (obvious) and intestinal cell type populations (using the same approach as for the intestinal modules). One expects a certain degree of coherence with regards to cell type assignment in both datasets. As an example, modules that are assigned to lymphocytes in blood are expected to be assigned to the lymphoid compartment in the intestine as well. We verified this concordance by performing an empirical test of independence. Cell types were groups in a limited number of “supergroups” (Blood: lymphoid, myeloid other than granulocytes, granulocytes, multiple cell types, undetected; Gut: lymphoid, myeloid, enterocyte precursor, mature enterocyte, stromal, multiple cell types, undetected; see also Fig. 4B). We first performed a test of independence within the group of 2,170 modules that were assigned to a supergroup in both data sets. We determined the proportion of modules assigned to each supergroup separately for blood 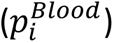 and gut 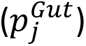. The observed number of modules assigned to supergroup *i* in blood and *j* in gut was then compared to the expected number computed as 2,170 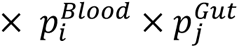. The probability to obtain an as large or larger deviation between expected and observed numbers by chance was determined from 1,000 replicates of 2,170 samplings with replacement with a probability of success of 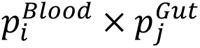. Secondly, within the group of 4,579 modules that were active in gut alone, we determined the proportion that were assigned to each one of the gut supergroups 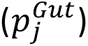 as well as the proportion that were not active in blood 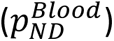. The observed number of modules assigned to supergroup *j* in gut yet undetected in blood was then compared to the expected number computed as 4,579 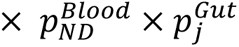. The probability to obtain an as large or larger deviation between expected and observed numbers by chance was determined from 1,000 replicates of 4,579 samplings with replacement with a probability of success of 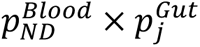. Finally, within the group of 22,336 modules that were active in blood alone, we determined the proportion that were assigned to each one of the gut supergroups 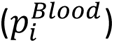 as well as the proportion that were not detected in gut 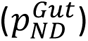. The observed number of modules assigned to supergroup *i* in blood yet undetected in gut was then compared to the expected number computed as 22,336 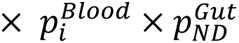. The probability to obtain an as large or larger deviation between expected and observed numbers by chance was determined from 1,000 replicates of 22,336 samplings with replacement with a probability of success of 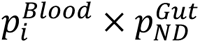.

### Identifying new cis-eQTL driving inherited predisposition to IBD

#### Comparing EAP and DAP using theta

We performed a colocalization analysis of our EAP with IBD, CD and UC risk loci coming from de Lange *et al*. [2017]. We lifted their data over from GRCh37 to GRCh38 and defined disease association patterns (DAP) in genomic locations where a risk locus was described in at least one of the diseases and where at least one variant had a *p*-value less or equal to 10^-5^. We established the limits of the DAPs manually to surround the peaks. In total, we tested 455 DAP corresponding to 206 risk loci (157 for CD, 173 for IBD and 125 for UC). Some DAP were subdivided into two or three parts. We evaluated the similarity between DAP and EAP using *θ* following Momozawa *et al*. [2018], for all EAP for which the top eQTL SNP was within the boundaries of the analyzed disease interval. *θ* was computed for all variants located within the limits of the disease interval, provided that their nominal association *p*-value ≤ 0.05 either in the EAP, DAP or both. To determine the statistical significance of *θ* (i.e., *p*_*θ*_), we performed up to (adaptive) 10,000 permutations (without replacement) of gene expression levels before recomputing *θ*. We defined *p*_*θ*_ as the proportion of permutations where the obtained |*θ*| is greater or equal to the observed.

To further define appropriate thresholds to declare a DAP-EAP match of interest, we repeated the full, genome-wide *cis*-eQTL analysis, both in blood and gut (i.e., in the 27 cell types and 551 leaves/nodes), after randomly disconnecting (i.e., permuting) the genotype (all variants) and expression (all genes) vectors. Genotype and expression vectors were maintained unaltered (hence LD structure on the one hand, and correlation structure between gene expression on the other hand, were conserved). We then repeated the colocalization exer*cis*e between all 455 DAPs, and the overlapping EAP obtained with the permuted data exactly as we did with the real data. Assume that a DAP matches a real EAP with |*θ*| = *x* ≥ 0.6, *p*_*θ*_ = *y*, and *p*_*eQTL,window adj*_ = *z*, we would determine how many matches satisfying |*θ*| ≥ *x*, *p*_*θ*_ ≤ *y*, and *p*_*eQTL,window adj*_ ≤ *z*, were obtained with the real data (= *N*_*R*_) and how many with the permuted data (= *N*_*P*_). The FDR of the corresponding DAP-EAP match was then computed 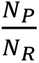. We defined two FDR thresholds of significance: Tier 1 (*FDR* ≤ 0.05) and Tier 2 (0.05 < *FDR* ≤ 0.10).

#### Differential expression analysis between active CD patients and controls

We collected blood from 55 active CD patients and performed RNA-Seq on the 27 fractionated circulating immune cell populations. Differential expression analysis between controls and patients was performed by cell type using the DESeq2 R package [Love *et al*., 2014], with the apeglm method for effect size shrinkage [Zhu *et al*., 2019]. Genes with a fold change above 1.5 and an adjusted p-value below 0.05 were considered differentially expressed (STable 26).

#### Comparing the number of DAP-EAP matches with the CEDAR2 cell-type specific information from ≤ 200 individuals versus the eQTLGen PBMC information from 35 K individuals

Summary statistics from eQTLGen, a meta-analysis of *cis*-eQTL results from 37 studies of blood and PBMC samples totaling 35K individuals, were downloaded (Võsa *et al*., 2021). We lifted their data over from GRCh37 to GRCh38. When arithmetic underflow was observed for the *p*-values, the log10 *p*-values were predicted from the z-scores using a local polynomial regression. We compared their 871 EAP (significant or not) with the 455 DAPs if the top variants were located within the disease interval. However, it was not possible to recompute the missing part of the EAP nor to compute a *p*-value for theta. Colocalized EAP correspond to EAP that have a |*θ*| ≥ 0.6 with a DAP (hence more permissive than the analysis of the CEDAR2 dataset).

## Acknowledgments

This project was conducted with funding from the *SYSCID* H2020 grant (ref. 733100, the MyQuant (ref. 30770923) and *BRIDGE* (O.0006.22 – RG3124) projects from the Excellence of Science (EOS) program (FNRS, Fédération Wallonie-Bruxelles and FWO, Flemish Community), the *CLIMAX* (WELBIO-CR-2022 A) project from WELBIO (Walloon Region), the *RHEAQT* (T.0096.19) and *IBD-GI-Seq* (T.0190.19) projects from the FNRS (Fédération Wallonie-Bruxelles), the ARC *RHEACT WITH HSPC* project from the University of Liège. Computational resources have been provided by the Consortium des Équipements de Calcul Intensif (CÉCI), funded by the Fonds de la Recherche Scientifique de Belgique (F.R.S.-FNRS) under Grant No. 2.5020.11 and by the Walloon Region. We thank Sandra Ormenese, Celine Vanwinge and the GIGA-Flow cytometry core facility for their support, as well as Naima Ahariz, Azeddine Bentaib and the other members of the GIGA genomics platform. Souad Rahmouni is a senior research associate of the FRS-FNRS. We are grateful to Yurii Aulchenko for many stimulating discussions.

## Members of the SYSCID Consortium

Konrad Aden, Vibeke Andersen, Diana Avalos, Aggelos Banos, George Bertsias, Marc Beyer, Johanna I Blase, Dimitrios Boumpas, Emmanouil T Dermitzakis, Axel Finckh, Andre Franke, Gilles Gasparoni, Michel Georges, Wei Gu, Robert Häsler, Mohamad Jawhara, Amy Kenyon, Christina Kratsch, Roland Krause, Gordan Lauc, Paul A Lyons, Massimo Mangino, Neha Mishra, Gioacchino Natoli, Marek Ostaszewski, Silja H Overgaard, Marija Pezer, Jeroen Raes, Souad Rahmouni, Benedikt Reiz, Elisa Rosati, Philip Rosenstiel, Despina Sanoudou, Venkata Satagopam, Reinhard Schneider, Jonas Schulte-Schrepping, Joachim L Schultze, Prodromos Sidiropoulos, Kenneth GC Smith, Signe B Sørensen, Timothy Spector, Doris Vandeputte, Sara Vieira-Silva, Aleksandar Vojta, Jörn Walter, Stefanie Warnat-Herresthal, and Vlatka Zoldoš.

## Members of the BRIDGE Consortium

Inna Afonina, Rudi Beyaert, Laure Dumoutier, Denis Franchimont, Michel Georges, Claude Libert, Claire Liefferinckx, Natalia Ferreras Moreno, Souad Rahmouni, Ramnik Xavier.

