## Supplementary figures and material for "Integrated cell type-specific analysis of blood and gut identifies matching eQTL for 140 IBD risk loci and entrectinib as possible repurposing candidate"

**for**

1. Unit of Animal Genomics, GIGA Institute & Faculty of Veterinary Medicine, University of Liège, Belgium. 2. Department of Gastroenterology, Faculty of Medicine, CHU & GIGA Institute, University of Liège, Belgium. 3. Genomics core facility, GIGA Institute, University of Liège, Belgium. 4. GIGA bioinformatics core facility, GIGA Institute, University of Liège, Belgium. 5. GIGA in vitro imaging and cell sorting core facility, GIGA Institute, University of Liège, Belgium. 6. Genomics core facility, University of Geneva, Switzerland. 7. Welbio Research Institute, Belgium.

### Supplemental Figure 1: Cell sorting and QC.

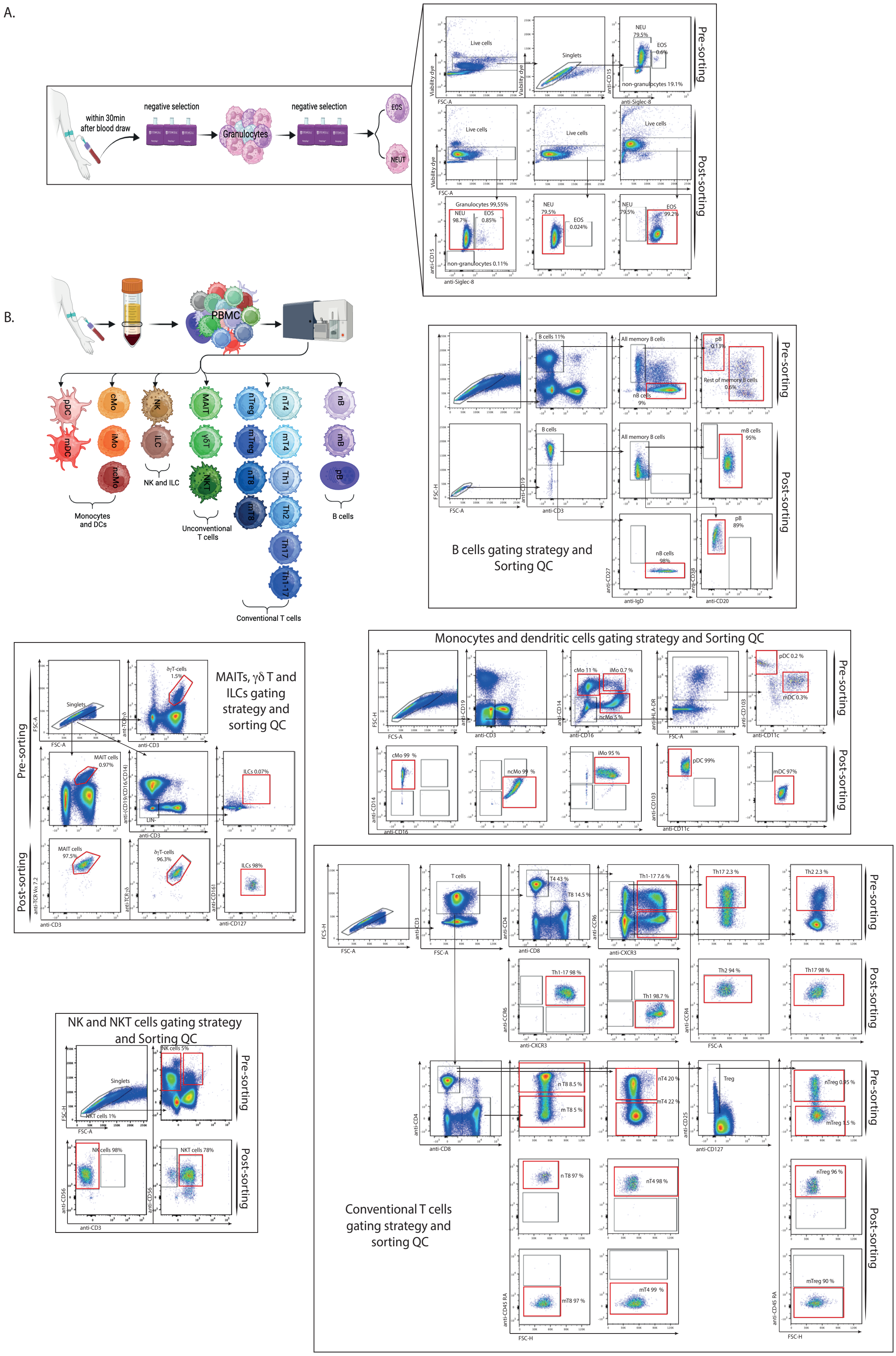

**(A)** Granulocytes magnetic cell sorting from EDTA-anticoagulated peripheral whole blood. Blood was processed for granulocytes negative cell sorting within 30 minutes after collection. ¼ and ½ of the sorted granulocytes were further processed separately and respectively for Neutrophils and Eosinophils selection. The remaining ¼ was immediately frozen until use. For QC of the cell sorting, an aliquot before and after each sorted fraction were stained using anti-CD15 and anti-Siglec-8 antibodies, together with viability dye. Left panel shows the percentage of each sorted population before and after sorting. **(B)** Flow cytometry cell sorting and QC of PBMC derived cell populations. Representative FACS plots illustrating gating strategies and QC after cell sorting for B cells (upper right panel), MAITs, gd T and ILC cells (middle left panel), monocytes and dendritic cells (middle right panel), NK and NKT cells (lower left panel), and conventional CD8 and CD4 T cells (lower right panel). For a detailed description of subset names, see STable 2. For information on the flow-cytometry antibodies used in this study, see STable 29. Red quadrants indicate the target sorted cell types.

### Supplemental Figure 2: Quality control of the bulk RNA-Seq data on 27 circulating immune cell populations.

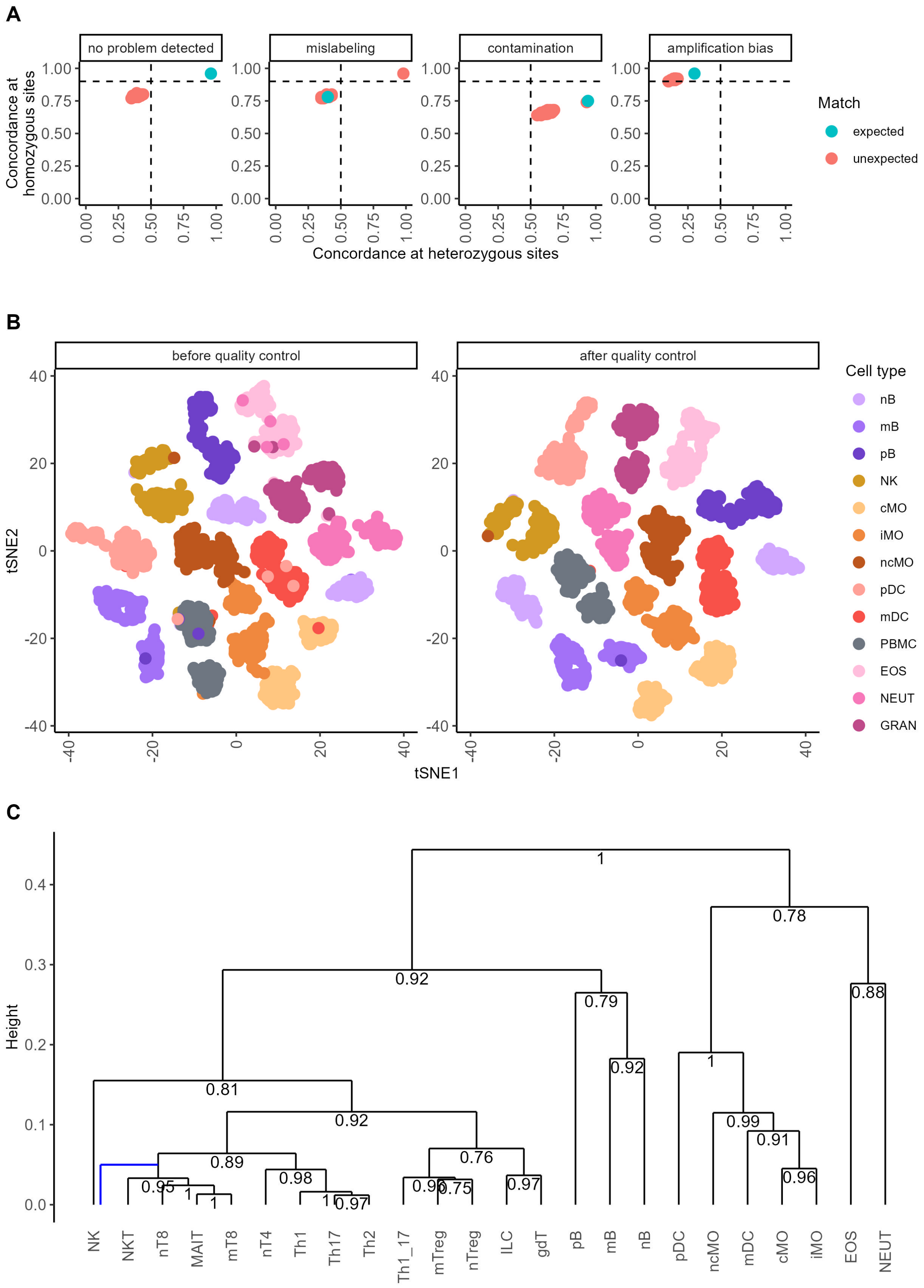

**(A)** Examples of issues detected by confronting genome and transcriptome genotypes. From left to right: (i) Example of an RNA sample that matches its presumed donor (green dot); the red dots correspond to the confrontation of that RNA sample with the DNA of all other individuals. (ii) Example of an RNA sample that matches the DNA of an individual that is not the presumed donor. (iii) Example of an RNA sample that is likely contaminated (mixture between two samples), as it is "too heterozygous" and hence shows a drop in concordance at homozygous genotypes for both the presumed donor (green dot) and the contaminating individual (co-positioned red dot). (iv) Example of a likely PCR issue; the RNA appears "too homozygous" and hence shows a drop in concordance at heterozygous genotypes. **(B)** Example of issues detected when confronting presumed cell type vs inferred transcriptome-based cell type: tSNA map of 2,444 samples from 13 cell types labeled by presumed cell type of origin before (left) and after (right) quality control. Based on the expression level of 500 most variable genes. **(C)** Hierarchical tree for 24 of the 27 cell types (PBMC, granulocytes and memory CD4 were not included because being mixtures of cell types) obtained from 1-|Spearman's correlation| between the mean (across all samples) expression levels of the 4,000 most variable (ANOVA F value of cell type effect) coding genes using "average" method. The numbers on the nodes correspond to the proportion of trees recapitulating the same split when varying the number of variable genes considered from 250 to 8,000 (32 analyses) and using the same method. The tree shown is the best supported tree in the method. Nearly identical (most supported) trees were obtained when applying the "ward.d" and "ward.d2" methods. The blue line marks the only differences that was observed with the "ward.d2" method.

### Supplemental Figure 3: eQTL mapping and construction of regulatory modules in 27 circulating blood cell populations.

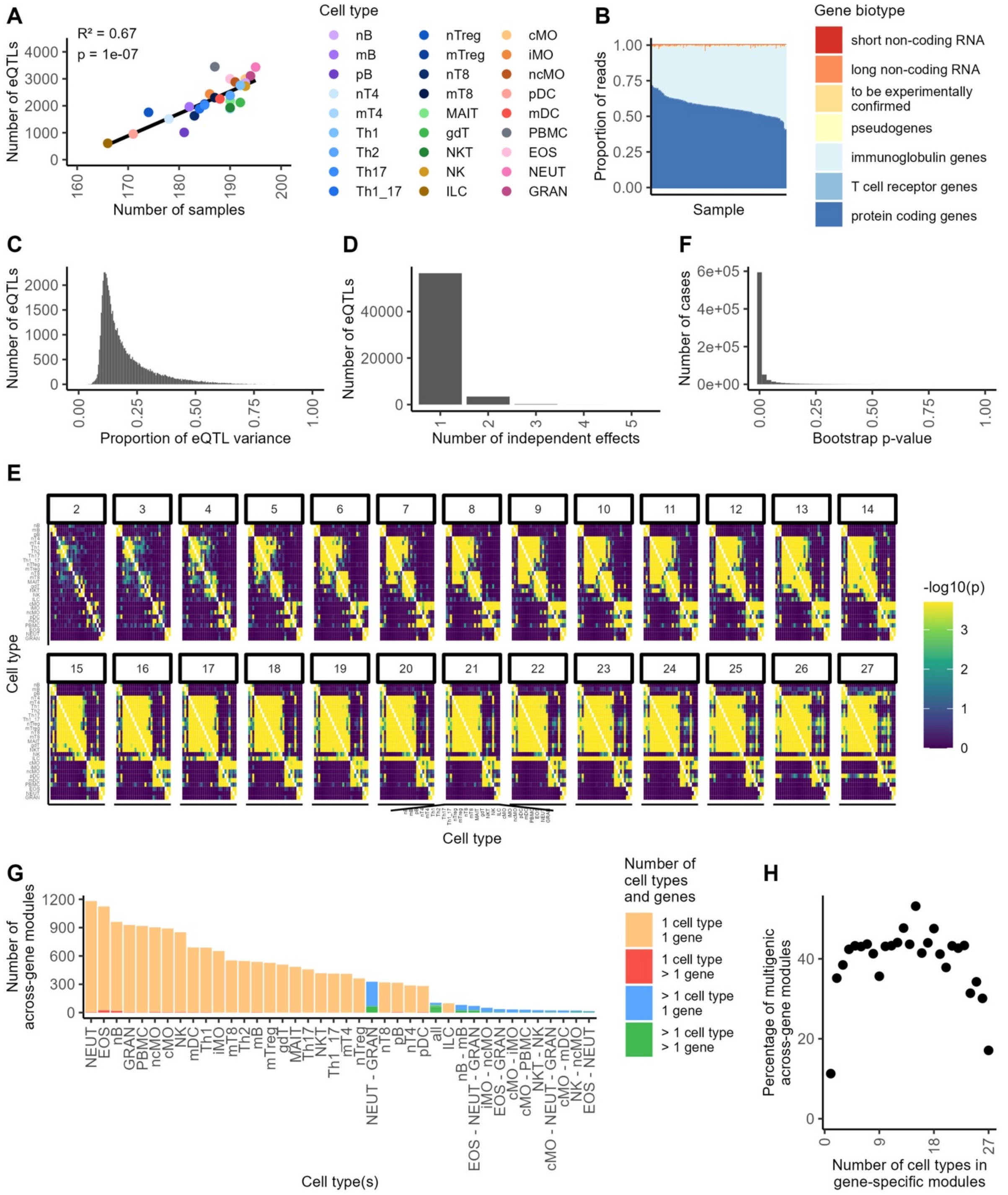

**(A)** Number of detected eQTL as a function of the number of usable samples for the corresponding (color labeled) cell type. **(B)** Demonstration of the inter-individual variation in the abundance of immunoglobulin reads (light blue) in plasmocytes of 181 individuals. **(C)** Distribution of the proportion of variation in gene expression level accounted for by the (primary) detected *cis*-eQTL. **(D)** Results of the conditional analysis for the detection of independent secondary *cis*-eQTL effects. **(E)** Log(1/p-value) of the excess number of shared modules over expectations (given number of detected eQTLs in the respective cell types) for all possible pairs of cell types. Computations are performed by including modules with two active cell types (upper left facet), three or less active cell types, four or less active cell types, …, 27 or less active cell types (lower right facet). **(F)** Frequency distribution of the p-values obtained using an empirical cell-type x eQTL effect "interaction" test. We determined the probability to obtain a *cis*-eQTL effect that is as low or lower than the one observed in cell type B, by sampling expression levels (bootstrapping) by variant genotype from cell type A. The large excess of low p-values is striking (uniform distribution expected un the null), yielding an estimate of $\pi_1$ [Storey & Tibshirani, 2003] of 99.9%. **(G)** Across-gene regulatory modules grouped by the combination of cell types in which they are active and ordered by the frequency of occurrence of the corresponding pattern. Modules encompassing one gene and active in one cell type only are labeled in yellow (n = 15,926). Modules encompassing more than one gene yet active in only one cell type are labeled in red (n = 84). Modules encompassing one gene yet active in more than one cell type are labeled in blue (n = 3,433). Modules encompassing more than one gene and active in more than one cell type are labeled in green (n = 1,451). Only the 40 more common activity patterns (out of 2,959 in total) are shown. These include the 27 patterns corresponding to modules active in only one cell type. **(H)** Estimation of the proportion of gene-specific modules that merge with another gene-specific module to create a multigenic module when considering gene-specific modules active in 1, 2, 3, … 27 cell types (x-axis). Interestingly, gene-specific modules active in one cell type rarely (11.3%) fuse with others to form multigenic modules. Likewise, gene-specific modules that are active in all 27 cell types rarely (17.1%) become multigenic modules. A contrario, gene-specific modules that are active in 2 to 26 cell type more often (average 41.4%; range: 30.1% - 53.2%) fuse with others to form multigenic modules. To make sure that the probability of merging between gene-specific modules was not affected by the number of EAPs in the modules, we exclusively used one EAP per gene-specific module, namely the consensus EAP for modules with more than one EAP.

### Supplemental Figure 4: Screenshots from the CEDAR website ([https://tools.giga.uliege.be/cedar](https://tools.giga.uliege.be/cedar)) for the 27 circulating immune cell populations.

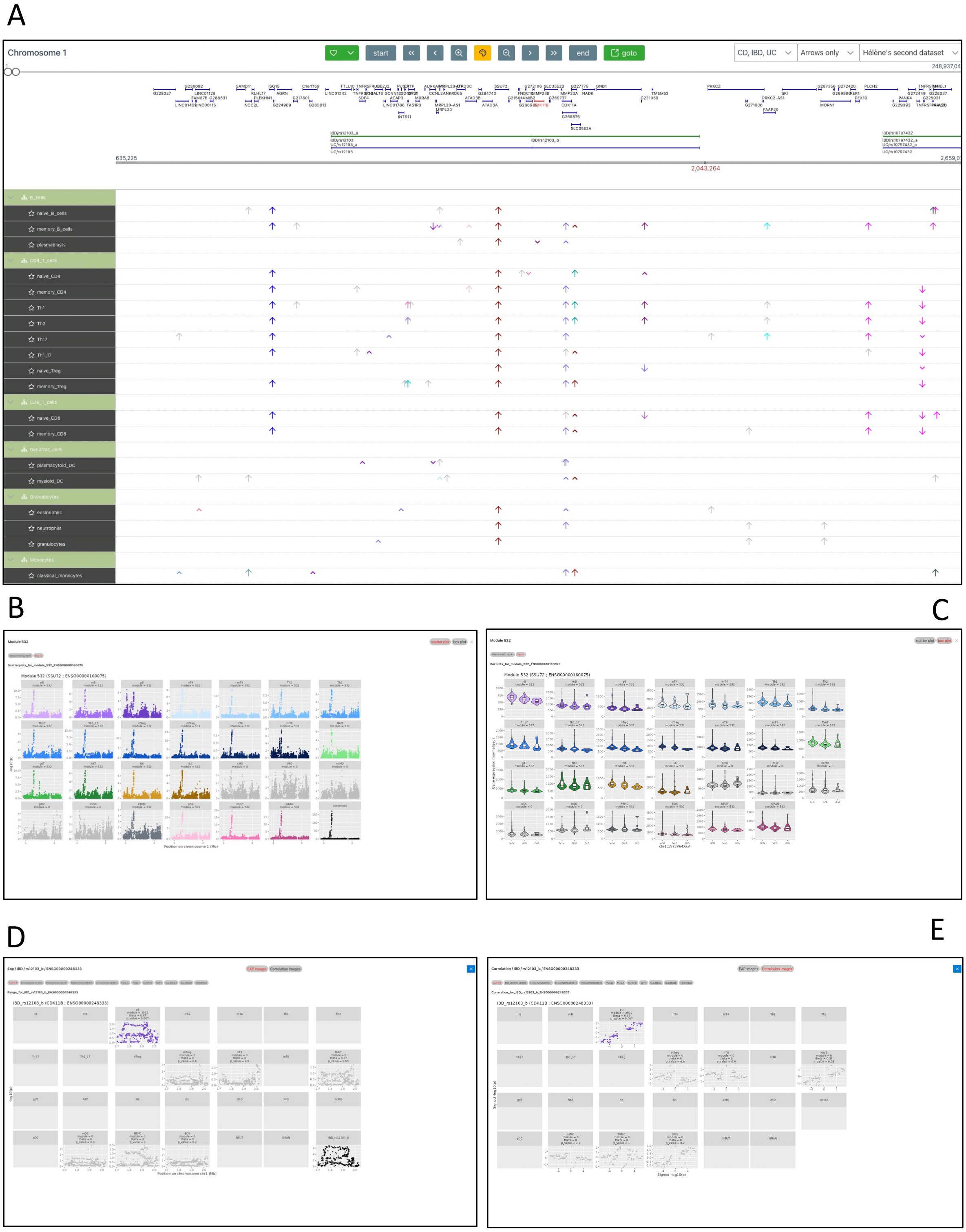

**(A)** Top panel: genome browser showing the position of genes and risk loci for inflammatory bowel disease (IBD). Bottom panel: activity of the eQTL and *cis*-acting regulatory modules (RM). Significant eQTL are marked by large arrows (↑). Arrows for eQTL that are part of the same module have the same color. eQTL in a module with effect sign opposite to the most significant eQTL in the module ("representative") are marked by a downward pointing arrow (↓). Non-significant eQTL that are part of a module by virtue of matching EAP ($|\theta| \geq 0.6$) are marked by small arrows (∧ or ∨). eQTL that are alone in their own module are in grey. Modules can be highlighted by clicking on them. **(B)** By clicking on a module, one has access to the EAP graphs showing the activity of the corresponding eQTL in all 27 cell types by gene (one page per gene). EAPs that are part of the selected module are colored, others are shown in gray. If the gene was not expressed in a given cell type (and the eQTL could not be tested) the EAP graph is "empty". The last facet (lower right) corresponds to the "consensus" EAP of the module. **(C)** By clicking on a module, one also has access to violin plots at the lead SNP of the consensus EAP showing the distribution of DESeq2 normalized gene expression values for individuals sorted by SNP genotype. **(D)** By clicking on the bars corresponding to the disease risk loci, one has access to the EAP in all 27 cell types of all genes (one page per gene) that have a matching DAP-EAP ($|\theta| \geq 0.6$, $p \leq 0.05$) with the corresponding DAP (lower right facet). **(E)** By clicking on the bars corresponding to the disease risk loci, one also has access to the theta-plots [Momozawa *et al.* 2018] in all 27 cell types of all genes (one page per gene) that have a matching DAP-EAP ($|\theta| \geq 0.6$, $p \leq 0.05$).

### Supplemental Figure 5: Graphical abstract of workflow of *cis*-eQTL analyses in scRNA-Seq data from intestinal biopsies.

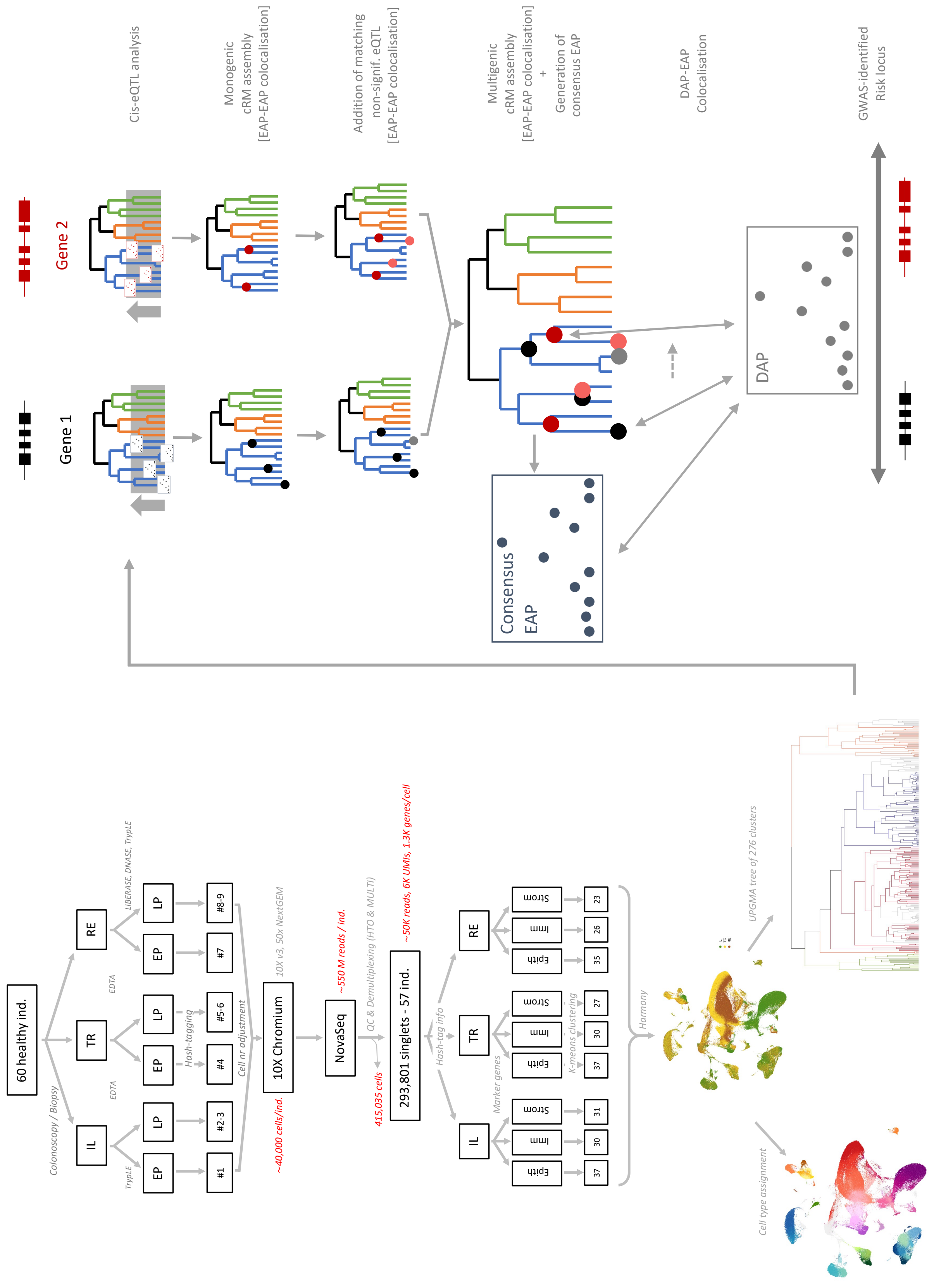

### Supplemental Figure 6: Cell type assignment of CEDAR2 intestinal scRNA-Seq data.

A 10 grand clusters

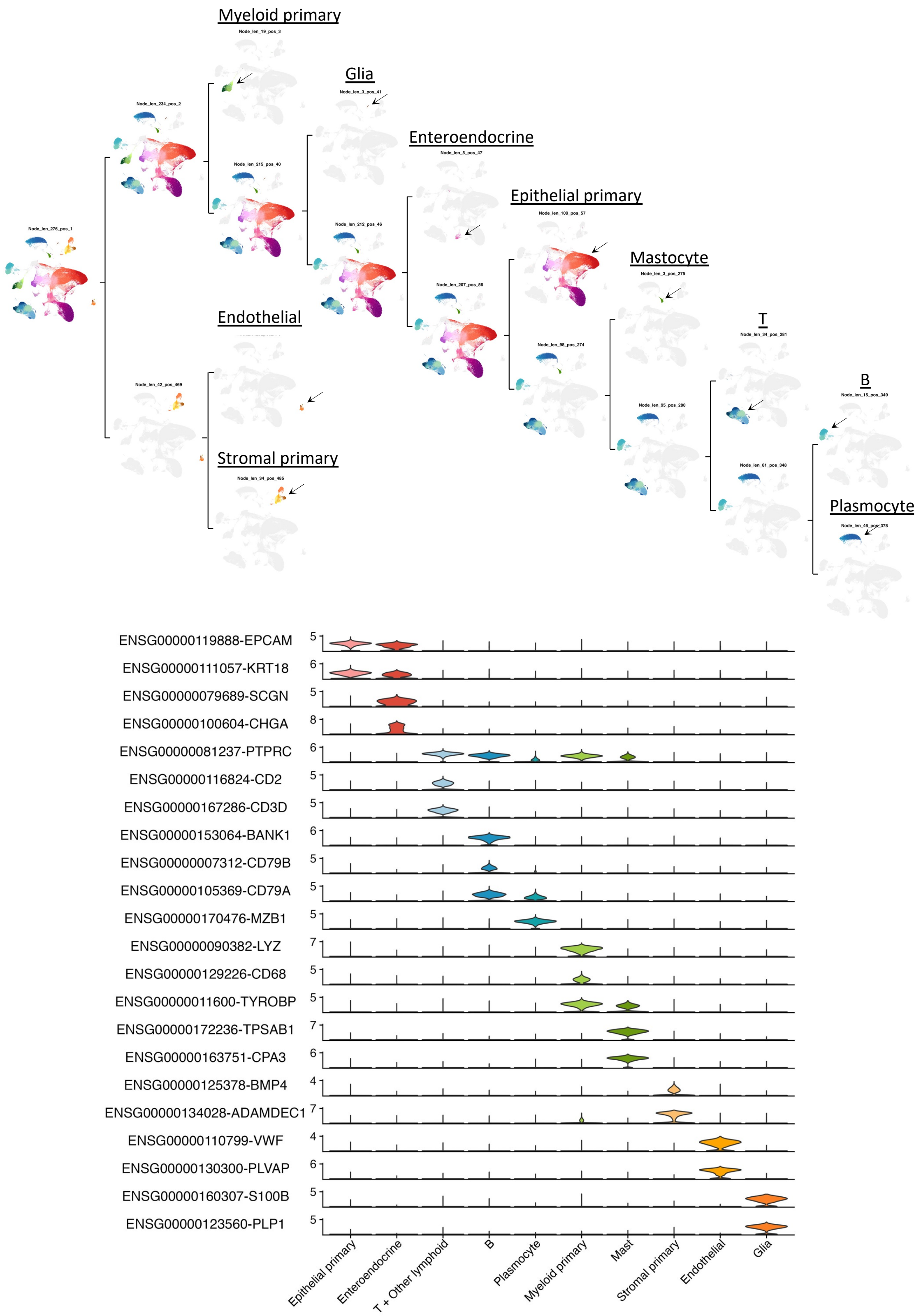

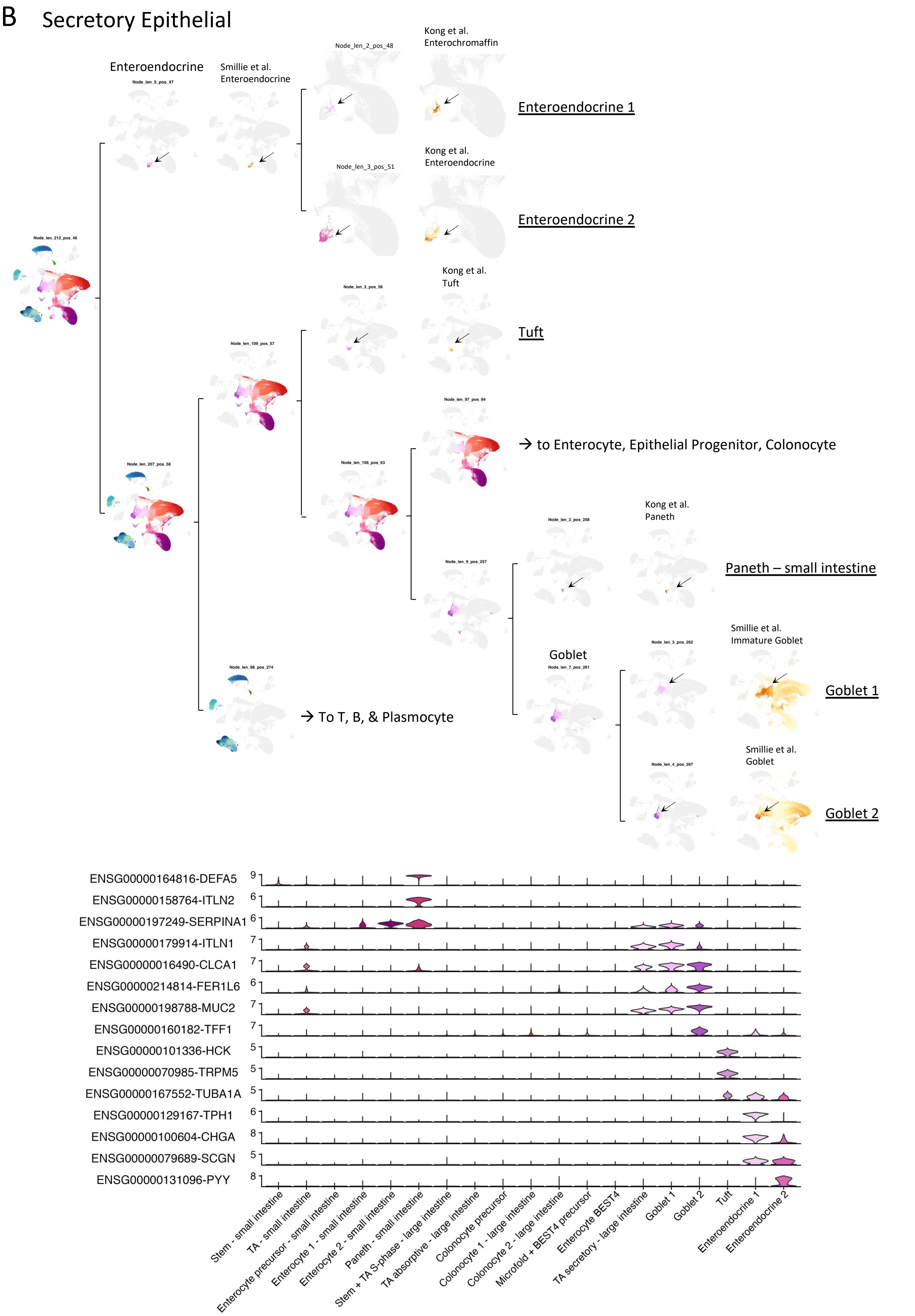

B
Secretory Epithelial
Enteroendocrine
Smillie et al.
Enteroendocrine
Kong et al.
Enterochromaffin
Enteroendocrine 1
Kong et al.
Enteroendocrine
Enteroendocrine 2
Kong et al.
Tuft
Tuft
→ to Enterocyte, Epithelial Progenitor, Colonocyte
Kong et al.
Paneth
Paneth – small intestine
Goblet
Smillie et al.
Immature Goblet
Goblet 1
→ To T, B, & Plasmocyte
Smillie et al.
Goblet
Goblet 2
ENSG00000164816-DEFA5
ENSG00000158764-ITLN2
ENSG00000197249-SERPINA1
ENSG00000179914-ITLN1
ENSG00000016490-CLCA1
ENSG00000214814-FER1L6
ENSG00000198788-MUC2
ENSG00000160182-TFF1
ENSG00000101336-HCK
ENSG00000070985-TRPM5
ENSG00000167552-TUBA1A
ENSG00000129167-TPH1
ENSG00000100604-CHGA
ENSG00000079689-SCGN
ENSG00000131096-PYY
Stem - small intestine
TA - small intestine
Enterocyte precursor - small intestine
Enterocyte 1 - small intestine
Enterocyte 2 - small intestine
Paneth - small intestine
Stem + TA S-phase - large intestine
TA absorptive - large intestine
Colonocyte precursor
Colonocyte 1 - large intestine
Colonocyte 2 - large intestine
Microfold + BEST4 precursor
Enterocyte BEST4
TA secretory - large intestine
Goblet 1
Goblet 2
Tuft
Enteroendocrine 1
Enteroendocrine 2

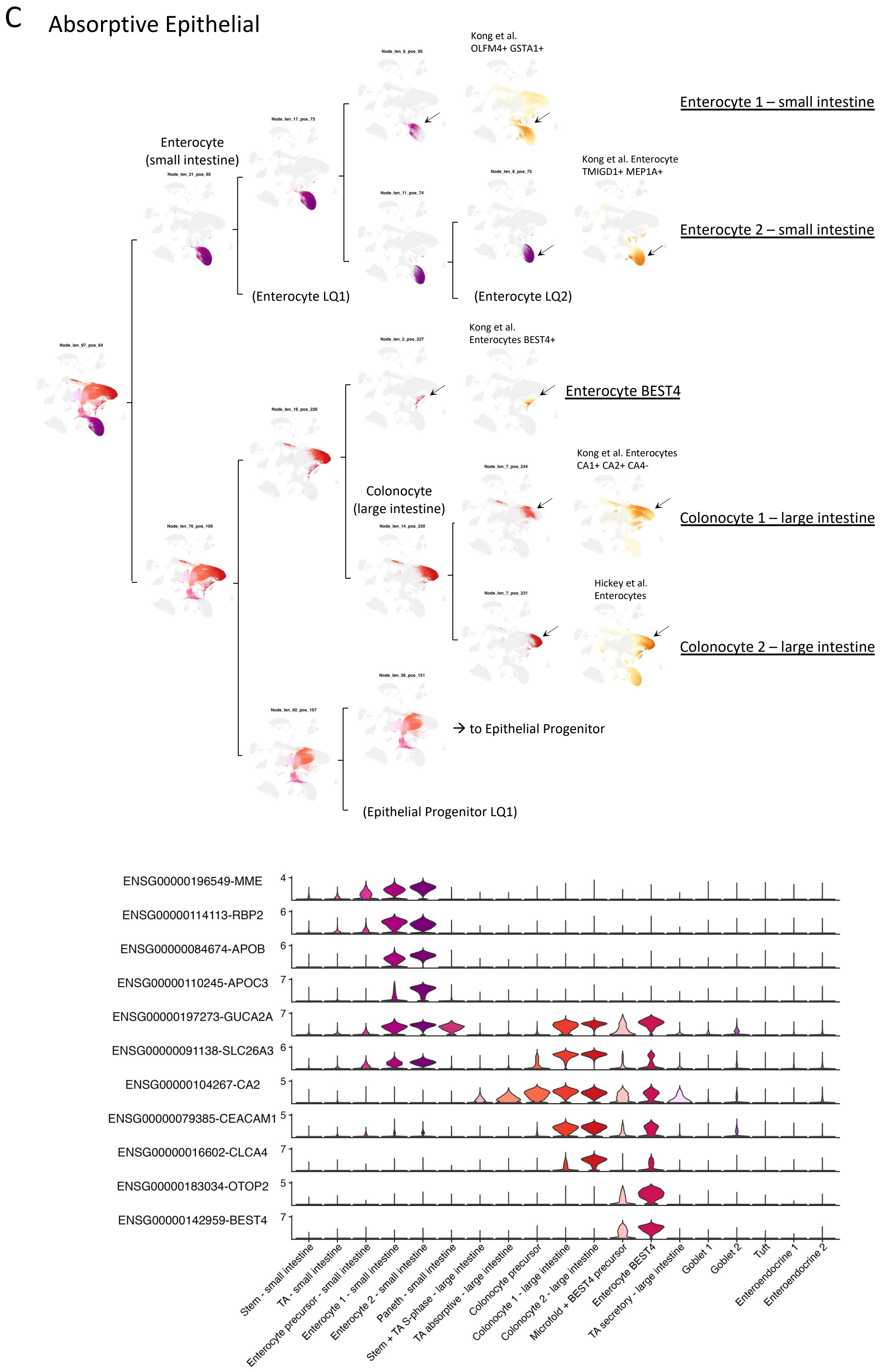
C
Absorptive Epithelial
Enterocyte (small intestine)
Kong et al. OLFM4+ GSTA1+
Enterocyte 1 – small intestine
Kong et al. Enterocyte TMIGD1+ MEP1A+
Enterocyte 2 – small intestine
(Enterocyte LQ1)
(Enterocyte LQ2)
Kong et al. Enterocytes BEST4+
Enterocyte BEST4
Colonocyte (large intestine)
Kong et al. Enterocytes CA1+ CA2+ CA4-
Colonocyte 1 – large intestine
Hickey et al. Enterocytes
Colonocyte 2 – large intestine
→ to Epithelial Progenitor
(Epithelial Progenitor LQ1)
ENSG00000196549-MME
ENSG00000114113-RBP2
ENSG00000084674-APOB
ENSG00000110245-APOC3
ENSG00000197273-GUCA2A
ENSG00000091138-SLC26A3
ENSG00000104267-CA2
ENSG00000079385-CEACAM1
ENSG00000016602-CLCA4
ENSG00000183034-OTOP2
ENSG00000142959-BEST4
Stem - small intestine
TA - small intestine
Enterocyte precursor - small intestine
Enterocyte 1 - small intestine
Enterocyte 2 - small intestine
Paneth - small intestine
Stem + TA S-phase - large intestine
TA absorptive - large intestine
Colonocyte precursor
Colonocyte 1 - large intestine
Colonocyte 2 - large intestine
Microfold + BEST4 precursor
Enterocyte BEST4
TA secretory - large intestine
Goblet 1
Goblet 2
Tuft
Enteroendocrine 1
Enteroendocrine 2

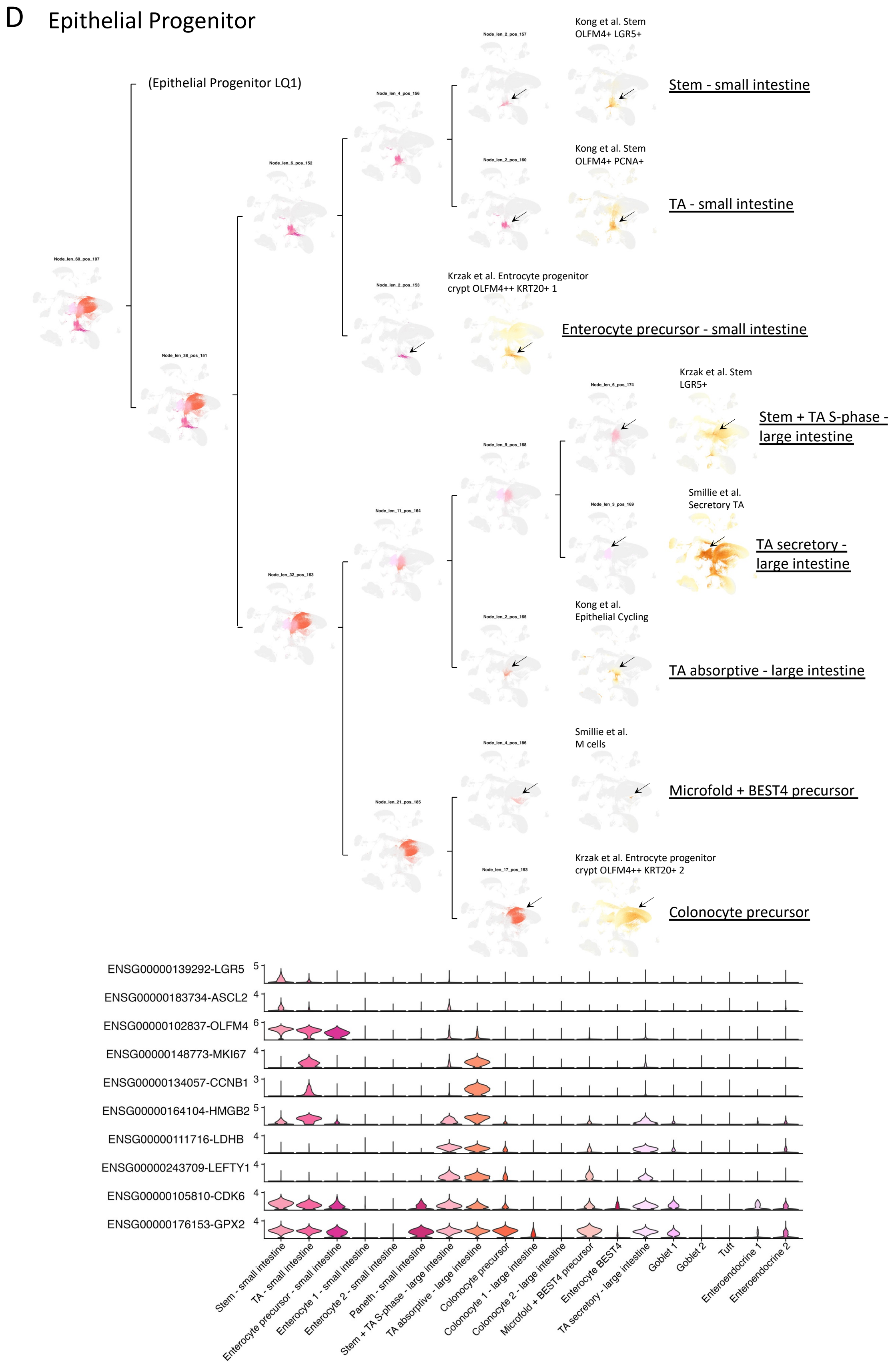
D
Epithelial Progenitor
(Epithelial Progenitor LQ1)
Kong et al. Stem
OLFM4+ LGR5+
Stem - small intestine
Kong et al. Stem
OLFM4+ PCNA+
TA - small intestine
Krzak et al. Entrocyte progenitor
crypt OLFM4++ KRT20+ 1
Enterocyte precursor - small intestine
Krzak et al. Stem
LGR5+
Stem + TA S-phase - large intestine
Smillie et al.
Secretory TA
TA secretory - large intestine
Kong et al.
Epithelial Cycling
TA absorptive - large intestine
Smillie et al.
M cells
Microfold + BEST4 precursor
Krzak et al. Entrocyte progenitor
crypt OLFM4++ KRT20+ 2
Colonocyte precursor
ENSG00000139292-LGR5
ENSG00000183734-ASCL2
ENSG00000102837-OLFM4
ENSG00000148773-MKI67
ENSG00000134057-CCNB1
ENSG00000164104-HMGB2
ENSG00000111716-LDHB
ENSG00000243709-LEFTY1
ENSG00000105810-CDK6
ENSG00000176153-GPX2
Stem - small intestine
TA - small intestine
Enterocyte precursor - small intestine
Enterocyte 1 - small intestine
Enterocyte 2 - small intestine
Paneth - small intestine
Stem + TA S-phase - large intestine
TA absorptive - large intestine
Colonocyte precursor
Colonocyte 1 - large intestine
Colonocyte 2 - large intestine
Microfold + BEST4 precursor
Enterocyte BEST4
TA secretory - large intestine
Goblet 1
Goblet 2
Tuft
Enteroendocrine 1
Enteroendocrine 2

E Myeloid primary

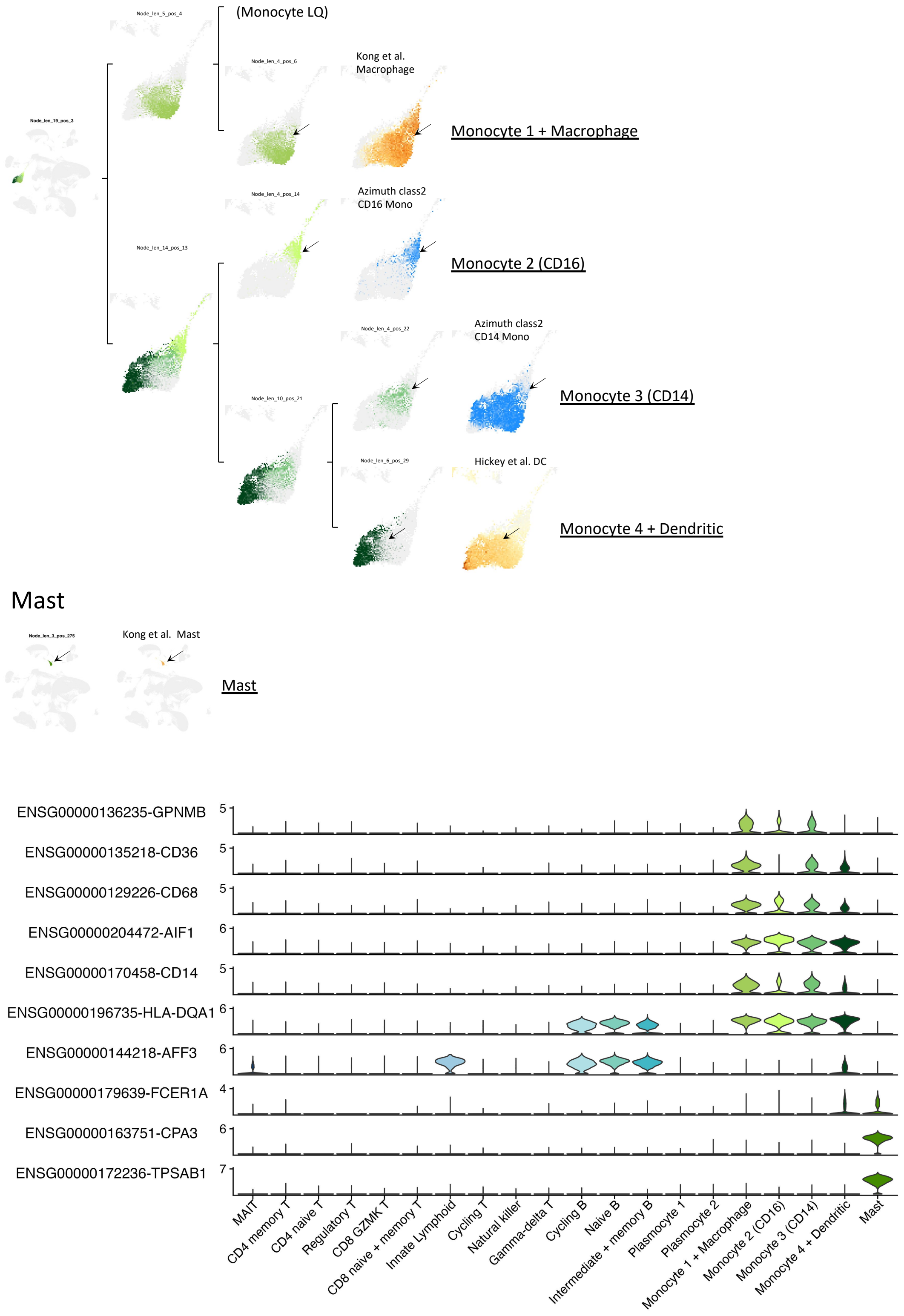

F T + Other lymphoid

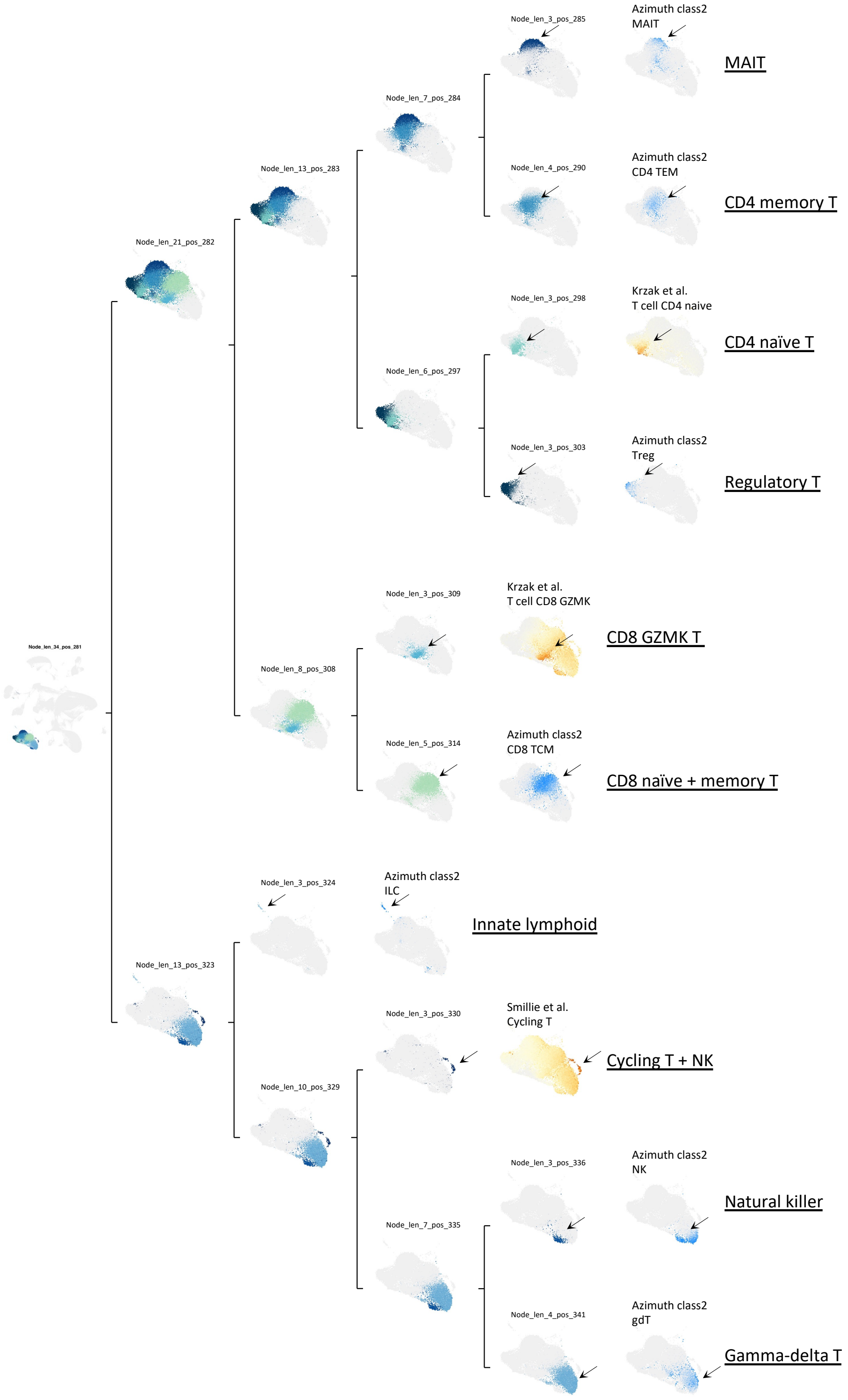

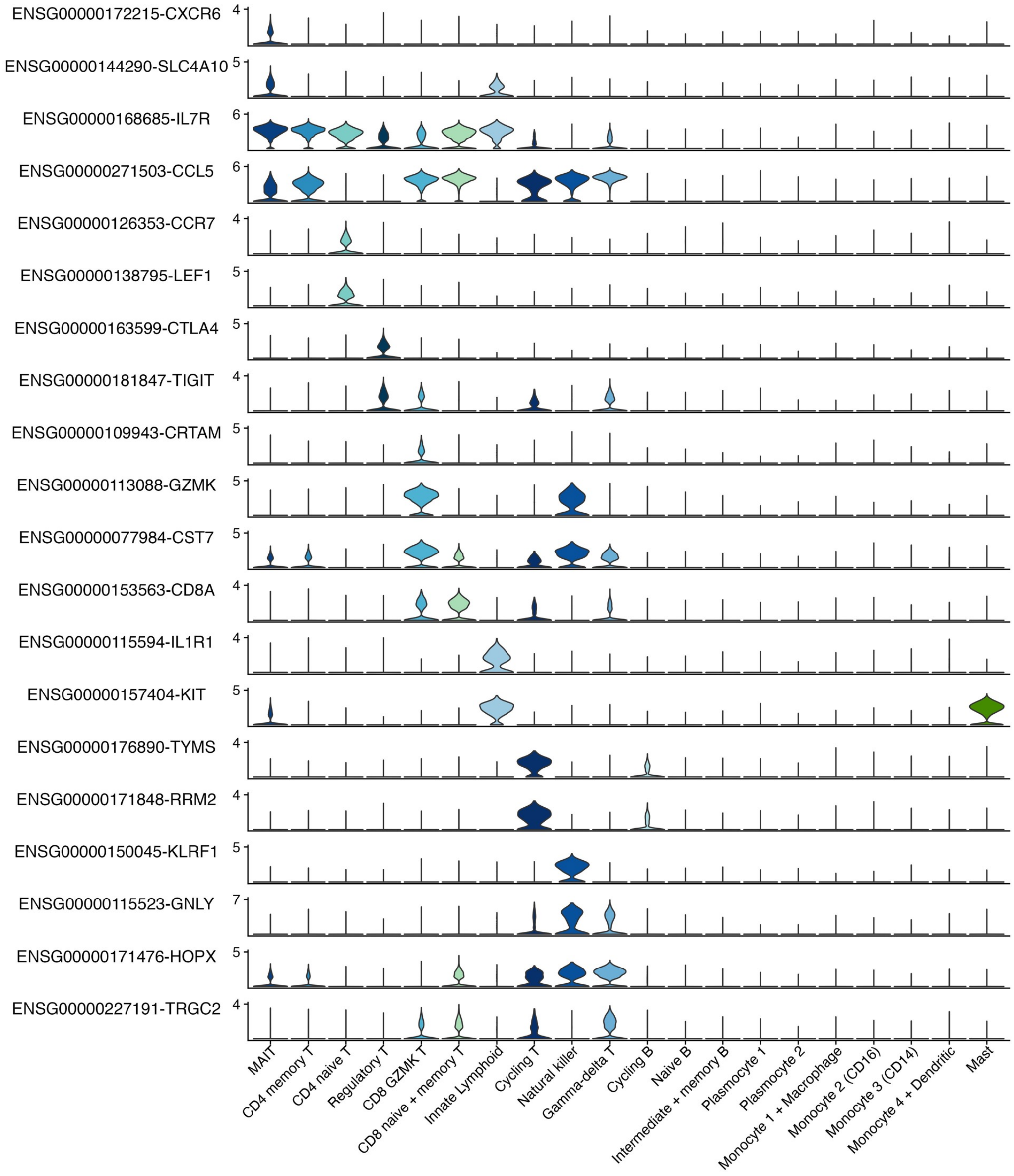
ENSG00000172215-CXCR6
ENSG00000144290-SLC4A10
ENSG00000168685-IL7R
ENSG00000271503-CCL5
ENSG00000126353-CCR7
ENSG00000138795-LEF1
ENSG00000163599-CTLA4
ENSG00000181847-TIGIT
ENSG00000109943-CRTAM
ENSG00000113088-GZMK
ENSG00000077984-CST7
ENSG00000153563-CD8A
ENSG00000115594-IL1R1
ENSG00000157404-KIT
ENSG00000176890-TYMS
ENSG00000171848-RRM2
ENSG00000150045-KLRF1
ENSG00000115523-GNLY
ENSG00000171476-HOPX
ENSG00000227191-TRGC2
MAIT
CD4 memory T
CD4 naive T
Regulatory T
CD8 GZMK T
CD8 naive + memory T
Innate Lymphoid
Cycling T
Natural killer
Gamma-delta T
Cycling B
Naive B
Intermediate + memory B
Plasmocyte 1
Plasmocyte 2
Monocyte 1 + Macrophage
Monocyte 2 (CD16)
Monocyte 3 (CD14)
Monocyte 4 + Dendritic
Mast

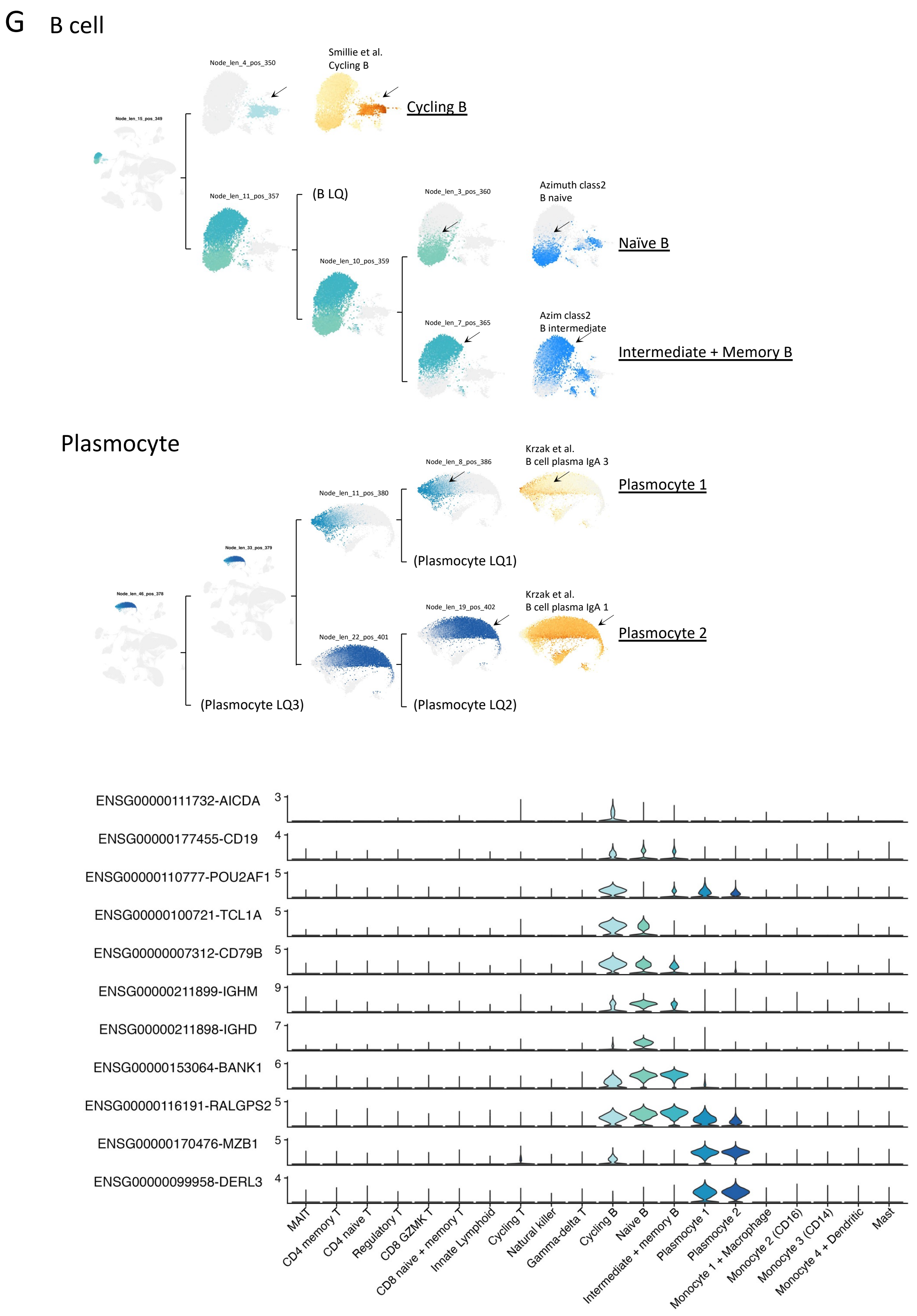
G
B cell
Node_len_4_pos_350
Smillie et al.
Cycling B
Cycling B
Node_len_11_pos_357
(B LQ)
Node_len_3_pos_360
Azimuth class2
B naive
Naïve B
Node_len_10_pos_359
Node_len_7_pos_365
Azim class2
B intermediate
Intermediate + Memory B
Plasmocyte
Node_len_8_pos_386
Krzak et al.
B cell plasma IgA 3
Plasmocyte 1
Node_len_11_pos_380
(Plasmocyte LQ1)
Krzak et al.
B cell plasma IgA 1
Node_len_19_pos_402
Plasmocyte 2
Node_len_22_pos_401
(Plasmocyte LQ3)
(Plasmocyte LQ2)
ENSG00000111732-AICDA
ENSG00000177455-CD19
ENSG00000110777-POU2AF1
ENSG00000100721-TCL1A
ENSG00000007312-CD79B
ENSG00000211899-IGHM
ENSG00000211898-IGHD
ENSG00000153064-BANK1
ENSG00000116191-RALGPS2
ENSG00000170476-MZB1
ENSG00000099958-DERL3
MAIT
CD4 memory T
CD4 naive T
Regulatory T
CD8 GZMK T
CD8 naive + memory T
Innate Lymphoid
Cycling T
Natural killer
Gamma-delta T
Cycling B
Naive B
Intermediate + memory B
Plasmocyte 1
Plasmocyte 2
Monocyte 1 + Macrophage
Monocyte 2 (CD16)
Monocyte 3 (CD14)
Monocyte 4 + Dendritic
Mast

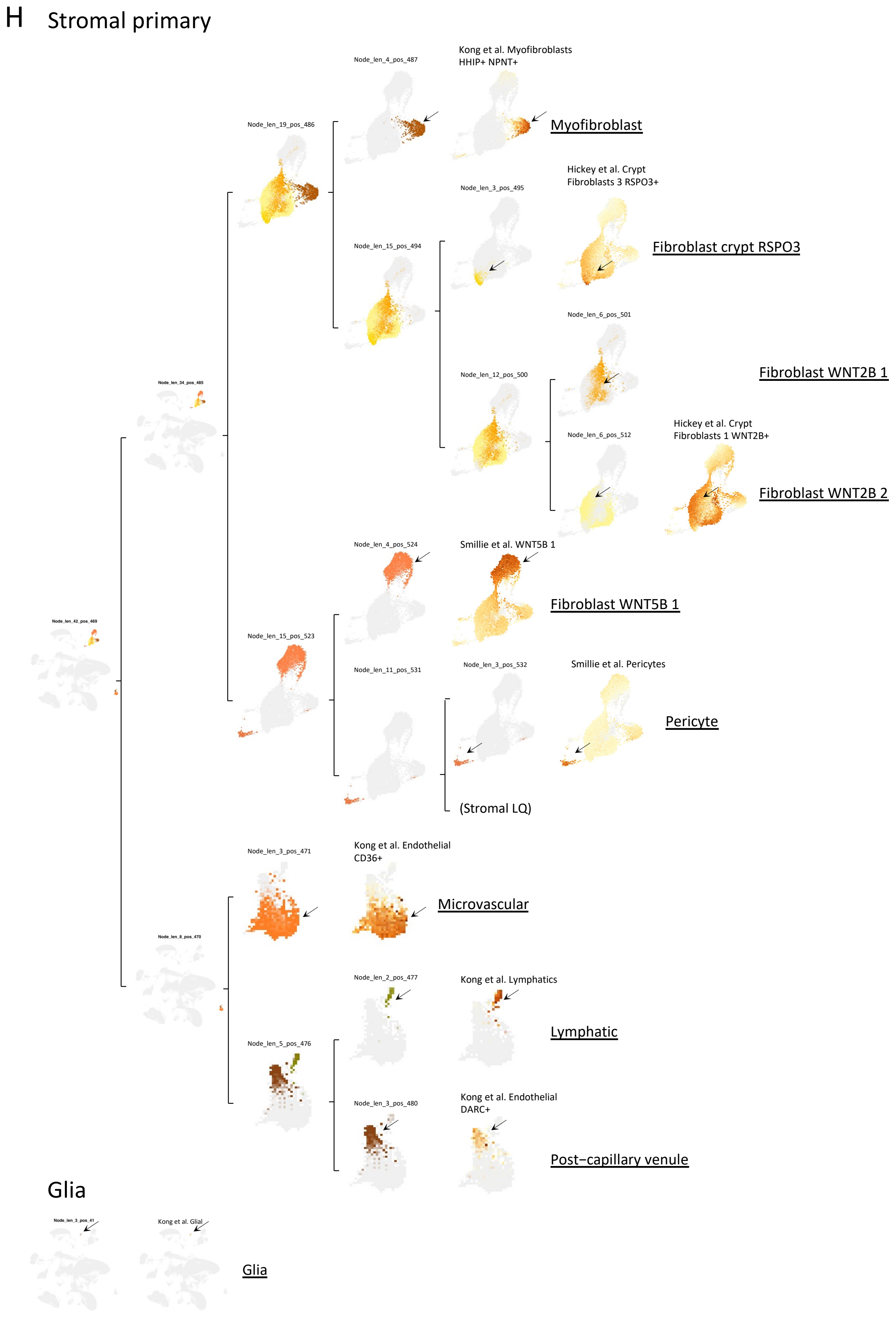
H
Stromal primary
Node_len_42_pos_469
Node_len_34_pos_485
Node_len_19_pos_486
Node_len_4_pos_487
Kong et al. Myofibroblasts HHIP+ NPNT+
Myofibroblast
Node_len_15_pos_494
Node_len_3_pos_495
Hickey et al. Crypt Fibroblasts 3 RSPO3+
Fibroblast crypt RSPO3
Node_len_12_pos_500
Node_len_6_pos_501
Fibroblast WNT2B 1
Node_len_6_pos_512
Hickey et al. Crypt Fibroblasts 1 WNT2B+
Fibroblast WNT2B 2
Node_len_15_pos_523
Node_len_4_pos_524
Smillie et al. WNT5B 1
Fibroblast WNT5B 1
Node_len_11_pos_531
Node_len_3_pos_532
Smillie et al. Pericytes
Pericyte
(Stromal LQ)
Node_len_8_pos_470
Node_len_3_pos_471
Kong et al. Endothelial CD36+
Microvascular
Node_len_5_pos_476
Node_len_2_pos_477
Kong et al. Lymphatics
Lymphatic
Node_len_3_pos_480
Kong et al. Endothelial DARC+
Post–capillary venule
Glia
Node_len_3_pos_41
Kong et al. Glial
Glia

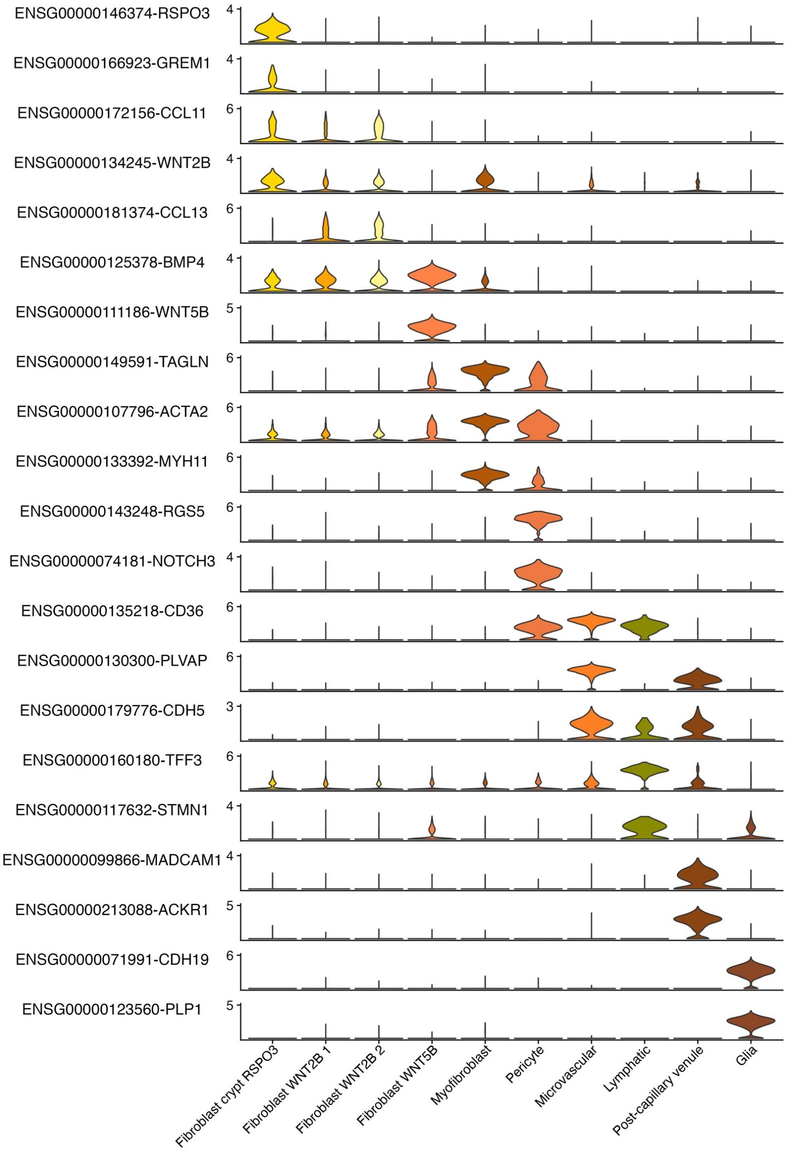
ENSG00000146374-RSPO3
ENSG00000166923-GREM1
ENSG00000172156-CCL11
ENSG00000134245-WNT2B
ENSG00000181374-CCL13
ENSG00000125378-BMP4
ENSG00000111186-WNT5B
ENSG00000149591-TAGLN
ENSG00000107796-ACTA2
ENSG00000133392-MYH11
ENSG00000143248-RGS5
ENSG00000074181-NOTCH3
ENSG00000135218-CD36
ENSG00000130300-PLVAP
ENSG00000179776-CDH5
ENSG00000160180-TFF3
ENSG00000117632-STMN1
ENSG00000099866-MADCAM1
ENSG00000213088-ACKR1
ENSG00000071991-CDH19
ENSG00000123560-PLP1
Fibroblast crypt RSPO3
Fibroblast WNT2B 1
Fibroblast WNT2B 2
Fibroblast WNT5B
Myofibroblast
Pericyte
Microvascular
Lymphatic
Post-capillary venule
Glia

I

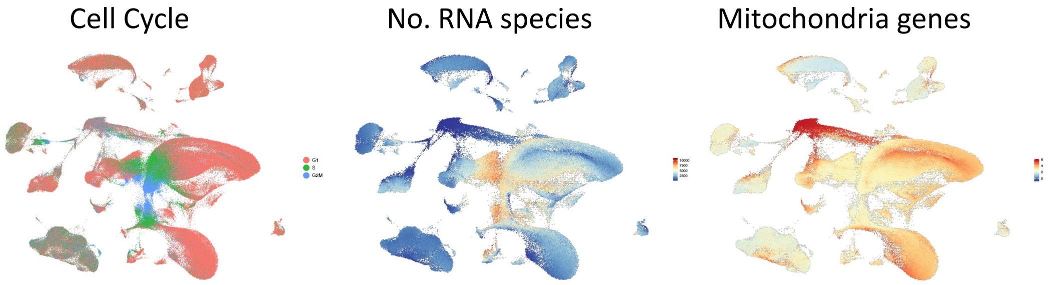
Cell Cycle
No. RNA species
Mitochondria genes

J

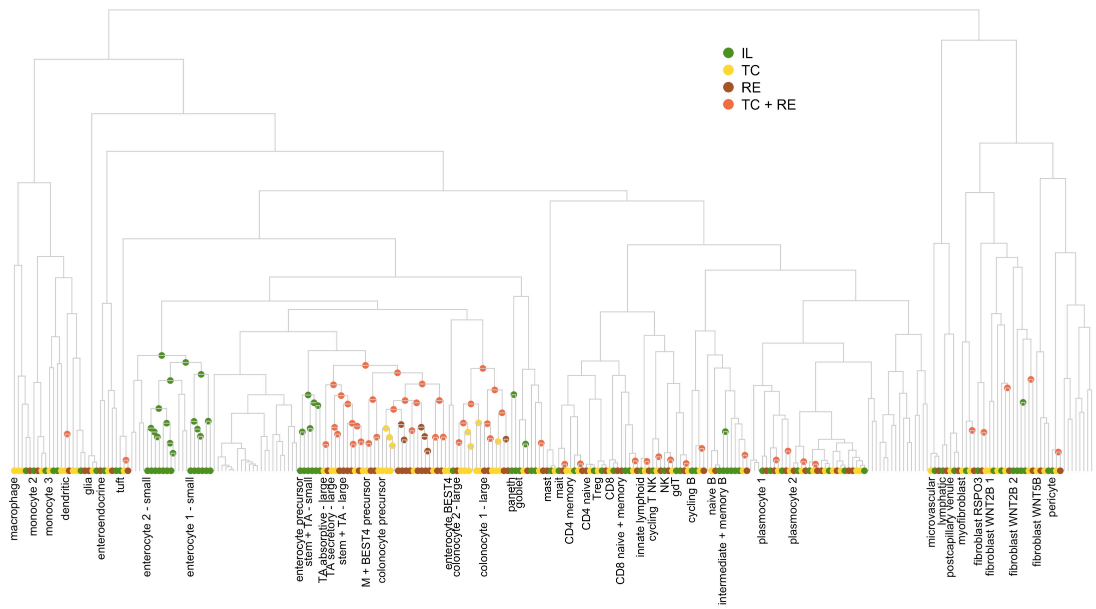

**(A-H)** Illustration of the cell content of the nodes and leaves of the hierarchical tree of cell clusters. The different panels sequentially walk the reader through the tree from the root towards the leaf. In terms of cellular content, each node in the tree corresponds to the agglomeration of the cells of all dependent leaves. For each node, we show the global UMAP yet color only the cells that are part of the node. The colors used correspond to the colors of Fig. 2D, with epithelial cells in reddish tones, lymphoid cells in bluish tones, myeloid cells in greenish tones, and stromal cells in orange tones. For part of the nodes, corresponding to what were, in the end, nodes identified as the upper levels of one of 49 specific cell types, we add the same UMAP yet this time labeled in pseudo-color measuring the intensity of expression of a set of signature genes. The source of the corresponding gene signature is provided, as well as the name of the corresponding cell type (underlined). If the source is one of the following references [Smillie *et al.* 2019; Hickey *et al.* 2023; Kong *et al.* 2023; Krzak *et al.* 2023], we use yellowish tones. If the source is the Azimuth human PBMC reference mapping program [Hao *et al.*, 2021], we use bluish tones. The reason why we herein mention 49 cell types as opposed to 43 in f.i. Fig. 2A, is because we herein (SFig. 6) separately highlight ileal, colonic and rectal cell clusters for some epithelial cell type, while these are considered as one in Fig. 2A. Each lower panel shows expressions of a subset of marker genes (rows) characteristic for the indicated cell types (columns) in references [Franzén *et al.* 2019; Smillie *et al.* 2019; Burclaff *et al.* 2022; Ishikawa *et al.* 2022; Hickey *et al.* 2023; Kong *et al.* 2023; Hao *et al.*, 2024].

**(I)** Seurat cell cycle phase scores, the number of RNA detected and module scores for mitochondrial genes are shown on the UMAP.

**(J)** Nodes, in the hierarchical tree, for which all dependent leaves derive from the same anatomical location (terminal ileum, transverse colon, rectum) are color-labeled accordingly. It appears that one has to go quite "high" (i.e., towards the leaves) in the tree, before colors appear. This means that for most cell types, the cells have very similar transcriptome across anatomical locations. This is especially true for myeloid and lymphoid cell types. Separation by anatomical location occurs a bit earlier in the tree for epithelial and, to a lesser extent, for stromal cells.

### Supplemental Figure 7: Screenshots from the CEDAR website (https://tools.giga.uliege.be/cedar) for the scRNA Seq analyses of the intestinal biopsies.

A

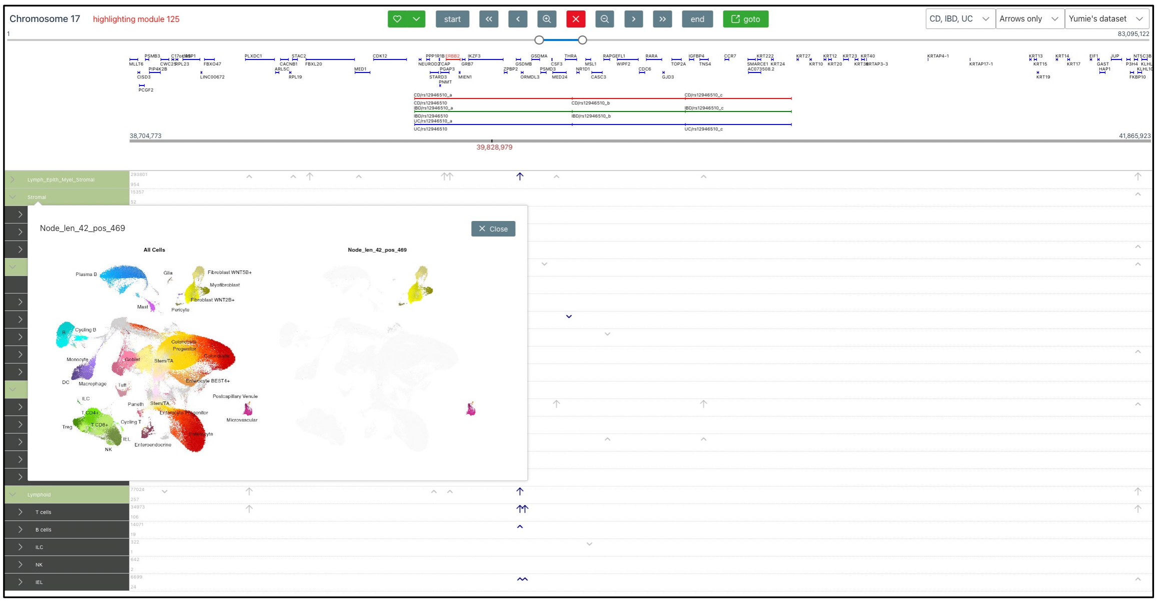

B

C

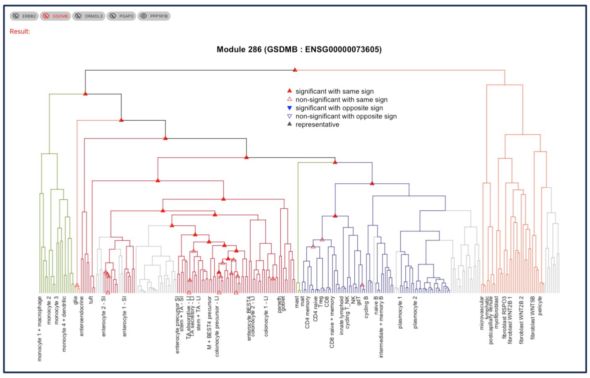

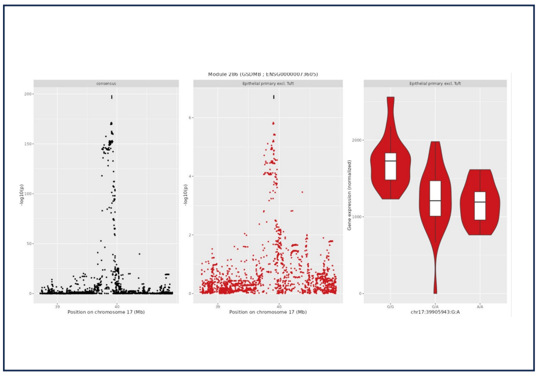

D

E

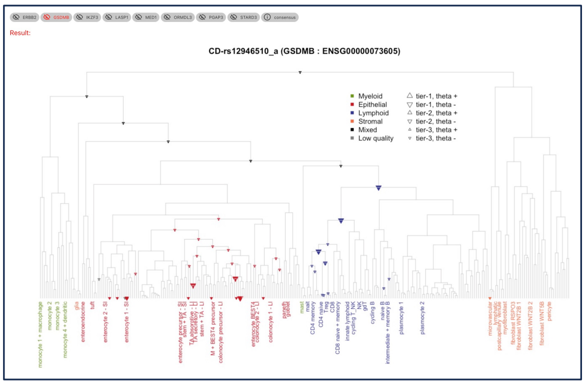

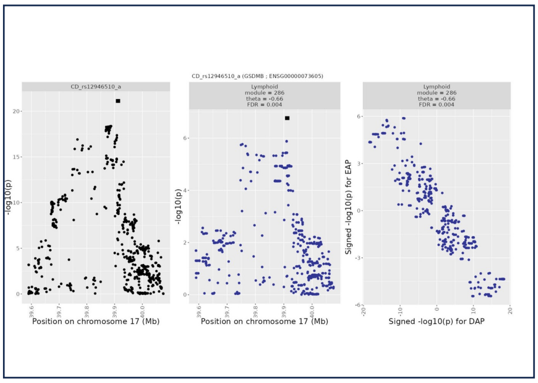

**(A)** The X axis of the main page corresponds to the genome coordinates showing, in the top panel, the position of the genes as well as the boundaries of color-coded disease risk loci (IBD, CD and UC in the example), and in the lower panel the expression levels (marbles, not shown) and eQTL activity (arrows, as in SFig. 4 for blood) for the different genes. The Y axis corresponds to the cell type hierarchy guided by the tree resulting from hierarchical clustering of the cell clusters and their annotations to specific intestinal cell types. Clicking on a level in the cell type hierarchy shows the corresponding cell selection on the 2D UMAP. **(B)** Clicking on an arrow (corresponding to an eQTL effect) calls a set of pages for each gene in the corresponding module, each with the hierarchical tree with symbols showing the activity of the eQTL (full circles: significant eQTL, empty circles: non-significant eQTL yet matching the consensus EAP of the module). **(C)** Clicking on the symbols calls a next page that shows the consensus EAP of the module, the EAP for the selected gene for the selected position in the tree and the corresponding violin plot for the top variants of the (consensus) EAP. **(D)** Clicking on a disease locus calls a set of images corresponding to trees for all genes that show a matching EAP-DAP for the selected risk locus. (E) Clicking on the corresponding symbols in the image, calls gene-specific pages showing the DAP for the corresponding risk locus, the EAP for the corresponding eQTL and the theta plot.

### Supplemental Figure 8:

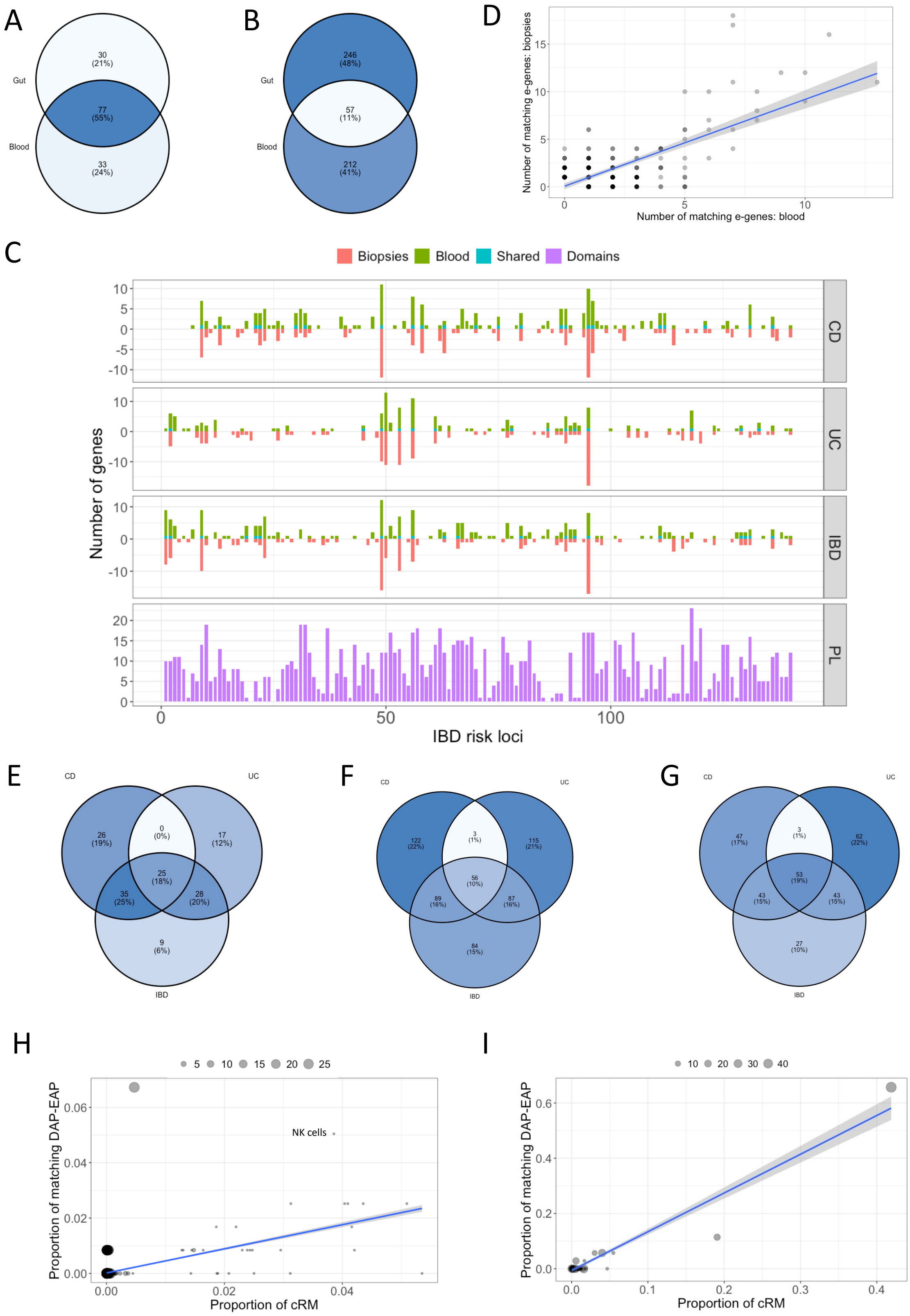

**(A)** Number of IBD risk loci with matching DAP-EAP in intestinal biopsies only (gut, n=30), in circulating immune populations only (blood, n=33), and both in gut and blood (n=77). **(B)** Number of e-genes with matching DAP-EAP in intestinal biopsies only (gut, n=246), in circulating immune populations only (blood, n=212), and both in gut and blood (n=57). **(C)** Number of e-genes with matching DAP-EAP in 206 IBD risk loci (ordered by chromosomal position along the X-axis) in blood (green bars) and biopsies (orange bars, numbers x -1). The blue bards correspond to the number of e-genes shared between blood and biopsies for a given risk locus. Numbers are given separately for CD, UC and IBD risk loci (see STable 21). Intestinal biopsies only (gut, n=246), in circulating immune populations only (blood, n=212), and both in gut and blood (n=57). The panel with the violet bars at the bottom, labeled "PL" (-eiotropy), reports the sum of the number of "trait domains" that are influenced by the pleiotropic genomic loci, defined by Watanabe *et al.*, 2019, that overlap with the corresponding IBD risk locus. **(D)** The number of matching e-genes (i.e., with matching DAP-EAP) in blood (X-axis) and biopsies (Y-axis) is highly correlated across the IBD risk loci, despite the fact that the corresponding e-genes mostly differ between blood and biopsies (see blue bars in panel C). **(E)** Numbers of risk loci with matching e-genes for CD, UC and IBD, with corresponding overlaps (i.e., risk loci with matching e-genes for CD and UC but not IBD (n=0), CD and IBD but not UC (n=35), UC and IBD but not CD (n=28), CD, UC and IBD (n=25)). **(F)** Numbers of matching e-genes for CD, UC and IBD, with corresponding overlaps (i.e., e-genes matching the DAP for CD and UC but not IBD (n=3), CD and IBD but not UC (n=89), UC and IBD but not CD (n=87), CD, UC and IBD (n=56)). **(G)** As for (F), yet restricting the analysis to 81 loci that are risk loci for both CD and UC. **(H)** The proportion of matching DAP-EAP involving "blood" cis-acting regulatory modules (= cRM) with a given "activity vector" (i.e., the combination of the 27 cell types in which the module is active) is correlated with the proportion of regulatory modules (= RM) with that "activity vector". Yet, some types of modules stand out. Regulatory modules that are active in all cell types account for only 0.0047 of all across-gene modules, yet account for 0.0672 of DAP-EAP matches (> 14-fold excess), reminiscent of Momozawa *et al.* [2018]. Each dot corresponds to one of 3,572 observed activity vectors. The size of the dot corresponds to the number of "1"'s in the vector (i.e., the number of cell-types out of 27 in which the module is active). The graph was obtained considering across-gene modules and the three diseases (CD, UC and IBD) combined. **(I)** The proportion of matching DAP-EAP involving "biopsy" regulatory modules encompassing a number of cell types (i.e., out of the 43 distinguished cell types; see also Fig. 2A) is correlated with the overall proportion of cognate regulatory modules (RM = *cis*-acting regulatory module). Yet regulatory modules assigned to the root of the hierarchical tree (Fig. 2F), and hence encompassing mostly modules that are active in many cell types, account for the highest proportion of DAP-EAP matches. Each dot corresponds to one of 64 observed cell-type groups. The size of the dot corresponds to the number of cell types (out of 43) included in the group. The graph was obtained considering gene-specific modules and the three diseases (CD, UC and IBD) combined.

### Supplemental Material 1: Comments on candidate drugs for repurposing to treat IBD with genetic support from this study.

**IL18R1 and IL18RAP.** IL18R1 and IL18RAP code for the IL18 receptor and its accessory subunit (enhancing IL18 binding), respectively. With IL1, IL18 is activated upon PAMP-recognition by the inflammasomes. Our data indicates that downregulation (predominantly) of both genes (IL18R1 and IL18RAP), primarily (but not exclusively) in circulating immune cells (including ILC, MAIT, and Th17), increases the risk to develop CD (predominantly), hence that "activating" both might have a beneficial effect. In agreement with these findings, IL18R KO mice are more susceptible to DSS-induced colitis [Takagi et al., 2003], and pre-treatment of mice with IL18 protected against DSS-induced colitis [Pu et al., 2019]. However, IL18 administration at later stages of DSS-induced colitis exacerbates symptoms [Pu et al., 2019], while administration of IL18 neutralizing antibodies reduces the severity of DSS-induced colitis [f.i. Ikegami et al., 2024]. In humans, abundance of IL18 is generally increased in IBD patients, both in blood and mucosa [Leach et al., 2008], while mendelian randomization indicates that increase in IL18 concentrations increases IBD risk [Mokry et al., 2019]. We observe an increase of IL18R1 mRNA abundance in naïve B cells of active CD patients, while Eldjarn *et al.* [2023] report an increased abundance of IL18R1 in IBD patients. Thus, the IL18 axis appears to have an anti-inflammatory effect prior to disease declaration, yet a pro-inflammatory effect after. Iboctadekin is reported as an agonist of IL18R1 and IL18RAP, and is in fact recombinant human IL18. Iboctadekin has been tested in the context of infectious diseases and oncology, but with mediocre results, and its development was discontinued. Given the apparent dual function of the IL18 axis in inflammation, the use of iboctadekin in the treatment of IBD appears difficult to support. Nevertheless, an iboctadekin-based treatment that might be considered in the context of IBD, is to prevent relapse, particularly after disease remission has been achieved following surgery, rather than its use during the active phase of the condition.

**CFTR.** Well-studied chloride channel essential for ion transport and fluid secretion in tissues, including the gastrointestinal tract, and playing an important role in mucin maturation in the intestine [Gustafsson et al., 2012]. Nullizygosity for *CFTR* causes cystic fibrosis (CF), while hemizygosity may increase resistance to infectious diseases [Bradbury, 1998]. The prevalence of IBD was shown to be 7 times higher in CF patients compared to controls [Lloyd-Still, 1994]. *A contrario*, CFTR hemizygosity was recently shown to protect against CD [Yu et al., 2024]. Also, glibenclamide, a (CFTR) inhibitor, was reported to protect rats against 2,4-dinitrobenzene sulfonic acid (DNBS)-induced gastrointestinal inflammation [Chidrawar & Alsuwayt, 2021]. In agreement with this, we herein shown that increased CFTR expression in intestinal epithelial and stromal cells is associated with increased risk for UC. Note, however, that there is some evidence (albeit non-significant) from our data that decreased expression of CFTR in specific enterocyte populations may increase CD risk, deserving further analysis in a larger dataset. Also, plasma levels of CFTR were found to be increased in IBD patients. This suggests that downregulating CFTR may improve IBD. Of note, CFTR has been shown to act as a pathogen recognition molecule that can induce NF-kB activation [Schroeder et al., 2002], and serves as epithelial receptor for *S. Typhi* transluminal migration [Pier et al., 1998]. Also, CFTR KO mice secreted more intestinal mucus [Norkina et al., 2004; Hodges et al., 2008], which plays a protective role in IBD. While most drug development targeting CFTR aims at restoring function in the context of CF, there exist at least one presumed CFTR inhibitor, namely crofelemer, approved for HIV-associated diarrhea [Tradrantip et al., 2010]. Crofelemer is a natural, (not so) small molecule derived from the *Croton lechleri* tree. It is taken orally to treat secretory diarrhea (as opposed to diarrhea resulting from an underlying inflammatory process) in various contexts. It has proven effective in HIV-seropositive patients on stable antiretroviral therapy [Macarthur et al., 2013]. Being primarily used for non-inflammatory diarrheal conditions, crofelemer may alleviate diarrheal symptoms IBD patients but it is unclear whether it also affects the underlying inflammatory process. Of note, inhibition of CFTR-mediated intestinal chloride secretion could be a potential therapy for bile acid diarrhea, a condition frequently present in IBD [Duan et al., 2019]. It seems important to determine whether the apparent effect of *CFTR* expression on IBD risk depends on CFTR's chloride channel activity (affected by crofelemer) or of distinct functions of this gene (such as pathogen recognition).

**LAMA2.** Codes for an $\alpha$ subunit of laminins, an essential component of the basement membrane with important roles in tissue organization. LAMA2 deficiency causes a muscular dystrophy accompanied by an exacerbated innate immunity response, autophagy and cell death [Jeudi et al., 2011]. Intriguingly, somatic LAMA2 mutations were recurrently observed in colonic mucosa of DSS-treated mice [He et al., 2021]. LAMA2 may play a role in intestinal barrier homeostasis. Along those lines, mutations in laminins have been associated with skin blistering [Schéele et al., 2007]. We observe that variants that increase *LAMA2* expression, including in precursor and mature enterocytes, consistently increase CD risk. However, *LAMA2* expression was found to be downregulated in fibroblasts of IBD inflamed terminal ileum. Ocriplasmin is an injectable (intravitreally) protease inhibitor acting on laminins (and fibronectin) that is used to treat vitreomacular adhesion [Stalman et al., 2012; Mastrapasqua et al., 2023]. Whether it is the down- or up-regulation, or any, that is desirable in this case is uncertain. There is a need for further exploration of the link between LAMA2 and IBD, which may also involve its role in autophagy [Mastrapasqua et al., 2023].

**CYP3A5.** Codes for a member of the cytochrome P450 monooxygenases metabolizing various endo- and xenobiotics. In particular CYP3A enzymes participate in bile acid biotransformation, which – if perturbed – could contribute to the disruption of gut homeostasis in colitis patients including by affecting microbiota composition [Hayes et al., 2016; Zhang et al., 2019; Lin et al., 2020]. We observe that an increase of CYP3A5 expression in naïve CD8 increases CD risk (tier 1). However, in enterocytes of the small intestine, the opposite seems to occur (negative theta), albeit with FDR above 0.10 but potentially more relevant. There was no detectable effect of disease status on CYP3A5 expression/abundance in blood, biopsies and plasma. There exist at least two small molecule inhibitors of CYP3A5: cobicistat and ritonavir, used to boost the effectiveness of other drugs (particularly antiretroviral). Ritonavir may have independent anti-inflammatory action, as well as affect the microbiome but this appears unrelated to the observed DAP-EAP match. Genotype at CYP3A5 variants has been proposed as a guide to tacrolimus dosage in the treatment of ulcerative colitis [Okabayashi et al., 2019; Yamamoto et al., 2020], yet this bears no relationship with DAP-EAP matching.

**PTGIR.** Encodes the G protein coupled receptor of prostacyclin (PGI2). Prostacyclin is mainly known as a potent vasodilator (and inhibitor of platelet activation). Concomitantly, small molecule agonists of PTGIR (often prostacyclin mimics) are primarily used in the treatment of pulmonary arterial hypertension. However, prostaglandins, including prostacyclin, are known to affect inflammation, having either pro-inflammatory (f.i. rheumatoid arthritis) or anti-inflammatory (f.i. allergic inflammation) effects depending on context [Stitham et al., 2011; Dorris & Peebles, 2012]. PTGIR has well-documented anti-inflammatory properties by inhibiting the production of pro-inflammatory cytokines and the immune response [Ricciotti et al., 2011]. Paradoxically, stimulating PTGIR signaling in cultured human monocytes augments calprotectin expression, a marker of inflammation [Karaky et al., 2022]. PTGIR may play a role in maintaining the intestinal epithelial barrier [Turner, 2009], and in regulating intestinal microcirculation which is impaired in IBD [f.i. Hatoum et al., 2003], and tissue repair. Of note, increased expression of the PTGER4 receptor for PGE2 (known to affect the immune system) has been associated with an increased risk for CD [Libioulle et al., 2007], yet PTGER4 knock-out mice are more susceptible to DDS-induced colitis, while PTGER4 agonists are protective [Kabashima et al., 2002]. We observe that reduced expression of PTGIR in multiple circulating T lymphocyte populations as well as resident monocytes is consistently associated with increased risk to develop UC. We don't see a clear effect of disease status on PTGIR expression/levels whether in circulating leukocytes, biopsies or plasma. We note that the use of cyclooxygenase inhibitors reportedly exacerbate symptoms in some IBD patients. As for IL18R1/IL18RAP, PTGIR activator-based treatments that could be considered in the context of IBD, are to prevent relapse, particularly after disease remission has been achieved following surgery, rather than their use during the active phase of the condition.

***INPP5D.*** Encoding inositol Polyphosphate-5-Phosphatase D (=SHIP1). SHIP acts as a multifunctional protein controlled by multiple regulatory inputs, and influences downstream signaling via both phosphatase-dependent and -independent means. SHIP1 has functions in B cells, T cells, NK cells, dendritic cells, mast cells, and macrophages [Pauls & Marshall, 2017]. Downregulation is generally considered to be pro-inflammatory. SHIP deficiency causes Crohn's disease-like ileitis in the mouse [Kerr et al., 2011], while deficiency in Inflammatory Bowel Disease is associated with severe Crohn's disease and peripheral T cell reduction [Fernandes et al., 2018]. Accordingly, we observe that *INPP5D* downregulation in circulating T lymphocytes, MAIT and NK cells, as well as resident monocytes increases CD risk. Also, INPP5D expression is decreased in circulating monocytes from IBD patients. The small molecule rosiptor, the most developed and characterized allosteric activator of INPP5D, has been unsuccessfully tested as anti-inflammatory drug to treat interstitial cystitis and bladder pain syndrome and its development has been discontinued [https://www.pharmaceutical-technology.com/news/aquinox-halts-development-drug-bladder-pain-syndrome/].
Binding of rosiptor to SHIP1 was shown to be weak when compared to other agonists which may in part explain poor clinical results [Chamberlain et al., 2020; Pedicone et al., 2021]. We conclude that rosiptor is probably not an optimal candidate for repurposing.

***NEK7.*** NIMA Related Kinase 7, plays a central role in assembly of the NLRP3 inflammasome [Shi et al., 2016]. Missense *NLRP3* mutation cause dominant congenital multisystem inflammatory diseases (Cinca (MIM 607115) and Muckle-Wells syndromes (MIM 191900)). Downregulating NEK7 by intraperitoneal injection of lentiviruses expression anti-NEK7 shRNAs attenuates DSS-induced colitis [Chen et al., 2019]. We observe that variants that increase NEK7 expression in circulating naïve B cells increase CD risk. Of note, however, these same variants appear to decrease NEK7 expression in gut-resident lymphoid and myeloid immune cells, albeit with FDR > 0.10, hence inversing the association with CD (decreased expression increases risk). Disease affects NEK7 expression but the sign of the affect depends on cell type: expression is increased in enterocytes from non-inflamed colon of patients when compared to controls, while expression is increased in some and decreased in other circulating immune cells of patients. Entrectinib (ENB) is a potent tyrosine multikinase small-molecule inhibitor that targets the NTRK, ROS1 and ALK oncogenes, approved by the FDA for the treatment of various tumors [Liu et al., 2018]. It was recently shown to bind to NEK7 R121, and interferes with its interaction with NLRP3. In mouse models, ENB effectively reduced symptoms of NLRP3 inflammasome-related diseases (other than colitis), indicating its potential as a therapeutic candidate [Jin et al., 2023]. ENB has a high safety profile and is well tolerated by almost all patients without cumulative toxicity [Jiang et al., 2022]. Inhibition of the inflammasome has shown beneficial effects in various inflammatory pathologies. ENB appears may be a repurposing candidate for IBD.

### *References*

Bradbury J. Why are cystic-fibrosis mutations so common? *The Lancet* **351**: 1409 (1998).

Chamberlain TC, Cheung ST, Yoon JSJ, Ming-Lum A, Gardill BR, Shakibakho S, Dzananovic E, Ban F, Samiea A, Jawanda K, Priatel J, Krystal G, Ong CJ, Cherkasov A, Andersen RJ, McKenna SA, Van Petegem F, Mui AL. Interleukin-10 and Small Molecule SHIP1 Allosteric Regulators Trigger Anti-inflammatory Effects through SHIP1/STAT3 Complexes. *iScience* **23**:101433 (2020). doi: 10.1016/j.isci.2020.101433.

Chen X, Liu G, Yuan Y, Wu G, Wang S, Yuan L. NEK7 interacts with NLRP3 to modulate the pyroptosis in inflammatory bowel disease via NF-κB signaling. *Cell Death Dis* **10**:906 (2019). doi: 10.1038/s41419-019-2157-1.

Chidrawar V, Alsuwayt B. Defining the role of CFTR channel blocker and ClC-2 activator in DNBS induced gastrointestinal inflammation. *Saudi Pharm J* **29**:291-304 (2021). doi: 10.1016/j.jsps.2021.02.005.

Danese S, Vermeire S, Hellstern P, Panaccione R, Rogler G, Fraser G, Kohn A, Desreumaux P, Leong RW, Comer GM, Cataldi F, Banerjee A, Maguire MK, Li C, Rath N, Beebe J, Schreiber S. Randomised trial and open-label extension study of an anti-interleukin-6 antibody in Crohn's disease (ANDANTE I and II). *Gut* **68**:40-48 (2019). doi: 10.1136/gutjnl-2017-314562.

Dorris SL, Peebles RS Jr. PGI2 as a regulator of inflammatory diseases. *Mediators Inflamm* **2012**:926968 (2012). doi: 10.1155/2012/926968.

Duan T, Cil O, Tse CM, Sarker R, Lin R, Donowitz M, Verkman AS. Inhibition of CFTR-mediated intestinal chloride secretion as potential therapy for bile acid diarrhea. *FASEB J* **33**:10924-10934 (2019). doi: 10.1096/fj.201901166R.

Fernandes S, Srivastava N, Sudan R, Middleton FA, Shergill AK, Ryan JC, Kerr WG. SHIP1 Deficiency in Inflammatory Bowel Disease Is Associated With Severe Crohn's Disease and Peripheral T Cell Reduction. *Front Immunol* **9**:1100 (2018). doi: 10.3389/fimmu.2018.01100.

Gustafsson JK, Ermund A, Ambort D, Johansson ME, Nilsson HE, Thorell K, Hebert H, Sjövall H, Hansson GC. Bicarbonate and functional CFTR channel are required for proper mucin secretion and link cystic fibrosis with its mucus phenotype. *J Exp Med* **209**:1263-1272 (2012). doi: 10.1084/jem.20120562.

Hanioka Y, Shimizu K, Yamagami K, Yao S, Nakamura R, Nakamura T, Goto H. Exacerbation of Ulcerative Colitis with Tocilizumab: A Report of Two Cases, One with Takayasu Arteritis and the Other with Relapsing Polychondritis. *Intern Med* **60**:1615-1620 (2021). doi: 10.2169/internalmedicine.5215-20.

Hatoum OA, Miura H, Binion DG. The vascular contribution in the pathogenesis of inflammatory bowel disease. *Am J Physiol Heart Circ Physiol* **285**: H1791–H1796 (2003)

Hayes MA, Li XQ, Grönberg G, Diczfalusy U, Andersson TB. CYP3A Specifically Catalyzes 1β-Hydroxylation of Deoxycholic Acid: Characterization and Enzymatic Synthesis of a Potential Novel Urinary Biomarker for CYP3A Activity. *Drug Metab Dispos* **44**:1480-1489 (2016). doi: 10.1124/dmd.116.070805.

He J, Han J, Liu J, Yang R, Wang J, Wang X, Chen X. Genetic and Epigenetic Impact of Chronic Inflammation on Colon Mucosa Cells. *Front Genet* **12**:722835 (2021). doi: 10.3389/fgene.2021.722835.

Hodges CA, Cotton CU, Palmert MR, Drumm ML. Generation of a conditional null allele for Cftr in mice. *Genesis* **46**:546-552 (2008). doi: 10.1002/dvg.20433.

Ikegami S, Maeda K, Urano T, Mu J, Nakamura M, Yamamura T, Sawada T, Ishikawa E, Yamamoto K, Muto H, Oishi A, Iida T, Mizutani Y, Ishikawa T, Kakushima N, Furukawa K, Ohno E, Honda T, Ishigami M, Kawashima H. Monoclonal Antibody Against Mature Interleukin-18 Ameliorates Colitis in Mice and Improves Epithelial Barrier Function. *Inflamm Bowel Dis* **30**:1353-1366 (2024). doi: 10.1093/ibd/izad292.

Ito H, Takazoe M, Fukuda Y, Hibi T, Kusugami K, Andoh A, Matsumoto T, Yamamura T, Azuma J, Nishimoto N, Yoshizaki K, Shimoyama T, Kishimoto T. A pilot randomized trial of a human anti-interleukin-6 receptor monoclonal antibody in active Crohn's disease. *Gastroenterology* **126**:989-996 (2004); discussion 947. doi: 10.1053/j.gastro.2004.01.012.

Jeudy S, Wardrop KE, Alessi A, Dominov JA (2011) Bcl-2 Inhibits the Innate Immune Response during Early Pathogenesis of Murine Congenital Muscular Dystrophy. *PLoS ONE* **6**: e22369. https://doi.org/10.1371/journal.pone.0022369

Jiang Q, Li M, Li H, Chen L. Entrectinib, a new multi-target inhibitor for cancer therapy. *Biomed Pharmacother* **150**:112974 (2022). doi: 10.1016/j.biopha.2022.112974.

Jin X, Liu D, Zhou X, Luo X, Huang Q, Huang Y. Entrectinib inhibits NLRP3 inflammasome and inflammatory diseases by directly targeting NEK7. *Cell Rep Med* **4**:101310 (2023). doi: 10.1016/j.xcrm.2023.101310.

Kabashima K, Saji T, Murata T, Nagamachi M, Matsuoka T, Segi E, Tsuboi K, Sugimoto Y, Kobayashi T, Miyachi Y, Ichikawa A, Narumiya S. The prostaglandin receptor EP4 suppresses colitis, mucosal damage and CD4 cell activation in the gut. *J Clin Invest* **109**:883-893 (2002). doi: 10.1172/JCI14459.

Karaky M, Boucher G, Mola S, Foisy S, Beauchamp C, Rivard ME, Burnette M, Gosselin H; iGenoMed Consortium; Bitton A, Charron G, Goyette P, Rioux JD. Prostaglandins and calprotectin are genetically and functionally linked to the Inflammatory Bowel Diseases. *PLoS Genet* **18**:e1010189 (2022). doi: 10.1371/journal.pgen.1010189.

Kerr WG, Park MY, Maubert M, Engelman RW. SHIP deficiency causes Crohn's disease-like ileitis. *Gut* **60**:177-188 (2011). doi: 10.1136/gut.2009.202283.

Leach ST, Messina I, Lemberg DA, Novick D, Rubenstein M, Day AS. Local and systemic interleukin-18 and interleukin-18-binding protein in children with inflammatory bowel disease. *Inflamm Bowel Dis* **14**:68-74 (2008). doi: 10.1002/ibd.20272.

Li S, Wang H, Peng B, Zhang M, Zhang D, Hou S, Guo Y, Ding J. Efalizumab binding to the LFA-1 alphaL I domain blocks ICAM-1 binding via steric hindrance. *Proc Natl Acad Sci USA* **106**:4349-4354 (2009). doi: 10.1073/pnas.0810844106.

Libioulle C, Louis E, Hansoul S, Sandor C, Farnir F, Franchimont D, Vermeire S, Dewit O, de Vos M, Dixon A, Demarche B, Gut I, Heath S, Foglio M, Liang L, Laukens D, Mni M, Zelenika D, Van Gossum A, Rutgeerts P, Belaiche J, Lathrop M, Georges M. Novel Crohn disease locus identified by genome-wide association maps to a gene desert on 5p13.1 and modulates expression of PTGER4. *PLoS Genet* **3**:e58 (2007). doi: 10.1371/journal.pgen.0030058.

Lin Q, Tan X, Wang W, Zeng W, Gui L, Su M, Liu C, Jia W, Xu L, Lan K. Species Differences of Bile Acid Redox Metabolism: Tertiary Oxidation of Deoxycholate is Conserved in Preclinical Animals. *Drug Metab Dispos* **48**:499-507 (2020). doi: 10.1124/dmd.120.090464.

Liu D, Offin M, Harnicar S, Li BT, Drilon A. Entrectinib: an orally available, selective tyrosine kinase inhibitor for the treatment of *NTRK*, *ROS1*, and *ALK* fusion-positive solid tumors. *Ther Clin Risk Manag* **14**:1247-1252 (2018). doi: 10.2147/TCRM.S147381.

Lloyd-Still JD. Crohn's disease and cystic fibrosis. *Dig Dis Sci* **39**:880-885 (1994). doi: 10.1007/BF02087437. PMID: 8149854.

Macarthur RD, Hawkins TN, Brown SJ, Lamarca A, Clay PG, Barrett AC, Bortey E, Paterson C, Golden PL, Forbes WP. Efficacy and safety of crofelemer for noninfectious diarrhea in HIV-seropositive individuals (ADVENT trial): a randomized, double-blind, placebo-controlled, two-stage study. *HIV Clin Trials* **14**:261-273 (2013). doi: 10.1310/hct1406-261.

Major EO. Progressive multifocal leukoencephalopathy in patients on immunomodulatory therapies. *Annu Rev Med* **61**:35-47 (2010). doi: 10.1146/annurev.med.080708.082655.

Mastrapasqua M, Rossi R, De Cosmo L, Resta A, Errede M, Bizzoca A, Zampatti S, Resta N, Giardina E, Ruggieri M, Virgintino D, Annese T, Laforgia N, Girolamo F. Autophagy increase in Merosin-Deficient Congenital Muscular Dystrophy type 1A. *Eur J Transl Myol* **33**:11501 (2023). doi: 10.4081/ejtm.2023.11501.

Mokry, L.E., Zhou, S., Guo, C. *et al.* Interleukin-18 as a drug repositioning opportunity for inflammatory bowel disease: A Mendelian randomization study. *Sci Rep* **9**: 9386 (2019). https://doi.org/10.1038/s41598-019-45747-2

Murphy CJ, Bentley E, Miller PE, McIntyre K, Leatherberry G, Dubielzig R, Giuliano E, Moore CP, Phillips TE, Smith PB, Prescott E, Miller JM, Thomas P, Scagliotti R, Esson D, Gadek T, O'Neill CA. The pharmacologic assessment of a novel lymphocyte function-associated antigen-1 antagonist (SAR 1118) for the treatment of keratoconjunctivitis sicca in dogs. *Invest Ophthalmol Vis Sci* **52**:3174-3180 (2011). doi: 10.1167/iovs.09-5078.

Norkina O, Burnett TG, De Lisle RC. Bacterial overgrowth in the cystic fibrosis transmembrane conductance regulator null mouse small intestine. *Infect Immun* **72**:6040-6049 (2004). doi: 10.1128/IAI.72.10.6040-6049.2004.

Okabayashi S, Kobayashi T, Saito E, Toyonaga T, Ozaki R, Sagami S, Nakano M, Tanaka J, Yagisawa K, Kuronuma S, Takeuchi O, Hibi T. Individualized treatment based on CYP3A5 single-nucleotide polymorphisms with tacrolimus in ulcerative colitis. *Intest Res* **17**:218-226 (2019). doi: 10.5217/ir.2018.00117.

Pauls SD, Marshall AJ. Regulation of immune cell signaling by SHIP1: A phosphatase, scaffold protein, and potential therapeutic target. *Eur J Immunol* **47**:932-945 (2017). doi: 10.1002/eji.201646795.

Pedicone C, Meyer ST, Chisholm JD, Kerr WG. Targeting SHIP1 and SHIP2 in Cancer. *Cancers (Basel)* **13**:890 (2021). doi: 10.3390/cancers13040890.

Pier GB, Grout M, Zaidi T, Meluleni G, Mueschenborn SS, Banting G, Ratcliff R, Evans MJ, Colledge WH. Salmonella typhi uses CFTR to enter intestinal epithelial cells. *Nature* **393**:79-82 (1998). doi: 10.1038/30006.

Pu Z, Che Y, Zhang W, Sun H, Meng T, Xie H, Cao L, Hao H. Dual roles of IL-18 in colitis through regulation of the function and quantity of goblet cells. *Int J Mol Med* **43**:2291-2302 (2019). doi: 10.3892/ijmm.2019.4156.

Ricciotti E, FitzGerald GA. Prostaglandins and inflammation. *Arterioscler Thromb Vasc Biol* **31**:986-1000 (2011). doi: 10.1161/ATVBAHA.110.207449.

Schéele S, Nyström A, Durbeej M, Talts JF, Ekblom M, Ekblom P. Laminin isoforms in development and disease. *J Mol Med (Berl)* **85**:825-836 (2007). doi: 10.1007/s00109-007-0182-5.

Schroeder TH, Lee MM, Yacono PW, Cannon CL, Gerçeker AA, Golan DE, Pier GB. CFTR is a pattern recognition molecule that extracts Pseudomonas aeruginosa LPS from the outer membrane into epithelial cells and activates NF-kappa B translocation. *Proc Natl Acad Sci USA* **99**:6907-6912 (2002). doi: 10.1073/pnas.092160899.

Shi H, Wang Y, Li X, Zhan X, Tang M, Fina M, Su L, Pratt D, Bu CH, Hildebrand S, Lyon S, Scott L, Quan J, Sun Q, Russell J, Arnett S, Jurek P, Chen D, Kravchenko VV, Mathison JC, Moresco EM, Monson NL, Ulevitch RJ, Beutler B. NLRP3 activation and mitosis are mutually exclusive events coordinated by NEK7, a new inflammasome component. *Nat Immunol* **17**:250-258 (2016). doi: 10.1038/ni.3333.

Sivakumar PV, Westrich GM, Kanaly S, Garka K, Born TL, Derry JM, Viney JL. Interleukin 18 is a primary mediator of the inflammation associated with dextran sulphate sodium induced colitis: blocking interleukin 18 attenuates intestinal damage. *Gut* **50**:812-820 (2002). doi: 10.1136/gut.50.6.812.

Stalmans P, Benz MS, Gandorfer A, Kampik A, Girach A, Pakola S, Haller JA; MIVI-TRUST Study Group. Enzymatic vitreolysis with ocriplasmin for vitreomacular traction and macular holes. *N Engl J Med* **367**:606-615 (2012). doi: 10.1056/NEJMoa1110823.

Stitham J, Midgett C, Martin KA, Hwa J. Prostacyclin: an inflammatory paradox. *Front Pharmacol* **2**:24 (2011). doi: 10.3389/fphar.2011.00024.

Ten Hove T, Corbaz A, Amitai H, Aloni S, Belzer I, Graber P, Drillenburg P, van Deventer SJ, Chvatchko Y, Te Velde AA. Blockade of endogenous IL-18 ameliorates TNBS-induced colitis by decreasing local TNF-alpha production in mice. *Gastroenterology* **121**:1372-1379 (2001). doi: 10.1053/gast.2001.29579.

Tradtrantip L, Namkung W, Verkman AS. Crofelemer, an antisecretory antidiarrheal proanthocyanidin oligomer extracted from Croton lechleri, targets two distinct intestinal chloride channels. *Mol Pharmacol* **77**:69-78 (2010). doi: 10.1124/mol.109.061051.

Turner JR. Intestinal mucosal barrier function in health and disease. *Nat Rev Immunol* **9**:799-809 (2009). doi: 10.1038/nri2653.

Xu X, Han Y, Deng J, Wang S, Zhuo S, Zhao K, Zhou W. Repurposing disulfiram with CuET nanocrystals: Enhancing anti-pyroptotic effect through NLRP3 inflammasome inhibition for treating inflammatory bowel diseases. *Acta Pharm Sin B* **14**:2698-2715 (2024). doi: 10.1016/j.apsb.2024.03.003.

Yamamoto Y, Nakase H, Matsuura M, Maruyama S, Masuda S. *CYP3A5* Genotype as a Potential Pharmacodynamic Biomarker for Tacrolimus Therapy in Ulcerative Colitis in Japanese Patients. Int J Mol Sci 21:4347 (2020). doi: 10.3390/ijms21124347.

Yu M, Zhang Q, Yuan K, Sazonovs A, Stevens C, IIBDGC, Anderson CA, Daly MJ, Huang H. Cystic fibrosis risk variants confer protective effects against inflammatory bowel disease in large-scale exome sequencing analysis.

Zhang J, Gao LZ, Chen YJ, Zhu PP, Yin SS, Su MM, Ni Y, Miao J, Wu WL, Chen H, Brouwer KLR, Liu CX, Xu L, Jia W, Lan K. Continuum of Host-Gut Microbial Co-metabolism: Host CYP3A4/3A7 are Responsible for Tertiary Oxidations of Deoxycholate Species. *Drug Metab Dispos* **47**:283-294 (2019). doi: 10.1124/dmd.118.085670.

### Supplemental material 2: References for STable 25

Abdelbaqi M, Chidlow JH, Matthews KM, Pavlick KP, Barlow SC, Linscott AJ, Grisham MB, Fowler MR, Kevil CG. Regulation of dextran sodium sulfate induced colitis by leukocyte beta 2 integrins. *Lab Invest* **86**:380-390 (2006). doi: 10.1038/labinvest.3700398.

Afzali B, Grönholm J, Vandrovcova J, O'Brien C, Sun HW, Vanderleyden I, Davis FP, Khoder A, Zhang Y, Hegazy AN, Villarino AV, Palmer IW, Kaufman J, Watts NR, Kazemian M, Kamenyeva O, Keith J, Sayed A, Kasperaviciute D, Mueller M, Hughes JD, Fuss IJ, Sadiyah MF, Montgomery-Recht K, McElwee J, Restifo NP, Strober W, Linterman MA, Wingfield PT, Uhlig HH, Roychoudhuri R, Aitman TJ, Kelleher P, Lenardo MJ, O'Shea JJ, Cooper N, Laurence ADJ. BACH2 immunodeficiency illustrates an association between super-enhancers and haploinsufficiency. *Nat Immunol* **18**:813-823 (2017). doi: 10.1038/ni.3753.

Allgayer H, Roisch U, Zehnter E, Ziegenhagen DJ, Dienes HP, Kruis W. Colonic ornithine decarboxylase in inflammatory bowel disease: ileorectal activity gradient, guanosine triphosphate stimulation, and association with epithelial regeneration but not the degree of inflammation and clinical features. *Dig Dis Sci* **l**:25-30 (2007). doi: 10.1007/s10620-006-9515-4.

Al-Rashed F, Ahmad Z, Thomas R, Melhem M, Snider AJ, Obeid LM, Al-Mulla F, Hannun YA, Ahmad R. Neutral sphingomyelinase 2 regulates inflammatory responses in monocytes/macrophages induced by TNF-α. *Sci Rep* **10**:16802 (2020). doi: 10.1038/s41598-020-73912-5.

Amatullah H, Fraschilla I, Digumarthi S, Huang J, Adiliaghdam F, Bonilla G, Wong LP, Rivard ME, Beauchamp C, Mercier V, Goyette P, Sadreyev RI, Anthony RM, Rioux JD, Jeffrey KL. Epigenetic reader SP140 loss of function drives Crohn's disease due to uncontrolled macrophage topoisomerases. *Cell* **185**:3232-3247 (2022) .e18. doi: 10.1016/j.cell.2022.06.048.

Anbazhagan AN, Ge Y, Priyamvada S, Kumar A, Jayawardena D, Palani ARV, Husain N, Kulkarni N, Kapoor S, Kaur P, Majumder A, Lin YD, Maletta L, Gill RK, Alrefai WA, Saksena S, Zadeh K, Hong S, Mohamadzadeh M, Dudeja PK. A Direct Link Implicating Loss of SLC26A6 to Gut Microbial Dysbiosis, Compromised Barrier Integrity, and Inflammation. *Gastroenterology* **167**:704-717.e3 (2024). doi: 10.1053/j.gastro.2024.05.002.

Andres PG, Beck PL, Mizoguchi E, Mizoguchi A, Bhan AK, Dawson T, Kuziel WA, Maeda N, MacDermott RP, Podolsky DK, Reinecker HC. Mice with a selective deletion of the CC chemokine receptors 5 or 2 are protected from dextran sodium sulfate-mediated colitis: lack of CC chemokine receptor 5 expression results in a NK1.1+ lymphocyte-associated Th2-type immune response in the intestine. *J Immunol* **164**:6303-6312 (2000). doi: 10.4049/jimmunol.164.12.6303.

Arampatzidou M, Schütte A, Hansson GC, Saftig P, Brix K. Effects of cathepsin K deficiency on intercellular junction proteins, luminal mucus layers, and extracellular matrix constituents in the mouse colon. *Biol Chem* **393**:1391-403 (2012). doi: 10.1515/hsz-2012-0204.

Astier A, Trescol-Biémont MC, Azocar O, Lamouille B, Rabourdin-Combe C. Cutting edge: CD46, a new costimulatory molecule for T cells, that induces p120CBL and LAT phosphorylation. *J Immunol* **164**:6091-6095 (2000). doi: 10.4049/jimmunol.164.12.6091.

Bakos E, Thaiss CA, Kramer MP, Cohen S, Radomir L, Orr I, Kaushansky N, Ben-Nun A, Becker-Herman S, Shachar I. CCR2 Regulates the Immune Response by Modulating the Interconversion and Function of Effector and Regulatory T Cells. *J Immunol* 198: 4659–4671 2017). https://doi.org/10.4049/jimmunol.1601458

Baranov MV, Bianchi F, Schirmacher A, van Aart MAC, Maassen S, Muntjewerff EM, Dingjan I, Ter Beest M, Verdoes M, Keyser SGL, Bertozzi CR, Diederichsen U, van den Bogaart G. The Phosphoinositide Kinase PIKfyve Promotes Cathepsin-S-Mediated Major Histocompatibility Complex Class II Antigen Presentation. *iScience* **11**:160-177 (2019). doi: 10.1016/j.isci.2018.12.015.

Beckmann, D., Römer-Hillmann, A., Krause, A. *et al.* Lasp1 regulates adherens junction dynamics and fibroblast transformation in destructive arthritis. *Nat Commun* **12**: 3624 (2021). https://doi.org/10.1038/s41467-021-23706-8

Belinky F, Nativ N, Stelzer G, Zimmerman S, Iny Stein T, Safran M, Lancet D. PathCards: multi-source consolidation of human biological pathways. *Database* **2015**: bav006 (2015). doi:10.1093/database/bav006.

Bessa Pereira C, Bocková M, Santos RF, Santos AM, Martins de Araújo M, Oliveira L, Homola J, Carmo AM. The Scavenger Receptor SSc5D Physically Interacts with Bacteria through the SRCR-Containing N-Terminal Domain. *Front Immunol* **7**:416 (2016). doi: 10.3389/fimmu.2016.00416.

Blunt K, Jacobse J, Allaman M, Washington M, Goettel J, Wilson K, Williams C, Short S. Gpx3 is required to mediate protective effects of Gpx1 loss in colitis. Inflammatory Bowel Diseases 28: S63 (2022). https://doi.org/10.1093/ibd/izac015.101

Bosa L, Batura V, Colavito D, Fiedler K, Gaio P, Guo C, Li Q, Marzollo A, Mescoli C, Nambu R, Pan J, Perilongo G, Warner N, Zhang S, Kotlarz D, Klein C, Snapper SB, Walters TD, Leon A, Griffiths AM, Cananzi M, Muise AM. Novel CARMIL2 loss-of-function variants are associated with pediatric inflammatory bowel disease. *Sci Rep* **11**:5945 (2021). doi: 10.1038/s41598-021-85399-9.

Bromberger T, Klapproth S, Rohwedder I, Wever J, Pcik R, Mittmann L, Min-Weissenhorn SJ, Reichel CA, Scheiermann C, Sperandio M, Moser M. Binding of Rap1 and Riam to Talin 1 fine-tune $\beta_2$-integrin activity during leukocyte traffickingf. *Front Immunol* **12**: 702345 (2021). https://doi.org/10.3389/fimmu.2021.702345.

Brüggemann TR, Carlo T, Krishnamoorthy N, Duvall MG, Abdulnour R-E E, Nijmeh J, Peh HY, Filippakis H, Croze RH, Goh B, Oh SF, Levy BD. Mouse phospholipid phosphatase 6 regulates dendritic cell cholesterol, micropinocytosis, and allergen sensitization. *iScience* **25**: 105185 (2022). https://doi.org/10.1016/j.isci.2022.105185

Buanne P, Di Carlo E, Caputi L, Brandolini L, Mosca M, Cattani F, Pellegrini L, Biordi L, Coletti G, Sorrentino C, Fedele G, Colotta F, Melillo G, Bertini R. Crucial pathophysiological role of CXCR2 in experimental ulcerative colitis in mice. *J Leukoc Biol* **82**:1239-1246 (2007). doi: 10.1189/jlb.0207118.

Cader MZ, Pereira de Alemida Rodrigues R, West JA, Sewell GW, Md-Ibrahim M, Reikine S, Sirago G, Unger LW, Belen Iglessias-Romero A, Ramshorn K, Haag L-M, Saveljeva S, Ebel J-F, Rosenstiel P, Kaneider NC, Lee JC, Lawley TD, Bradley A, Dougan, G, Modis Y, Griffin JL, Kaser A. FAMIN is a multifunctional purine enzyme enabling the purine nucleotide cycle. Cell 180: 278-295 (2020). https://doi.org/10.1016/j.cell.2019.12.017

Caldwell JM, Collins MH, Kemme KA, Sherrill JD, Wen T, Rochman M, Stucke EM, Amin L, Tai H, Putnam PE, Jiménez-Dalmaroni MJ, Wormald MR, Porollo A, Abonia JP, Rothenberg ME. Cadherin 26 is an alpha integrin-binding epithelial receptor regulated during allergic inflammation. *Mucosal Immunol* **10**:1190-1201 (2017). doi: 10.1038/mi.2016.120.

Calon A, Gross I, Lhermitte B, Martin E, Beck F, Duclos B, Kedinger M, Duluc I, Domon-Dell C, Freund JN. Different effects of the Cdx1 and Cdx2 homeobox genes in a murine model of intestinal inflammation. *Gut* **56**:1688-1695 (2007). doi: 10.1136/gut.2007.125542.

Capalbo G, Mueller-Kuller T, Koschmieder S, Klein H-U, Ottmann OG, Hoelzer D, Scheuring UJ. Characterization of ZC3H15 as a potential TRAF-2-interacting protein implicated in the NFκB pathway and overexpressed in AML. *Int J Oncol* **43**: 246-254 (2013). https://doi.org/10.3892/ijo.2013.1924

Carlo T, Kalwa H, Levy BD. 15-Epi-lipoxin A4 inhibits human neutrophil superoxide anion generation by regulating polyisoprenyl diphosphate phosphatase 1. *FASEB J.* **27**:2733-2741 (2013). doi: 10.1096/fj.12-223982.

Catalano A. The neuroimmune semaphorin-3A reduces inflammation and progression of experimental autoimmune arthritis. *J Immunol* **185**:6373-6383 (2010). doi: 10.4049/jimmunol.0903527.

Chabod M, Pedros C, Lamouroux L, Colacios C, Bernard I, Lagrange D, Balz-Hara D, Mosnier JF, Laboisse C, Vergnolle N, Andreoletti O, Roth MP, Liblau R, Fournié GJ, Saoudi A, Dejean AS. A spontaneous mutation of the rat Themis gene leads to impaired function of regulatory T cells linked to inflammatory bowel disease. *PLoS Genet* **8**:e1002461 (2012). doi: 10.1371/journal.pgen.1002461.

Chandra V, Bhagyaraj E, Nanduri R, Ahuja N, Gupta P. NR1D1 ameliorates Mycobacterium tuberculosis clearance through regulation of autophagy. *Autophagy* **11**:1987-1997 (2015). https://doi.org/10.1080/15548627.2015.1091140

Chapoval SP, Gao H, Fanaroff R, Keegan AD. Plexin B1 controls Treg numbers, limits allergic airway inflammation, and regulates mucins. *Front Immunol* **14**:1297354 (2024). doi: 10.3389/fimmu.2023.1297354.

Chatterjee M, Rauen T, Kis-Toth K, Kyttaris VC, Hedrich CM, Terhorst C, Tsokos GC. Increased Expression of SLAM Receptors SLAMF3 and SLAMF6 in Systemic Lupus Erythematosus T Lymphocytes Promotes Th17 Differentiation. *J Immunol* **188**: 1206–1212 (2012). https://doi.org/10.4049/jimmunol.1102773

Chen YG, Liu HX, Hong Y, Dong PZ, Liu SY, Gao YR, Lu D, Li T, Wang DY, Wu DD, Ji XY. PCNP is a novel regulator of proliferation, migration, and invasion in human thyroid cancer. *Int J Biol Sci* **18**:3605-3620 (2022). doi: 10.7150/ijbs.70394.

Chen R, Pang X, Li L, Zeng Z, Chen M, Zhang S. Ubiquitin-specific proteases in inflammatory bowel disease-related signalling pathway regulation. *Cell Death Dis* **13**:139 (2022). doi: 10.1038/s41419-022-04566-6.

Chen Y, Li J, Li S, Cheng Y, Fu X, Li J, Zhu L. Uncovering the Novel Role of NR1D1 in Regulating BNIP3-Mediated Mitophagy in Ulcerative Colitis. *Int J Mol Sci* **24**:14222 (2023). doi: 10.3390/ijms241814222.

Cheng Y, Hall TR, Xu X, Yung I, Souza D, Zheng J, Schiele F, Hoffmann M, Mbow ML, Garnett JP, Li J. Targeting uPA-uPAR interaction to improve intestinal epithelial barrier integrity in inflammatory bowel disease. *EbioMedicine* **75**:103758 (2022). doi: 10.1016/j.ebiom.2021.103758.

Chewchuk, S., Jahan, S. & Lohnes, D. Cdx2 regulates immune cell infiltration in the intestine. *Sci Rep* **11**,:15841 (2021). https://doi.org/10.1038/s41598-021-95412-w

Choi CY, Vo MT, Nicholas J, Choi YB. Autophagy-competent mitochondrial translation elongation factor TUFM inhibits caspase-8-mediated apoptosis. *Cell Death Differ* **29**:451-464 (2022). doi: 10.1038/s41418-021-00868-y.

Chuang HC, Hsueh CH, Hsu PM, Tsai CY, Shih YC, Chiu HY, Chen YM, Yu WK, Chen MH, Tan TH. DUSP8 induces TGF-β-stimulated IL-9 transcription and Th9-mediated allergic inflammation by promoting nuclear export of Pur-α. J Clin Invest **133**:e166269 (2023). doi: 10.1172/JCI166269.

Chulkina M, Beswick EJ, Pinchuk IV. Role of PD-L1 in Gut Mucosa Tolerance and Chronic Inflammation. *Int J Mol Sci* **21**:9165 (2020). doi: 10.3390/ijms21239165.

Courtney AH, Lo WL, Weiss A. TCR Signaling: Mechanisms of Initiation and Propagation. *Trends Biochem Sci* **43**:108-123 (2018). doi: 10.1016/j.tibs.2017.11.008.

Cui Y, Yu H, Zheng X, Peng R, Wang Q, Zhou Y, Wang R, Wang J, Qu B, Shen N, Guo Q, Liu X, Wang C. SENP7 Potentiates cGAS Activation by Relieving SUMO-Mediated Inhibition of Cytosolic DNA Sensing. *PLoS Pathog* **13**:e1006156 (2017). doi: 10.1371/journal.ppat.1006156.

Cui Y, Parashar S, Zahoor M, Needham PG, Mari M, Zhu M, Chen S, Ho HC, Reggiori F, Farhan H, Brodsky JL, Ferro-Novick S. A COPII subunit acts with an autophagy receptor to target endoplasmic reticulum for degradation. *Science* **365**:53-60 (2019). doi: 10.1126/science.aau9263.

Dambacher J, Beigel F, Zitzmann K, De Toni EN, Göke B, Diepolder HM, Auernhammer CJ, Brand S. The role of the novel Th17 cytokine IL-26 in intestinal inflammation. *Gut* **58**:1207-1217 (2009). doi: 10.1136/gut.2007.130112.

Danne C, Michaudel C, Skerniskyte J, Planchais J, Magniez A, Agus A, Michel ML, Lamas B, Da Costa G, Spatz M, Oeuvray C, Galbert C, Poirier M, Wang Y, Lapière A, Rolhion N, Ledent T, Pionneau C, Chardonnet S, Bellvert F, Cahoreau E, Rocher A, Arguello RR, Peyssonnaux C, Louis S, Richard ML, Langella P, El-Benna J, Marteyn B, Sokol H. CARD9 in neutrophils protects from colitis and controls mitochondrial metabolism and cell survival. *Gut* **72**:1081-1092 (2023). doi: 10.1136/gutjnl-2022-326917.

de Andrade STQ, Guidugli TI, Borrego A, Rodrigues BLC, Fernandes NCCA, Guerra JM, de Sousa JG, Starobinas N, Jensen JR, Cabrera WHK, De Franco M, Ibañez OM, Massa S, Ribeiro OG. Slc11a1 gene polymorphism influences dextran sulfate sodium (DSS)-induced colitis in a murine model of acute inflammation. *Genes Immun* **24**:71-80 (2023). doi: 10.1038/s41435-023-00199-7.

Decout A, Katz JD, Venkatraman S, Ablasser A. The cGAS-STING pathway as a therapeutic target in inflammatory diseases. *Nat Rev Immunol* **21**:548-569 (2021). doi: 10.1038/s41577-021-00524-z.

Deiss LP, Feinstein E, Berissi H, Cohen O, Kimchi A. Identification of a novel serine/threonine kinase and a novel 15-kD protein as potential mediators of the gamma interferon-induced cell death. *Genes Dev* **9**:15-30 (1995). doi: 10.1101/gad.9.1.15.

Dimitrov G, Bamberger S, Navard C, Dreux S, Badens C, Bourgeois P, Buffat C, Hugot JP, Fabre A. Congenital Sodium Diarrhea by mutation of the SLC9A3 gene. *Eur J Med Genet*. **262**:103712 (2019). doi: 10.1016/j.ejmg.2019.103712.

Dokoshi T, Zhang LJ, Nakatsuji T, Adase CA, Sanford JA, Paladini RD, Tanaka H, Fujiya M, Gallo RL. Hyaluronidase inhibits reactive adipogenesis and inflammation of colon and skin. *JCI Insight* **3**:e123072 (2018). doi: 10.1172/jci.insight.123072.

Dotan I, Allez M, Danese S, Keir M, Tole S, McBride J. The role of integrins in the pathogenesis of inflammatory bowel disease: Approved and investigational anti-integrin therapies. *Med Res Rev* **40**:245-262 (2020). doi: 10.1002/med.21601.

Dovey CM, Diep J, Clarke BP, Hale AT, McNamara DE, Guo H, Brown NW Jr, Cao JY, Grace CR, Gough PJ, Bertin J, Dixon SJ, Fiedler D, Mocarski ES, Kaiser WJ, Moldoveanu T, York JD, Carette JE. MLKL Requires the Inositol Phosphate Code to Execute Necroptosis. *Mol Cell* **70**:936-948.e7 (2018). doi: 10.1016/j.molcel.2018.05.010.

Du G, Xiong L, Li X, Zhuo Z, Zhuang X, Yu Z, Wu L, Xiao D, Liu Z, Jie M, Liu X, Guo Z, Chen H. Peroxisome elevation induces stem cell differentiation and intestinal epithelial repair. *Dev Cell* **53**: 169-184.e11 (2020). https://doi.org/10.1016/j.devcel.2020.03.002

Dzikiewicz-Krawczyk A, Kok K, Slezak-Prochazka I, Robertus JL, Bruining J, Tayari MM, Rutgers B, de Jong D, Koerts J, Seitz A, Li J, Tillema B, Guikema JE, Nolte IM, Diepstra A, Visser L, Kluiver J, van den Berg A. ZDHHC11 and ZDHHC11B are critical novel components of the oncogenic MYC-miR-150-MYB network in Burkitt lymphoma. *Leukemia* **31**:1470-1473 (2017). doi: 10.1038/leu.2017.94.

Eijkelkamp N, Heijnen CJ, Lucas A, Premont RT, Elsenbruch S, Schedlowski M, Kavelaars A. G protein-coupled receptor kinase 6 controls chronicity and severity of dextran sodium sulphate-induced colitis in mice. *Gut* **56**:847-854 (2007). doi: 10.1136/gut.2006.107094.

Eldjarn GH, Ferkingstad E, Lund SH, Helgason H, Magnusson OT, Gunnarsdottir K, Olafsdottir TA, Halldorsson BV, Olason PI, Zink F, Gudjonsson SA, Sveinbjornsson G, Magnusson MI, Helgason A, Oddsson A, Halldorsson GH, Magnusson MK, Saevarsdottir S, Eiriksdottir T, Masson G, Stefansson H, Jonsdottir I, Holm H, Rafnar T, Melsted P, Saemundsdottir J, Norddahl GL, Thorleifsson G, Ulfarsson MO, Gudbjartsson DF, Thorsteinsdottir U, Sulem P, Stefansson K. Large-scale plasma proteomics comparisons through genetics and disease associations. *Nature* **622**:348-358 (2023). doi: 10.1038/s41586-023-06563-x.

Esworthy RS, Aranda R, Martín MG, Doroshow JH, Binder SW, Chu FF. Mice with combined disruption of Gpx1 and Gpx2 genes have colitis. *Am J Physiol Gastrointest Liver Physiol* **281**:G848-55 (2001). doi: 10.1152/ajpgi.2001.281.3.G848.

Fan H, Hall P, Santos LL, Gregory JL, Fingerle-Rowson G, Bucala R, Morand EF, Hickey MJ. Macrophage migration inhibitory factor and CD74 regulate macrophage chemotactic responses via MAPK and Rho GTPase. *J Immunol* **186**:4915-4924 (2011). doi: 10.4049/jimmunol.1003713.

Fan F, He Z, Kong LL, Chen Q, Yuan Q, Zhang S, Ye J, Liu H, Sun X, Geng J, Yuan L, Hong L, Xiao C, Zhang W, Sun X, Li Y, Wang P, Huang L, Wu X, Ji Z, Wu Q, Xia NS, Gray NS, Chen L, Yun CH, Deng X, Zhou D. Pharmacological targeting of kinases MST1 and MST2 augments tissue repair and regeneration. *Sci Transl Med* **8**:352ra108 (2016). doi: 10.1126/scitranslmed.aaf2304.

Farooq SM, Stillie R, Svensson M, Svanborg C, Strieter RM, Stadnyk AW. Therapeutic effect of blocking CXCR2 on neutrophil recruitment and dextran sodium sulfate-induced colitis. *J Pharmacol Exp Ther* **329**:123-129 (2009). doi: 10.1124/jpet.108.145862.

Farr L, Ghosh S, Moonah S. Role of MIF Cytokine/CD74 Receptor Pathway in Protecting Against Injury and Promoting Repair. *Front Immunol* **11**:1273 (2020). doi: 10.3389/fimmu.2020.01273.

Fathman JW, Gurish MF, Hemmers S, Bonham K, Friend DS, Grusby MJ, Glimcher LH, Mowen KA. NIP45 controls the magnitude of the type 2 T helper cell response. *Proc Natl Acad Sci USA* **107**:3663-3668 (2010). doi: 10.1073/pnas.0914700107.

Fernandes S, Srivastava N, Sudan R, Middleton FA, Shergill AK, Ryan JC, Kerr WG. SHIP1 Deficiency in Inflammatory Bowel Disease Is Associated With Severe Crohn's Disease and Peripheral T Cell Reduction. *Front Immunol* **9**:1100 (2018). doi: 10.3389/fimmu.2018.01100.

Fraschilla I, Amatullah H, Rahman RU, Jeffrey KL. Immune chromatin reader SP140 regulates microbiota and risk for inflammatory bowel disease. *Cell Host Microbe* **30**:1370-1381.e5. (2022) doi: 10.1016/j.chom.2022.08.018.

Fremder M, Kim SW, Khamaysi A, Shimshilashvili L, Eini-Rider H, Park IS, Hadad U, Cheon JH, Ohana E. A transepithelial pathway delivers succinate to macrophages, thus perpetuating their pro-inflammatory metabolic state. *Cell Rep* **36**:109521 (2021). doi: 10.1016/j.celrep.2021.109521.

Gahloth, D., Heaven, G., Jowitt, T.A. *et al.* The open architecture of HD-PTP phosphatase provides new insights into the mechanism of regulation of ESCRT function. *Sci Rep* **7**: 9151 (2017). https://doi.org/10.1038/s41598-017-09467-9

Genau HM,Huber J, Bachieri F, Akutsu M, Dötsch V, Farhan H, Rogov V, Behrends C. CUL3-KBTBD6/KBTBD7 Ubiquitin Ligase Cooperates with GABARAP Proteins to Spatially Restrict TIAM1-RAC1 Signaling. *Molecular Cell* **57**: 995-1010 (2015). https://doi.org/10.1016/j.molcel.2014.12.040

Goettel JA, Scott Algood HM, Olivares-Villagómez D, Washington MK, Chaturvedi R, Wilson KT, Van Kaer L, Polk DB. KSR1 protects from interleukin-10 deficiency-induced colitis in mice by suppressing T-lymphocyte interferon-γ production. *Gastroenterology* **140**:265-274 (2011). doi: 10.1053/j.gastro.2010.09.041.

Gordon M, El-Kalla M, Zhao Y, Fiteih Y, Law J, Volodko N, Anwar-Mohamed A, El-Kadi AO, Liu L, Odenbach J, Thiesen A, Onyskiw C, Ghazaleh HA, Park J, Lee SB, Yu VC, Fernandez-Patron C, Alexander RT, Wine E, Baksh S. The tumor suppressor gene, RASSF1A, is essential for protection against inflammation -induced injury. *PLoS One* **8**:e75483 (2013). doi: 10.1371/journal.pone.0075483.

Goswami AB, Karadarević D, Castaño-Rodríguez N. Immunity-related GTPase IRGM at the intersection of autophagy, inflammation, and tumorigenesis. *Inflamm Res* **71**:785-795 (2022). doi: 10.1007/s00011-022-01595-x.

Greulich W, Wagner M, Gaidt MM, Stafford C, Cheng Y, Linder A, Carell T, Hornung V. TLR8 Is a Sensor of RNase T2 Degradation Products. *Cell* **179**:1264-1275.e13 (2019). doi: 10.1016/j.cell.2019.11.001.

Guha P, Tyagi R, Chowdhury S, Reilly L, Fu C, Xu R, Resnick AC, Snyder SH. IPMK Mediates Activation of ULK Signaling and Transcriptional Regulation of Autophagy Linked to Liver Inflammation and Regeneration. *Cell Rep* **26**:2692-2703.e7 (2019). doi: 10.1016/j.celrep.2019.02.013.

Hainzl E, Stockinger S, Rauch I, Heider S, Berry D, Lassnig C, Schwab C, Rosebrock F, Milinovich G, Schlederer M, Wagner M, Schleper C, Loy A, Urich T, Kenner L, Han X, Decker T, Strobl B, Müller M. Intestinal Epithelial Cell Tyrosine Kinase 2 Transduces IL-22 Signals To Protect from Acute Colitis. *J Immunol* **195**:5011-24 (2015). doi: 10.4049/jimmunol.1402565.

Hallows KR, Kobinger GP, Wilson JM, Witters LA, Foskett JK. Physiological modulation of CFTR activity by AMP-activated protein kinase in polarized T84 cells. *Am J Physiol Cell Physiol* **284**:C1297-308 (2003). doi: 10.1152/ajpcell.00227.2002.

Hamaoui D, Subtil A. ATG16L1 functions in cell homeostasis beyond autophagy. *FEBS J* **289**: 1779-1800 (2022). https://doi.org/10.1111/febs.15833

Hao L, Zhu G, Lu Y, Wang M, Jules J, Zhou X, Chen W. Deficiency of cathepsin K prevents inflammation and bone erosion in rheumatoid arthritis and periodontitis and reveals its shared osteoimmune role. *FEBS Lett* **589**:1331-1339 (2015). doi: 10.1016/j.febslet.2015.04.008.

Harit, K., Bhattacharjee, R., Matuschewski, K. *et al.* The deubiquitinating enzyme OTUD7b protects dendritic cells from TNF-induced apoptosis by stabilizing the E3 ligase TRAF2. *Cell Death Dis* **14**: 480 (2023). https://doi.org/10.1038/s41419-023-06014-5

Hassan SW, Doody KM, Hardy S, Uetani N, Cournoyer D, Tremblay ML. Increased susceptibility to dextran sulfate sodium induced colitis in the T cell protein tyrosine phosphatase heterozygous mouse. *PLoS One* **5**:e8868 (2010). doi: 10.1371/journal.pone.0008868.

He R, Chen J, Zhao Z, Shi C, Du Y, Yi M, Feng L, Peng Q, Cui Z, Gao R, Wang H, Huang Y, Liu Z, Wang C. T-cell activation Rho GTPase-activating protein maintains intestinal homeostasis by regulating intestinal T helper cells differentiation through the gut microbiota. *Front Microbiol* **13**:1030947 (2023). doi: 10.3389/fmicb.2022.1030947.

Hechter D, Vahkal B, Tied T, Good SV. Reviewing the physiological roles of the novel hormone-receptor pair INSL5-RXFP4: a protective energy sensor? *J Mol End* **6**:R45-R62 (2022)

Helke, K., Angel, P., Lu, P. *et al.* Ceramide Synthase 6 Deficiency Enhances Inflammation in the DSS model of Colitis. *Sci Rep* **8**: 1627 (2018). https://doi.org/10.1038/s41598-018-20102-z

Hernandez-Lara MA, Richard J, Deshpande DA. Diacylglycerol kinase is a keystone regulator of signaling relevant to the pathophysiology of asthma. *Am J Physiol Lung Cell Mol Physiol* **327**:L3-L18 (2024). doi: 10.1152/ajplung.00091.2024.

Hirai T, Kanda T, Sato K, Takaishi M, Nakajima M, Yamamoto M, Kamijima R, DiGiovanni J, Sano S. Cathepsin K Is Involved in Development of Psoriasis-like Skin Lesions through TLR-Dependent Th17 Activation. *J Immunol* **190**: 4805–4811 (2013). https://doi.org/10.4049/jimmunol.1200901

Homer CR, Richmond AL, Rebert NA, Achkar JP, McDonald C. ATG16L1 and NOD2 interact in an autophagy-dependent antibacterial pathway implicated in Crohn's disease pathogenesis. *Gastroenterology* **139**:1630-1641.e1-2 (2010). doi: 10.1053/j.gastro.2010.07.006.

Hong SS, Marotte H, Courbon G. *et al. PUMA* gene delivery to synoviocytes reduces inflammation and degeneration of arthritic joints. *Nat Commun* **8**: 146 (2017). https://doi.org/10.1038/s41467-017-00142-1

Hong G, Zhao Y, Li Q, Liu S. Fut2 deficiency aggravates chronic colitis through 2-oxindole-AHR mediated cGAS-STING pathway. *Int Immunopharmacol* **137**:112512 (2024). doi: 10.1016/j.intimp.2024.112512.

Hossain M, Qadri SM, Xu N, Su Y, Cayabyab FS, Heit B, Liu L. Endothelial LSP1 Modulates Extravascular Neutrophil Chemotaxis by Regulating Nonhematopoietic Vascular PECAM-1 Expression. *J Immunol* 195: 2408–2416 (2015). https://doi.org/10.4049/jimmunol.1402225

Howlader MA, Demina EP, Samarani S, Guo T, Caillon A, Ahmad A, Phezhetsky AV, Cairo CW. The Janus-like role of neuraminidase isoenzymes in inflammation. FASEB J 36: e22285 (2022). https://doi.org/10.1096/fj.202101218R

Hrdinka M, Fiil BK, Zucca M, Leske D, Bagola K, Yabal M, Elliott PR, Damgaard RB, Komander D, Jost PJ, Gyrd-Hansen M. CYLD Limits Lys63- and Met1-Linked Ubiquitin at Receptor Complexes to Regulate Innate Immune Signalling. *Cell Rep* 14:2846-2858 (2016). doi: 10.1016/j.celrep.2016.02.062.

Hu, H., Brittain, G., Chang, JH. *et al.* OTUD7B controls non-canonical NF-κB activation through deubiquitination of TRAF3. *Nature* **494**: 371–374 (2013). https://doi.org/10.1038/nature11831

Hu H, Wang H, Xiao Y, Jin J, Chang JH, Zou Q, Xie X, Cheng X, Sun SC. Otud7b facilitates T cell activation and inflammatory responses by regulating Zap70 ubiquitination. *J Exp Med* **213**:399-414 (2016). doi: 10.1084/jem.20151426.

Huang C, Hedl M, Ranjan K, Abraham C. LACC1 Required for NOD2-Induced, ER Stress-Mediated Innate Immune Outcomes in Human Macrophages and LACC1 Risk Variants Modulate These Outcomes. *Cell Rep* **29**:4525-4539.e4 (2019). doi: 10.1016/j.celrep.2019.

Inamdar M, Wulligundam P, Sinha S, Sinha A, Abe T, Kiyonari H, VijayRaghavan K. The ASRIJ interactor OCIAD2 is essential for mouse hematopoietic homeostasis. Exp Hematology 88 S64 (2020). https://doi.org/10.1016/j.exphem.2020.09.101

Inaoki M, Sato S, Weintraub BC, Goodnow CC, Tedder TF. CD19-regulated signaling thresholds control peripheral tolerance and autoantibody production in B lymphocytes. *J Exp Med* **186**:1923-1931 (1997). doi: 10.1084/jem.186.11.1923.

Ingoglia F, Visigalli R, Rotoli BM, Barilli A, Riccardi B, Puccini P, Milioli M, Di Lascia M, Bernuzzi G, Dall'Asta V. Human macrophage differentiation induces OCTN2-mediated L-carnitine transport through stimulation of mTOR-STAT3 axis. *J Leukoc Biol* **101**:665-674 (2017). doi: 10.1189/jlb.1A0616-254R.

Ito T, Carson WF 4th, Cavassani KA, Connett JM, Kunkel SL. CCR6 as a mediator of immunity in the lung and gut. Exp Cell Res 317:613-619 (2011). doi: 10.1016/j.yexcr.2010.12.018.

Jabado N, Jankowski A, Dougaparsad S, Picard V, Grinstein S, Gros P. Natural resistance to intracellular infections: natural resistance-associated macrophage protein 1 (Nramp1) functions as a pH-dependent manganese transporter at the phagosomal membrane. *J Exp Med* **192**:1237-1248 (2000). doi: 10.1084/jem.192.9.1237.

Jang SW, Hwang SS, Kim HS, Kim MK, Lee WH, Hwang SU, Gwak J, Yew SK, Flavell RA, Lee GR. Homeobox protein Hhex negatively regulates Treg cells by inhibiting Foxp3 expression and function. *Proc Natl Acad Sci USA* **116**:25790-25799 (2019). doi: 10.1073/pnas.1907224116.

Jeudy S, Wardrop KE, Alessi A, Dominov JA (2011) Bcl-2 Inhibits the Innate Immune Response during Early Pathogenesis of Murine Congenital Muscular Dystrophy. *PLoS ONE* **6**: e22369. https://doi.org/10.1371/journal.pone.0022369

Jiang HR, Gilchrist DS, Popoff JF, Jamieson SE, Truscott M, White JK, Blackwell JM. Influence of Slc11a1 (formerly Nramp1) on DSS-induced colitis in mice. *J Leukoc Biol* **85**:703-710 (2009). doi: 10.1189/jlb.0708397.

Jin X, Liu D, Zhou X, Luo X, Huang Q, Huang Y. Entrectinib inhibits NLRP3 inflammasome and inflammatory diseases by directly targeting NEK7. *Cell Rep Med* **4**:101310 (2023). doi: 10.1016/j.xcrm.2023.101310.

Joosse ME, Nederlof I, Walker LSK, Samsom JN. Tipping the balance: inhibitory checkpoints in intestinal homeostasis. *Mucosal Immunol* **12**:21-35 (2019). doi: 10.1038/s41385-018-0113-5.

Južnić L, Peuker K, Strigli A, Brosch M, Herrmann A, Häsler R, Koch M, Matthiesen L, Zeissig Y, Löscher BS, Nuber A, Schotta G, Neumeister V, Chavakis T, Kurth T, Lesche M, Dahl A, von Mässenhausen A, Linkermann A, Schreiber S, Aden K, Rosenstiel PC, Franke A, Hampe J, Zeissig S. SETDB1 is required for intestinal epithelial differentiation and the prevention of intestinal inflammation. *Gut* **70**:485-498 (2021). doi: 10.1136/gutjnl-2020-321339.

Kadiyska T, Tourtourikov I, Popmihaylova AM, Kadian H, Chavoushian A. Role of *TNFSF15* in the intestinal inflammatory response. *World J Gastrointest Pathophysiol* 9:73-78 (2018). doi: 10.4291/wjgp.v9.i4.73.

Kaminski S, Hermann-Kleiter N, Meisel M, Thuille N, Cronin S, Hara H, Fresser F, Penninger JM, Baier G. Coronin 1A is an essential regulator of the TGFβ receptor/SMAD3 signaling pathway in Th17 CD4(+) T cells. *J Autoimmun* **37**:198-208 (2011). doi: 10.1016/j.jaut.2011.05.018.

Karaky M, Boucher G, Mola S, Foisy S, Beauchamp C, Rivard ME, Burnette M, Gosselin H; iGenoMed Consortium; Bitton A, Charron G, Goyette P, Rioux JD. Prostaglandins and calprotectin are genetically and functionally linked to the Inflammatory Bowel Diseases. *PLoS Genet* **18**:e1010189 (2022). doi: 10.1371/journal.pgen.1010189.

Katayama H, Mori T, Seki Y, Anraku M, Iseki M, Ikutani M, Iwasaki Y, Yoshida N, Takatsu K, Takaki S. Lnk prevents inflammatory $CD8^+$ T-cell proliferation and contributes to intestinal homeostasis. *Eur J Immunol* **44**:1622-32 (2014). doi: 10.1002/eji.201343883.

Kattah MG, Shao L, Rosli YY, Shimizu H, Whang MI, Advincula R, Achacoso P, Shah S, Duong BH, Onizawa M, Tanbun P, Malynn BA, Ma A. A20 and ABIN-1 synergistically preserve intestinal epithelial cell survival. *J Exp Med* **215**:1839-1852 (2018). doi: 10.1084/jem.20180198.

Kawabe K, Lindsay D, Braitch M, Fahey AJ, Showe L, Constantinescu CS. IL-12 inhibits glucocorticoid-induced T cell apoptosis by inducing GMEB1 and activating PI3K/Akt pathway. Immunobiol 217: 118-123 (2012). https://doi.org/10.1016/j.imbio.2011.07.018

Kawashima A, Karasawa T, Tago K, Kimura H, Kamata R, Usui-Kawanishi F, Watanabe S, Ohta S, Funakoshi-Tago M, Yanagisawa K, Kasahara T, Suzuki K, Takahashi M. ARIH2 Ubiquitinates NLRP3 and Negatively Regulates NLRP3 Inflammasome Activation in Macrophages. *J Immunol* **199**:3614-3622 (2017). doi: 10.4049/jimmunol.1700184.

Kerr WG, Park MY, Maubert M, Engelman RW. SHIP deficiency causes Crohn's disease-like ileitis. *Gut* **60**:177-188 (2011). doi: 10.1136/gut.2009.202283.

Kiela PR, Laubitz D, Larmonier CB, Midura-Kiela MT, Lipko MA, Janikashvili N, Bai A, Thurston R, Ghishan FK. Changes in mucosal homeostasis predispose NHE3 knockout mice to increased susceptibility to DSS-induced epithelial injury. *Gastroenterology* **137**:965-75, 975.e1-10 (2009). doi: 10.1053/j.gastro.2009.05.043.

Kim TW, Park HJ, Choi EY, Jung KC. Overexpression of CIITA in T cells aggravates Th2-mediated colitis in mice. *J Korean Med Sci* **21**:877-882 (2006). doi: 10.3346/jkms.2006.21.5.877.

Kim E, Beon J, Lee S, Park SJ, Ahn H, Kim MG, Park JE, Kim W, Yuk JM, Kang SJ, Lee SH, Jo EK, Seong RH, Kim S. Inositol polyphosphate multikinase promotes Toll-like receptor-induced inflammation by stabilizing TRAF6. *Sci Adv* **3**:e1602296 (2017). doi: 10.1126/sciadv.1602296.

Kim YR, Volpert G, Shin KO, Kim SY, Shin SH, Lee Y, Sung SH, Lee YM, Ahn JH, Pewzner-Jung Y, Park WJ, Futerman AH, Park JW. Ablation of ceramide synthase 2 exacerbates dextran sodium sulphate-induced colitis in mice due to increased intestinal permeability. *J Cell Mol Med* **21**:3565-3578 (2017). doi: 10.1111/jcmm.13267.

Kim N, Kim TH, Kim C, Lee JE, Kang MG, Shin S, Jung M, Kim JS, Mun JY, Rhee HW, Park SY, Shin Y, Yoo JY. Intrinsically disordered region-mediated condensation of IFN-inducible SCOTIN/SHISA-5 inhibits ER-to-Golgi vesicle transport. *Dev Cell* **58**:1950-1966.e8 (2023). doi: 10.1016/j.devcel.2023.08.030.

Klunk, J., Vilgalys, T.P., Demeure, C.E. *et al.* Evolution of immune genes is associated with the Black Death. *Nature* **611**: 312–319 (2022). https://doi.org/10.1038/s41586-022-05349-x

Kolesnick R, Xing HR. Inflammatory bowel disease reveals the kinase activity of KSR1. *J Clin Invest* **114**:1233-1237 (2004). doi: 10.1172/JCI23441

Kong D, Shen Y, Liu G, Zuo S, Ji Y, Lu A, Nakamura M, Lazarus M, Stratakis CA, Breyer RM, Yu Y. PKA regulatory IIα subunit is essential for PGD2-mediated resolution of inflammation. *J Exp Med* **213**:2209-26 (2016). doi: 10.1084/jem.20160459.

Koren I, Reem E, Kimchi A. DAP1, a novel substrate of mTOR, negatively regulates autophagy. *Curr Biol* **20**:1093-1098 (2010). doi: 10.1016/j.cub.2010.04.041.

Kulkarni RM, Stuart WD, Gurusamy D, Waltz SE. Ron receptor signaling is protective against DSS-induced colitis in mice. *Am J Physiol Gastrointest Liver Physiol* **306**:G1065-1074 (2014). doi: 10.1152/ajpgi.00421.2013.

Kumar S, Jain A, Choi SW, da Silva GPD, Allers L, Mudd MH, Peters RS, Anonsen JH, Rusten TE, Lazarou M, Deretic V. Mammalian Atg8 proteins and the autophagy factor IRGM control mTOR and TFEB at a regulatory node critical for responses to pathogens. *Nat Cell Biol* **22**:973-985 (2020). doi: 10.1038/s41556-020-0549-1.

Laidlaw, B.J., Duan, L., Xu, Y. *et al.* The transcription factor Hhex cooperates with the corepressor Tle3 to promote memory B cell development. *Nat Immunol* **21**: 1082–1093 (2020). https://doi.org/10.1038/s41590-020-0713-6

Lantieri F, Bachetti T. OSM/OSMR and Interleukin 6 Family Cytokines in Physiological and Pathological Condition. *Int J Mol Sci* **23**:11096 (2022). doi: 10.3390/ijms231911096.

Latomanski EA, Newton HJ. Interaction between autophagic vesicles and the Coxiella-containing vacuole requires CLTC (clathrin heavy chain). *Autophagy* **14**:1710-1725 (2018). doi: 10.1080/15548627.2018.1483806.

Latour YL, McNamara KM, Allaman MM, Barry DP, Smith TM, Asim M, Williams KJ, Hawkins CV, Jacobse J, Goettel JA, Delgado AG, Piazuelo MB, Washington MK, Gobert AP, Wilson KT. Myeloid deletion of talin-1 reduces mucosal macrophages and protects mice from colonic inflammation. *Sci Rep* **13**:22368 (2023). doi: 10.1038/s41598-023-49614-z.

Latour YL, Allaman MM, Barry DP, Smith TM, Williams KJ, McNamara KM, Jacobse J, Goettel JA, Delgado AG, Piazuelo MB, Zhao S, Gobert AP, Wilson KT. Epithelial talin-1 protects mice from *citrobacter rodentium*-induced colitis by restricting bacterial crypt intrusion and enhancing t cell immunity. *Gut Microbes* **15**:2192623 (2023). doi: 10.1080/19490976.2023.2192623.

Laubitz D, Larmonier CB, Bai A, Midura-Kiela MT, Lipko MA, Thurston RD, Kiela PR, Ghishan FK. Colonic gene expression profile in NHE3-deficient mice: evidence for spontaneous distal colitis. *Am J Physiol Gastrointest Liver Physiol* **295**:G63-G77 (2008). doi: 10.1152/ajpgi.90207.2008.

Laura G, Liu Y, Fernandes K, Willis-Owen SAG, Ito K, Cookson WO, Moffatt MF, Zhang Y. ORMDL3 regulates poly I:C induced inflammatory responses in airway epithelial cells. *BMC Pulm Med* **21**:167 (2021). doi: 10.1186/s12890-021-01496-5.

Lechner K, Mott S, Al-Saifi R, Knipfer L, Wirtz S, Atreya R, Vieth M, Rath T, Fraass T, Winter Z, August A, Luban J, Zimmermann VS, Weigmann B, Neurath MF. Targeting of the Tec Kinase ITK Drives Resolution of T Cell-Mediated Colitis and Emerges as Potential Therapeutic Option in Ulcerative Colitis. *Gastroenterology* **161**:1270-1287.e19 (2021). doi: 10.1053/j.gastro.2021.06.072.

Lee SH, Yun S, Lee J, Kim MJ, Piao ZH, Jeong M, Chung JW, Kim TD, Yoon SR, Greenberg PD, Choi I. RasGRP1 is required for human NK cell function. *J Immunol* **183:**7931-8 (2009). doi: 10.4049/jimmunol.0902012.

Lee IY, Lim JM, Cho H, Kim E, Kim Y, Oh HK, Yang WS, Roh KH, Park HW, Mo JS, Yoon JH, Song HK, Choi EJ. MST1 Negatively Regulates TNFα-Induced NF-κB Signaling through Modulating LUBAC Activity. *Mol Cell* **73**:1138-1149.e6 (2019). doi: 10.1016/j.molcel.2019.01.022.

Lee NR, Kim BJ, Lee CH. *et al.* Role of 11β-hydroxysteroid dehydrogenase type 1 in the development of atopic dermatitis. *Sci Rep* **10**, 20237 (2020). https://doi.org/10.1038/s41598-020-77281-x

Lee JE, Kim N, Jung M, Mun JY, Yoo JY. SHISA5/SCOTIN restrains spontaneous autophagy induction by blocking contact between the ERES and phagophores. *Autophagy* **18**:1613-1628. (2022) doi: 10.1080/15548627.2021.

Lefebvre C, Legouis R, Culetto E. ESCRT and autophagies: endosomal functions and beyond. Sem Cell Dev Biol 74:21-28 (2018). https://doi.org/10.1016/j.semcdb.2017.08.014

Lei Y, Wen H, Ting JP. The NLR protein, NLRX1, and its partner, TUFM, reduce type I interferon, and enhance autophagy. *Autophagy* **9**:432-343 (2013). doi: 10.4161/auto.23026.

Lei CQ, Wu X, Zhong X, Jiang L, Zhong B, Shu HB. USP19 Inhibits TNF-α- and IL-1β-Triggered NF-κB Activation by Deubiquitinating TAK1. *J Immunol* **203**:259-268 (2019). doi: 10.4049/jimmunol.1900083.

Leonard A, Rahman A, Fazal F. Importins α and β signalling mediates endothelial cell inflammation and barrier disruption. *Cell Signal* **44**:103-117 (2018). doi: 10.1016/j.cellsig.2018.01.011.

Lepelletier Y, Moura IC, Hadj-Slimane R, Renand A, Fiorentino S, Baude C, Shirvan A, Barzilai A, Hermine O. Immunosuppressive role of semaphorin-3A on T cell proliferation is mediated by inhibition of actin cytoskeleton reorganization. *Eur J Immunol* **36**:1782-1793 (2006). doi: 10.1002/eji.200535601.

Li Q, Ching AK, Chan BC, Chow SK, Lim PL, Ho TC, Ip WK, Wong CK, Lam CW, Lee KK, Chan JY, Chui YL. A death receptor-associated anti-apoptotic protein, BRE, inhibits mitochondrial apoptotic pathway. *J Biol Chem* **279**:52106-52116 (2004). doi: 10.1074/jbc.M408678200.

Li MG, Liu XY, Liu ZQ, Hong JY, Liu JQ, Zhou CJ, Hu TY, Xiao XJ, Ran PX, Zheng PY, Liu ZG, Yang PC. Bcl2L12 Contributes to Th2-Biased Inflammation in the Intestinal Mucosa by Regulating $CD4^+$ T Cell Activities. *J Immunol* **201**:725-733 (2018). doi: 10.4049/jimmunol.1800139.

Li J, Chen Z, Kim G, Luo J, Hori S, Wu C. Cathepsin W restrains peripheral regulatory T cells for mucosal immune quiescence. *Sci Adv* **9**:eadf3924 (2023). doi: 10.1126/sciadv.adf3924

Li T, Wen Y, Lu Q, Hua S, Hou Y, Du X, Zheng Y, Sun S. MST1/2 in inflammation and immunity. *Cell Adh Migr* **17**:1-15 (2023). doi: 10.1080/19336918.2023.2276616.

Li S, Zhuge A, Chen H, Han S, Shen J, Wang K, Xia J, Xia H, Jiang S, Wu Y, Li L. Sedanolide alleviates DSS-induced colitis by modulating the intestinal FXR-SMPD3 pathway in mice. *J Adv Res* S2090-1232(24)00128-0 (2024). doi: 10.1016/j.jare.2024.03.026.

Liang J, Nagahashi M, Kim EY, Harikumar KB, Yamada A, Huang WC, Hait NC, Allegood JC, Price MM, Avni D, Takabe K, Kordula T, Milstien S, Spiegel S. Sphingosine-1-phosphate links persistent STAT3 activation, chronic intestinal inflammation, and development of colitis-associated cancer. *Cancer Cell* **23**:107-120 (2013). doi: 10.1016/j.ccr.2012.11.013.

Liao FH, Shui JW, Hsing EW, Hsiao WY, Lin YC, Chan YC, Tan TH, Huang CY. Protein phosphatase 4 is an essential positive regulator for Treg development, function, and protective gut immunity. *Cell Biosci* **4**:25 (2014). doi: 10.1186/2045-3701-4-25.

Liao X, Liu J, Guo X, Meng R, Zhang W, Zhou J, Xie X, Zhou H. Origin and Function of Monocytes in Inflammatory Bowel Disease. *J Inflamm Res* **17**:2897-2914 (2024) https://doi.org/10.2147/JIR.S450801

Lillehoj EP, Luzina IG, Atamas SP. Mammalian Neuraminidases in Immune-Mediated Diseases: Mucins and Beyond. *Front Immunol* **13**:883079 (2022). doi: 10.3389/fimmu.2022.883079.

Lin AE, Ebert G, Ow Y, Preston SP, Toe JG, Cooney JP, Scott HW, Sasaki M, Saibil SD, Dissanayake D, Kim RH, Wakeham A, You-Ten A, Shahinian A, Duncan G, Silvester J, Ohashi PS, Mak TW, Pellegrini M. ARIH2 is essential for embryogenesis, and its hematopoietic deficiency causes lethal activation of the immune system. *Nat Immunol* **14**:27-33 (2013). doi: 10.1038/ni.2478.

Lin N, Simon MC. Hypoxia-inducible factors: key regulators of myeloid cells during inflammation. *J Clin Invest* **126**:3661-3671 (2016). https://doi.org/10.1172/JCI84426.

Lin N, Shay JES, Xie H, Lee DSM, Skuli N, Tang Q, Zhou Z, Azzam A, Meng H, Wang H, FitzGerald GA, Simon MC. Myeloid Cell Hypoxia-Inducible Factors Promote Resolution of Inflammation in Experimental Colitis. *Front Immunol* **9**:2565 (2018). doi: 10.3389/fimmu.2018.02565.

Lin J, Chen K, Chen W, Yao Y, Ni S, Ye M, Zhuang G, Hu M, Gao J, Gao C, Liu Y, Yang M, Zhang Z, Zhang X, Huang J, Chen F, Sun L, Zhang X, Yu S, Chen Y, Jiang Y, Wang S, Yang X, Liu K, Zhou HM, Ji Z, Deng H, Haque ME, Li J, Mi LZ, Li Y, Yang Y. Paradoxical Mitophagy Regulation by PINK1 and TUFm. *Mol Cell* **80**:607-620.e12 (2020). doi: 10.1016/j.molcel.2020.10.007.

Liu L, Cara DC, Kaur J, Raharjo E, Mullaly SC, Jongstra-Bilen J, Jongstra J, Kubes P. LSP1 is an endothelial gatekeeper of leukocyte transendothelial migration. *J Exp Med* **201**:409-418 (2005). doi: 10.1084/jem.20040830.

Liu B, Gulati AS, Cantillana V, Henry SC, Schmidt EA, Daniell X, Grossniklaus E, Schoenborn AA, Sartor RB, Taylor GA. Irgm1-deficient mice exhibit Paneth cell abnormalities and increased susceptibility to acute intestinal inflammation. *Am J Physiol Gastrointest Liver Physiol* **305**:G573-84 (2013). doi: 10.1152/ajpgi.00071.2013.

Liu F, Li X, Yue H, Ji J, You M, Ding L, Fan H, Hou Y. TLR-Induced SMPD3 Defects Enhance Inflammatory Response of B Cell and Macrophage in the Pathogenesis of SLE. *Scand J Immunol* **86**:377-388 (2017). doi: 10.1111/sji.12611.

Liu, H., Zhu, Y., Gao, Y. *et al.* NR1D1 modulates synovial inflammation and bone destruction in rheumatoid arthritis. *Cell Death Dis* **11**: 129 (2020). https://doi.org/10.1038/s41419-020-2314-6

Liu E, Sun J, Yang J, Li L, Yang Q, Zeng J, Zhang J, Chen D, Sun Q. ZDHHC11 Positively Regulates NF-κB Activation by Enhancing TRAF6 Oligomerization. *Front Cell Dev Biol* **9**:710967 (2021). doi: 10.3389/fcell.2021.710967.

Liu T, Wang L, Liang P, Wang X, Liu Y, Cai J, She Y, Wang D, Wang Z, Guo Z, Bates S, Xia X, Huang J, Cui J. USP19 suppresses inflammation and promotes M2-like macrophage polarization by manipulating NLRP3 function via autophagy. *Cell Mol Immunol* **18**:2431-2442 (2021). doi: 10.1038/s41423-020-00567-7.

Liu Q, Zhu F, Liu X, Lu Y, Yao K, Tian N, Tong L, Figge DA, Wang X, Han Y, Li Y, Zhu Y, Hu L, Ji Y, Xu N, Li D, Gu X, Liang R, Gan G, Wu L, Zhang P, Xu T, Hu H, Hu Z, Xu H, Ye D, Yang H, Li B, Tong X. Non-oxidative pentose phosphate pathway controls regulatory T cell function by integrating metabolism and epigenetics. *Nat Metab* **4**:559-574 (2022). doi: 10.1038/s42255-022-00575-z.

Liu H, Zhen C, Xie J, Luo Z, Zeng L, Zhao G, Lu S, Zhuang H, Fan H, Li X, Liu Z, Lin S, Jiang H, Chen Y, Cheng J, Cao Z, Dai K, Shi J, Wang Z, Hu Y, Meng T, Zhou C, Han Z, Huang H, Zhou Q, He P, Feng D. TFAM is an autophagy receptor that limits inflammation by binding to cytoplasmic mitochondrial DNA. *Nat Cell Biol* **26**:878-891 (2024). doi: 10.1038/s41556-024-01419-6.

Logtenberg, M.E.W., Jansen, J.H.M., Raaben, M. *et al.* Glutaminyl cyclase is an enzymatic modifier of the CD47-SIRPα axis and a target for cancer immunotherapy. *Nat Med* **25**: 612–619 (2019). https://doi.org/10.1038/s41591-019-0356-z

Luo Y, de Lange K, Jostins L. *et al.* Exploring the genetic architecture of inflammatory bowel disease by whole-genome sequencing identifies association at *ADCY7*. *Nat Genet* **49**: 186–192 (2017). https://doi.org/10.1038/ng.3761.

Luo P, Yang Z, Chen B, Zhong X. The multifaceted role of CARD9 in inflammatory bowel disease. *J Cell Mol Med* **24**:34-39 (2020). doi: 10.1111/jcmm.14770.

Luthers CR, Dunn TM, Snow AL. ORMDL3 and Asthma: Linking Sphingolipid Regulation to Altered T Cell Function. *Front Immunol* **11**:597945 (2020). doi: 10.3389/fimmu.2020.597945.

Ma X, Meng Z, Jin L, Xiao Z, Wang X, Tsark WM, Ding L, Gu Y, Zhang J, Kim B, He M, Gan X, Shively JE, Yu H, Xu R, Huang W. CAMK2γ in intestinal epithelial cells modulates colitis-associated colorectal carcinogenesis via enhancing STAT3 activation. *Oncogene* **36**:4060-4071 (2017). doi: 10.1038/onc.2017.16.

Macho-Fernandez E, Koroleva EP, Spencer CM, Tighe M, Torrado E, Cooper AM, Fu YX, Tumanov AV. Lymphotoxin beta receptor signaling limits mucosal damage through driving IL-23 production by epithelial cells. *Mucosal Immunol* **8**:403-413 (2015). doi: 10.1038/mi.2014.78.

Maffucci P, Chavez J, Jurkiw TJ, O'Brien PJ, Abbott JK, Reynolds PR, Worth A, Notarangelo LD, Felgentreff K, Cortes P, Boisson B, Radigan L, Cobat A, Dinakar C, Ehlayel M, Ben-Omran T, Gelfand EW, Casanova JL, Cunningham-Rundles C. Biallelic mutations in DNA ligase 1 underlie a spectrum of immune deficiencies. *J Clin Invest* **128**:5489-5504 (2018) . doi: 10.1172/JCI99629.

Magg T, Shcherbina A, Arslan D, Desai MM, Wall S, Mitsialis V, Conca R, Unal E, Karacabey N, Mukhina A, Rodina Y, Taur PD, Illig D, Marquardt B, Hollizeck S, Jeske T, Gothe F, Schober T, Rohlfs M, Koletzko S, Lurz E, Muise AM, Snapper SB, Hauck F, Klein C, Kotlarz D. CARMIL2 Deficiency Presenting as Very Early Onset Inflammatory Bowel Disease. *Inflamm Bowel Dis* **25**:1788-1795 (2019). doi: 10.1093/ibd/izz103.

Makita S, Takatori H, Nakajima H. Post-Transcriptional Regulation of Immune Responses and Inflammatory Diseases by RNA-Binding ZFP36 Family Proteins. Front. Immunol 12 (2021). https://doi.org/10.3389/fimmu.2021.711633

Mandal M, Maienschein-Cline M, Maffucci P. *et al.* BRWD1 orchestrates epigenetic landscape of late B lymphopoiesis. *Nat Commun* **9**: 3888 (2018). https://doi.org/10.1038/s41467-018-06165-6

Martínez Gómez JM, Chen L, Schwarz H, Karrasch T. CD137 facilitates the resolution of acute DSS-induced colonic inflammation in mice. *PLoS One* **8**:e73277 (2013). doi: 10.1371/journal.pone.0073277.

Marzesco AM, Dunia I, Pandjaitan R, Recouvreur M, Dauzonne D, Benedetti EL, Louvard D, Zahraoui A. The small GTPase Rab13 regulates assembly of functional tight junctions in epithelial cells. *Mol Biol Cell* **13**:1819-1831 (2002). doi: 10.1091/mbc.02-02-0029.

Mashukova A, Wald FA, Salas PJ. Tumor necrosis factor alpha and inflammation disrupt the polarity complex in intestinal epithelial cells by a posttranslational mechanism. *Mol Cell Biol* **31**:756-765 (2011). doi: 10.1128/MCB.00811-10.

McRae MP. Coenzyme Q10 Supplementation in Reducing Inflammation: An Umbrella Review. *J Chiropr Med* **22**:131-137 (2023). doi: 10.1016/j.jcm.2022.07.001.

Melhem H, Spalinger MR, Cosin-Roger J, Atrott K, Lang S, Wojtal KA, Vavricka SR, Rogler G, Frey-Wagner I. Prdx6 Deficiency Ameliorates DSS Colitis: Relevance of Compensatory Antioxidant Mechanisms. *J Crohns Colitis* **11**:871-884 (2017). doi: 10.1093/ecco-jcc/jjx016.

Mencarelli A, Khameneh HJ, Fric J, Vacca M, El Daker S, Janela B, Tang JP, Nabti S, Balachander A, Lim TS, Ginhoux F, Ricciardi-Castagnoli P, Mortellaro A. Calcineurin-mediated IL-2 production by $CD11c^{high}MHCII^{+}$ myeloid cells is crucial for intestinal immune homeostasis. *Nat Commun* **9**:1102 (2018). doi: 10.1038/s41467-018-03495-3.

Menning M, Kufer TA. A role for the Ankyrin repeat containing protein Ankrd17 in Nod1- and Nod2-mediated inflammatory responses. *FEBS Lett* **587**:2137-2142 (2013). doi: 10.1016/j.febslet.2013.05.037.

Mercado-Lubo R. The interaction of gut microbes with host ABC transporters. *Gut Microbes* **1**: 301-306 (2010). https://doi.org/10.4161/gmic.1.5.12925

Meylan F, Song YJ, Fuss I, Villarreal S, Kahle E, Malm IJ, Acharya K, Ramos HL, Lo L, Mentink-Kane MM, Wynn TA, Migone TS, Strober W, Siegel RM. The TNF-family cytokine TL1A drives IL-13-dependent small intestinal inflammation. *Mucosal Immunol* **4**:172-185 (2011). doi: 10.1038/mi.2010.67.

Mi C, Wang Z, Li MY, Zhang ZH, Ma J, Jin X. Zinc finger protein 91 positively regulates the production of IL-1β in macrophages by activation of MAPKs and non-canonical caspase-8 inflammasome. *Br J Pharmacol* **175**:4338-4352 (2018). doi: 10.1111/bph.14493.

Mikuda N, Kolesnichenko M, Beaudette P, Popp O, Uyar B, Sun W, Tufan AB, Perder B, Akalin A, Chen W, Mertins P, Dittmar G, Hinz M, Scheidereit C. The IκB kinase complex is a regulator of mRNA stability. *EMBO J* **37**:e98658 (2018). doi: 10.15252/embj.201798658.

Miyahara, Y., Chen, H., Moriyama, M. *et al.* Toll-like receptor 9-positive plasmacytoid dendritic cells promote Th17 immune responses in oral lichen planus stimulated by epithelium-derived cathepsin K. *Sci Rep* **13**: 19320 (2023). https://doi.org/10.1038/s41598-023-46090-3

Mishra, V., Crespo-Puig, A., McCarthy, C. *et al.* IL-1β turnover by the UBE2L3 ubiquitin conjugating enzyme and HECT E3 ligases limits inflammation. *Nat Commun* **14**: 4385 (2023). https://doi.org/10.1038/s41467-023-40054-x

Mitsuyama K, Matsumoto S, Rose-John S, Suzuki A, Hara T, Tomiyasu N, Handa K, Tsuruta O, Funabashi H, Scheller J, Toyonaga A, Sata M. STAT3 activation via interleukin 6 trans-signalling contributes to ileitis in SAMP1/Yit mice. *Gut* **55**:1263-1269 (2006). doi: 10.1136/gut.2005.079343.

Mohanan V, Nakata T, Desch AN, Lévesque C, Boroughs A, Guzman G, Cao Z, Creasey E, Yao J, Boucher G, Charron G, Bhan AK, Schenone M, Carr SA, Reinecker HC, Daly MJ, Rioux JD, Lassen KG, Xavier RJ. *C1orf106* is a colitis risk gene that regulates stability of epithelial adherens junctions. *Science* **359**:1161-1166 (2018). doi: 10.1126/science.aan0814.

Mota AC, Dominguez M, Weigert A, Snodgrass RG, Namgaladze D, Brüne B. Lysosome-Dependent LXR and PPARδ Activation Upon Efferocytosis in Human Macrophages. *Front Immunol* **12**:637778 (2021). doi: 10.3389/fimmu.2021.637778.

Mukherjee S, Kumar R, Tsakem Lenou E, Basrur V, Kontoyiannis DL, Ioakeimidis F, Mosialos G, Theiss AL, Flavell RA, Venuprasad K. Deubiquitination of NLRP6 inflammasome by Cyld critically regulates intestinal inflammation. *Nat Immunol* 21:626-635 (2020). doi: 10.1038/s41590-020-0681-x.

Murata Y, Kotani T, Supriatna Y, Kitamura Y, Imada S, Kawahara K, Nishio M, Daniwijaya EW, Sadakata H, Kusakari S, Mori M, Kanazawa Y, Saito Y, Okawa K, Takeda-Morishita M, Okazawa H, Ohnishi H, Azuma T, Suzuki A, Matozaki T. Protein tyrosine phosphatase SAP-1 protects against colitis through regulation of CEACAM20 in the intestinal epithelium. *Proc Natl Acad Sci USA* **112**:E4264-4271 (2015). doi: 10.1073/pnas.1510167112.

Naydenov NG, Lechuga S, Zalavadia A, Mukherjee PK, Gordon IO, Skvasik D, Vidovic P, Huang E, Rieder F, Ivanov AI. P-Cadherin Regulates Intestinal Epithelial Cell Migration and Mucosal Repair, but Is Dispensable for Colitis Associated Colon Cancer. *Cells* **11**:1467 (2022). doi: 10.3390/cells11091467.

Ni H, Chen Y, Xia W, Wang C, Hu C, Sun L, Tang W, Cui H, Shen T, Liu Y, Li J. SATB2 Defect Promotes Colitis and Colitis-associated Colorectal Cancer by Impairing Cl-/HCO3- Exchange and Homeostasis of Gut Microbiota. *J Crohns Colitis* **15**:2088-2102 (2021). doi: 10.1093/ecco-jcc/jjab094.

Ntunzwenimana, J.C., Boucher, G., Paquette, J. *et al.* Functional screen of inflammatory bowel disease genes reveals key epithelial functions. *Genome Med* **13**: 181 (2021). https://doi.org/10.1186/s13073-021-00996-7

Oceandy D, Amanda B, Ashari FY, Faizah Z, Azis MA, Stafford N. The Cross-Talk Between the TNF-α and RASSF-Hippo Signalling Pathways. *Int J Mol Sci* **20**:2346 (2019). doi: 10.3390/ijms20092346.

Oertel S, Scholich K, Weigert A, Thomas D, Schmetzer J, Trautmann S, Wegner MS, Radeke HH, Filmann N, Brüne B, Geisslinger G, Tegeder I, Grösch S. Ceramide synthase 2 deficiency aggravates AOM-DSS-induced colitis in mice: role of colon barrier integrity. *Cell Mol Life Sci* **74**:3039-3055 (2017). doi: 10.1007/s00018-017-2518-9.

Ohira M, Oshitani N, Hosomi S, Watanabe K, Yamagami H, Tominaga K, Watanabe T, Fujiwara Y, Maeda K, Hirakawa K, Arakawa T. Dislocation of Rab13 and vasodilator-stimulated phosphoprotein in inactive colon epithelium in patients with Crohn's disease. *Int J Mol Med* **24**:829-835 (2009). doi: 10.3892/ijmm_00000300.

Ohl K, Winer A, Lippe R, Schippers A, Zorn C, Roth J, Wagner N, Tenbrock K. CREM alpha enhances IL-21 production in T cells in vivo and inb vitre. *Front Immunol* **7**: (2016) https://doi.org/10.3389/fimmu.2016.00618

Okabayashi S, Kobayashi T, Saito E, Toyonaga T, Ozaki R, Sagami S, Nakano M, Tanaka J, Yagisawa K, Kuronuma S, Takeuchi O, Hibi T. Individualized treatment based on CYP3A5 single-nucleotide polymorphisms with tacrolimus in ulcerative colitis. *Intest Res* **17**:218-226 (2019). doi: 10.5217/ir.2018.00117.

Olivera A, Urtz N, Mizugishi K, Yamashita Y, Gilfillan AM, Furumoto Y, Gu H, Proia RL, Baumruker T, Rivera J. IgE-dependent activation of sphingosine kinases 1 and 2 and secretion of sphingosine 1-phosphate requires Fyn kinase and contributes to mast cell response. *Mech Signal Transd* **281**: 2515-2525 (2006). https://doi.org/10.1074/jbc.M508931200

Paladini F, Fiorillo MT, Tedeschi V, Mattorre B, Sorrentino R. The Multifaceted Nature of Aminopeptidases ERAP1, ERAP2, and LNPEP: From Evolution to Disease. *Front Immunol* **11**:1576 (2020). doi: 10.3389/fimmu.2020.01576.

Palazon A, Goldrath AW, Nizet V, Johnson RS. HIF transcription factors, inflammation, and immunity. *Immunity* **41**:518-528 (2014). doi: 10.1016/j.immuni.2014.09.008.

Pandey SP, Yan J, Turner JR, Abraham C. Reducing IRF5 expression attenuates colitis in mice, but impairs the clearance of intestinal pathogens. *Mucosal Immunol* **12**:874-887 (2019). doi: 10.1038/s41385-019-0165-1.

Park I, Son M, Ahn E, Kim YW, Kong YY, Yun Y. The Transmembrane Adaptor Protein LIME Is Essential for Chemokine-Mediated Migration of Effector T Cells to Inflammatory Sites. *Mol Cells* **43**:921-934 (2020). doi: 10.14348/molcells.2020.0124.

Park SE, Lee D, Jeong JW, Lee SH, Park SJ, Ryu J, Oh SK, Yang H, Fang S, Kim S. Gut Epithelial Inositol Polyphosphate Multikinase Alleviates Experimental Colitis via Governing Tuft Cell Homeostasis. *Cell Mol Gastroenterol Hepatol* **14**:1235-1256 (2022). doi: 10.1016/j.jcmgh.2022.08.004.

Patkunarajah A, Stear JH, Moroni M, Schroeter L, Blaszkiewicz J, Tearle JL, Cox CD, Fürst C, Sánchez-Carranza O, Ocaña Fernández MDÁ, Fleischer R, Eravci M, Weise C, Martinac B, Biro M, Lewin GR, Poole K. TMEM87a/Elkin1, a component of a novel mechanoelectrical transduction pathway, modulates melanoma adhesion and migration. *Elife* **9**:e53308 (2020). doi: 10.7554/eLife.53308.

Pauls SD, Marshall AJ. Regulation of immune cell signaling by SHIP1: A phosphatase, scaffold protein, and potential therapeutic target. *Eur J Immunol* **47**:932-945 (2017). doi: 10.1002/eji.201646795.

Pei, W., Tanaka, K., Huang, S. *et al.* Extracellular HSP60 triggers tissue regeneration and wound healing by regulating inflammation and cell proliferation.*npj Regen Med* **1**: 16013 (2016). https://doi.org/10.1038/npjregenmed.2016.13

Petrey AC, Obery DR, Kessler SP, Zawerton A, Flamion B, de la Motte CA. Platelet hyaluronidase-2 regulates the early stages of inflammatory disease in colitis. *Blood* **134**:765-775 (2019). doi: 10.1182/blood.2018893594.

Pick R, Begandt D, Stocker TJ, Salvermoser M, Thome S, Böttcher RT, Montanez E, Harrison U, Forné I, Khandoga AG, Coletti R, Weckbach LT, Brechtefeld D, Haas R, Imhof A, Massberg S, Sperandio M, Walzog B. Coronin 1A, a novel player in integrin biology, controls neutrophil trafficking in innate immunity. *Blood* **130**:847-858 (2017). doi: 10.1182/blood-2016-11-749622.

Plant T, Eamsamarng S, Sanchez-Garcia MA, Reyes L, Renshaw SA, Coelho P, Mirchandani AS, Morgan JM, Ellett FE, Morrison T, Humphries D, Watts ER, Murphy F, Raffo-Iraolagoitia XL, Zhang A, Cash JL, Loynes C, Elks PM, Van Eeden F, Carlin LM, Furley AJ, Whyte MK, Walmsley SR. Semaphorin 3F signaling actively retains neutrophils at sites of inflammation. *J Clin Invest* **130**:3221-3237 (2020). doi: 10.1172/JCI130834.

Pochini L, Galluccio M, Console L, Scalise M, Eberini I, Indiveri C. Inflammation and Organic Cation Transporters Novel (OCTNs). *Biomolecules* **14**:392 (2024). doi: 10.3390/biom14040392.

Podolsky DK, Lobb R, King N, Benjamin CD, Pepinsky B, Sehgal P, deBeaumont M. Attenuation of colitis in the cotton-top tamarin by anti-alpha 4 integrin monoclonal antibody. *J Clin Invest* 92:372-380 (1993). doi: 10.1172/JCI116575.

Popović B, Nicolet BP, Guislain A, Engels S, Jurgens AP, Paravinja N, Freen-van Heeren JJ, van Alphen FPJ, van den Biggelaar M, Salerno F, Wolkers MC. Time-dependent regulation of cytokine production by RNA binding proteins defines T cell effector function. *Cell Rep* **42**:112419 (2023). doi: 10.1016/j.celrep.2023.112419.

Prasad M, Brzostek J, Gautam N, Balyan R, Rybakin V, Gascoigne NRJ. Themis regulates metabolic signaling and effector functions in $CD4^+$ T cells by controlling NFAT nuclear translocation. *Cell Mol Immunol* **18**:2249-2261 (2021). doi: 10.1038/s41423-020-00578-4.

Pravoverov K, Fatima I, Barman S, Jühling F, Primeaux M, Baumert TF, Singh AB, Dhawan P. IL-22 regulates MASTL expression in intestinal epithelial cells. Am J Physiol Gastrointest Liver Physiol 327: G123-G139 (2024). DOI: 10.1152/ajpgi.00260.2023.

Prieto S, Dubra G, Camasses A, Aznar AB, Begon_Pescia C, Simboeck E, Pirot N, Gerbe F, Angevin L, Jay P, Krasinska L, Fisher D. CDK8 and CDK19 act redundantly to control the CFTR pathway in the intestinal epithelium. *EMBO rep* **24**: e54261 (2022). https://doi.org/10.15252/embr.202154261

Qiu W, Wu B, Wang X, Buchanan ME, Regueiro MD, Hartman DJ, Schoen RE, Yu J, Zhang L. PUMA-mediated intestinal epithelial apoptosis contributes to ulcerative colitis in humans and mice. *J Clin Invest* **121**:1722-1732 (2011). doi: 10.1172/JCI42917.

Ranganathan P, Jayakumar C, Manicassamy S, Ramesh G. CXCR2 knockout mice are protected against DSS-colitis-induced acute kidney injury and inflammation. *Am J Physiol Renal Physiol* **305**:F1422-7 (2013). doi: 10.1152/ajprenal.00319.2013.

Read, K.A., Jones, D.M., Pokhrel, S. *et al.* Aiolos represses CD4⁺ T cell cytotoxic programming via reciprocal regulation of $T_{FH}$ transcription factors and IL-2 sensitivity. *Nat Commun* **14**: 1652 (2023). https://doi.org/10.1038/s41467-023-37420-0

Reith W, Mach B. The bare lymphocyte syndrome and the regulation of MHC expression. *Ann Rev Immunol* **19**:331-373 (2001). https://doi.org/10.1146/annurev.immunol.19.1.331

Ren F, Geng Y, Minami T, Qiu Y, Feng Y, Liu C, Zhao J, Wang Y, Fan X, Wang Y, Li M, Li J, Chang Z. Nuclear termination of STAT3 signaling through SIPAR (STAT3-Interacting Protein As a Repressor)-dependent recruitment of T cell tyrosine phosphatase TC-PTP. *FEBS Lett* **589**:1890-1896 (2015). doi: 10.1016/j.febslet.2015.05.031.

Renoux F, Stellato M, Haftmann C, Vogetseder A, Huang R, Subramaniam A, Becker MO, Blyszczuk P, Becher B, Distler JHW, Kania G, Boyman O, Distler O. The AP1 Transcription Factor Fosl2 Promotes Systemic Autoimmunity and Inflammation by Repressing Treg Development. *Cell Reports* **31**: 107826 (2020). https://doi.org/10.1016/j.celrep.2020.107826.

Reyat JS, Chimen M, Noy PJ, Szyroka J, Rainger GE, Tomlinson MG. ADAM10-Interacting Tetraspanins Tspan5 and Tspan17 Regulate VE-Cadherin Expression and Promote T Lymphocyte Transmigration. *J Immunol* **199**:666-676 (2017). doi: 10.4049/jimmunol.1600713.

Roncagalli R, Mingueneau M, Grégoire C, Malissen M, Malissen B. LAT signaling pathology: an "autoimmune" condition without T cell self-reactivity. *Trends Immunol* **31**:253-259 (2010). doi: 10.1016/j.it.2010.05.001.

Roncagalli R, Cucchetti M, Jarmuzynski N, Grégoire C, Bergot E, Audebert S, Baudelet E, Menoita MG, Joachim A, Durand S, Suchanek M, Fiore F, Zhang L, Liang Y, Camoin L, Malissen M, Malissen B. The scaffolding function of the RLTPR protein explains its essential role for CD28 co-stimulation in mouse and human T cells. *J Exp Med* **213**:2437-2457 (2016). doi: 10.1084/jem.20160579.

Rose-John, S., Jenkins, B.J., Garbers, C. *et al.* Targeting IL-6 trans-signalling: past, present and future prospects. *Nat Rev Immunol* **23**: 666–681 (2023). https://doi.org/10.1038/s41577-023-00856-y

Sabui S, Bohl JA, Kapadia R, Cogburn K, Ghosal A, Lambrecht NW, Said HM. Role of the sodium-dependent multivitamin transporter (SMVT) in the maintenance of intestinal mucosal integrity. *Am J Physiol Gastrointest Liver Physiol* **311**:G561-570 (2016). doi: 10.1152/ajpgi.00240.2016.

Salcedo R, Worschech A, Cardone M, Jones Y, Gyulai Z, Dai RM, Wang E, Ma W, Haines D, O'hUigin C, Marincola FM, Trinchieri G. MyD88-mediated signaling prevents development of adenocarcinomas of the colon: role of interleukin 18. *J Exp Med* **207**:1625-1636 (2010). doi: 10.1084/jem.20100199.

Sandner L, Alteneder M, Zhu C, Hladik A, Högler S, Rica R, Van Greuningen LW., Sharif O, Sakaguchi S, Knapp S, Kenner L, Trauner M, Ellmeier W, Boucheron N. The Tyrosine Kinase Tec Regulates Effector Th17 Differentiation, Pathogenicity, and Plasticity in T-Cell-Driven Intestinal Inflammation. *Front Immunol* **12**: (2021). https://www.frontiersin.org/journals/immunology/articles/10.3389/fimmu.2021.750466

Seto S, Tsujimura K, Koide Y. Coronin-1a inhibits autophagosome formation around Mycobacterium tuberculosis-containing phagosomes and assists mycobacterial survival in macrophages. *Cell Microbiol* **14**:710-727 (2012). doi: 10.1111/j.1462-5822.2012.01754.x.

Seto E, Yoshida-Sugitani R, Kobayashi T, Toyama-Sorimachi N. The Assembly of EDC4 and Dcp1a into Processing Bodies Is Critical for the Translational Regulation of IL-6. PLoS One **10**:e0123223 (2015). doi: 10.1371/journal.pone.0123223.

Sharma J, Khan S, Singh NC, Sahu S, Raj D, Prakash S, Bandyopadhyah P, Srkar K, Bhosale V, Chandra T, kumaravelu J, Barthwal MK, Gupta SK, Srivastava M, Guha R, Ammanathan V, Ghoshal UC, Mitra K, Lahiri A. ORMDL3 regulates NLRP3 inflammasome activation by maintaining ER-mitochondria contacts in human macrophages and dictates ulcerative colitis patient outcome. *J Biol Chem* **300**: 107120 (2024). https://doi.org/10.1016/j.jbc.2024.107120

Shen H, Campanello GC, Flicker D, Grabarek Z, Hu J, Luo C, Banerjee R, Mootha VK. The Human Knockout Gene CLYBL Connects Itaconate to Vitamin $B_{12}$. *Cell* **171**:771-782.e11 (2017). doi: 10.1016/j.cell.2017.09.051.

Shi C, Zhang X, Chen Z, Sulaiman K, Feinberg MW, Ballantyne CM, Jain MK, Simon DI. Integrin engagement regulates monocyte differentiation through the forkhead transcription factor Foxp1. *J Clin Invest* **114**:408-418 (2004). doi: 10.1172/JCI21100.

Shi H, Wang Y, Li X, Zhan X, Tang M, Fina M, Su L, Pratt D, Bu CH, Hildebrand S, Lyon S, Scott L, Quan J, Sun Q, Russell J, Arnett S, Jurek P, Chen D, Kravchenko VV, Mathison JC, Moresco EM, Monson NL, Ulevitch RJ, Beutler B. NLRP3 activation and mitosis are mutually exclusive events coordinated by NEK7, a new inflammasome component. *Nat Immunol* **17**:250-258 (2016). doi: 10.1038/ni.3333.

Shon WJ, Song JW, Oh SH *et al.* Gut taste receptor type 1 member 3 is an intrinsic regulator of Western diet-induced intestinal inflammation. *BMC Med* **21**: 165 (2023). https://doi.org/10.1186/s12916-023-02848-0

Sidwell, T., Liao, Y., Garnham, A.L. *et al.* Attenuation of TCR-induced transcription by Bach2 controls regulatory T cell differentiation and homeostasis. *Nat Commun* **11**: 252 (2020). https://doi.org/10.1038/s41467-019-14112-2

Silvera-Ruiz SM, Gemperle C, Peano N, Olivero V, Becerra A, Häberle J, Gruppi A, Larovere LE, Motrich RD. Immune Alterations in a Patient With Hyperornithinemia-Hyperammonemia-Homocitrullinuria Syndrome: A Case Report. *Front Immunol* **13**:861516 (2022). doi: 10.3389/fimmu.2022.861516.

Sina C, Lipinski S, Gavrilova O, Aden K, Rehman A, Till A, Rittger A, Podschun R, Meyer-Hoffert U, Haesler R, Midtling E, Pütsep K, McGuckin MA, Schreiber S, Saftig P, Rosenstiel P. Extracellular cathepsin K exerts antimicrobial activity and is protective against chronic intestinal inflammation in mice. *Gut* **62**:520-530 (2013). doi: 10.1136/gutjnl-2011-300076.

Sinha, S., Bheemsetty, V.A. & Inamdar, M.S. A double helical motif in OCIAD2 is essential for its localization, interactions and STAT3 activation. *Sci Rep* **8**: 7362 (2018). https://doi.org/10.1038/s41598-018-25667-3

Smyth P, Sasiwachirangkul J, Williams R, Scott CJ. Cathepsin S (CTSS) activity in health and disease - A treasure trove of untapped clinical potential. *Mol Aspects Med* **88**:101106 (2022). doi: 10.1016/j.mam.2022.101106.

Sommer J, Engelowski E, Baran P, Garbers C, Floss DM, Scheller J. Interleukin-6, but not the interleukin-6 receptor plays a role in recovery from dextran sodium sulfate-induced colitis. *Int J Mol Med* **34**:651-660 (2014). doi: 10.3892/ijmm.2014.1825.

Song S, He X, Wang J, Wang R, Wang L, Zhao W, Wang Y, Zhang Y, Yu Z, Miao D, Xue Y. ELF3-AS1 contributes to gastric cancer progression by binding to hnRNPK and induces thrombocytosis in peripheral blood. *Cancer Sci* **112**:4553-4569 (2021). doi: 10.1111/cas.15104.

Sorbara MT, Ellison LK, Ramjeet M, Travassos LH, Jones NL, Girardin SE, Philpott DJ. The protein ATG16L1 suppresses inflammatory cytokines induced by the intracellular sensors Nod1 and Nod2 in an autophagy-independent manner. *Immunity* **39**:858-873 (2013). doi: 10.1016/j.immuni.2013.10.013.

Souza-Costa LP, Andrade-Chaves JT, Andrade JM, Costa VV, Franco LH. Uncovering new insights into the role of the ubiquitin ligase Smurf1 on the regulation of innate immune signaling and resistance to infection. *Front Immunol* **14**:1185741 (2023). doi: 10.3389/fimmu.2023.1185741.

Spalinger MR, Manzini R, Hering L, Riggs JB, Gottier C, Lang S, Atrott K, Fettelschoss A, Olomski F, Kündig TM, Fried M, McCole DF, Rogler G, Scharl M. PTPN2 regulates inflammasome activation and controls onset of intestinal inflammation and colon cancer. *Cell Rep* **22**:1835-1848 (2018).

Spalinger M, Sanchez Alvarez R, Gottier C, Montalban-Arques A, Schwarzfischer M, Niechcial A, Scharl M. Loss of PTPN23 in the intestinal epithelium results in epithelial hyperproliferation and lethal diarrhea in a microbiota dependent manner, *Journal of Crohn's and Colitis* **16**: i099 (2022). https://doi.org/10.1093/ecco-jcc/jjab232.090

Stankey, C.T., Bourges, C., Haag, L.M. *et al.* A disease-associated gene desert directs macrophage inflammation through ETS2. *Nature* **630**: 447–456 (2024). https://doi.org/10.1038/s41586-024-07501-1

Stegen M, Frey UH. The Role of G Protein-Coupled Receptor Kinase 6 Regulation in Inflammation and Pain. *Int J Mol Sci* **23**:15880 (2022). doi: 10.3390/ijms232415880.

Storey JD, Tibshirani R. Statistical significance for genome wide studies. *Proc. Natl. Acad. Sc. US* **100**: 9440-9445 (2003).

Subramanian VS, Marchant JS, Boulware MJ, Ma TY, Said HM. Membrane targeting and intracellular trafficking of the human sodium-dependent multivitamin transporter in polarized epithelial cells. *Am J Physiol Cell Physiol* **296**:C663-671 (2009). doi: 10.1152/ajpcell.00396.2008.

Sukocheva OA, Lukina E, McGowan E, Bishayee A. Sphingolipids as mediators of inflammation and novel therapeutic target in inflammatory bowel disease. *Adv Protein Chem Struct Biol* **120**:123-158 (2020). doi: 10.1016/bs.apcsb.2019.11.003.

Sun X, Liu T, Zhao J, Xia H, Xie J, Guo Y, Zhong L, Li M, Yang Q, Peng C, Rouvet I, Belot A, Shu HB, Feng P, Zhang J. DNA-PK deficiency potentiates cGAS-mediated antiviral innate immunity. *Nat Commun* **11:**6182 (2020). doi: 10.1038/s41467-020-19941-0.

Sun R, Lim S-O. FBXL20-mediated ubiquitination triggers the proteasomal degradation of 4-1BB. *FEBS J* **289**: 4549-4563 (2020). https://doi.org/10.1111/febs.16383

Takagi H, Kanai T, Okazawa A, Kishi Y, Sato T, Takaishi H, Inoue N, Ogata H, Iwao Y, Hoshino K, Takeda K, Akira S, Watanabe M, Ishii H, Hibi T. Contrasting action of IL-12 and IL-18 in the development of dextran sodium sulphate colitis in mice. *Scand J Gastroenterol* **38**:837-844 (2003). doi: 10.1080/00365520310004047.

Takedatsu H, Michelsen KS, Wei B, Landers CJ, Thomas LS, Dhall D, Braun J, Targan SR. TL1A (TNFSF15) regulates the development of chronic colitis by modulating both T-helper 1 and T-helper 17 activation. *Gastroenterology* **135**:552-567 (2008). doi: 10.1053/j.gastro.2008.04.037.

Tamehiro N, Nishida K, Yanobu-Takanashi R, Goto M, Okamura T, Suzuki H. T-cell activation RhoGTPase-activating protein plays an important role in $T_H$17-cell differentiation. *Immunol Cell Biol* **95**:729-735 (2017) . doi: 10.1038/icb.2017.27.

Taniguchi K, Inoue M, Arai K, Uchida K, Migita O, Akemoto Y, Hirayama J, Takeuchi I, Shimizu H, Hata K. Novel TNFAIP3 microdeletion in a girl with infantile-onset inflammatory bowel disease complicated by a severe perianal lesion. *Hum Genome Var* **8**:1 (2021). doi: 10.1038/s41439-020-00128-4.

Tatari MN, De Craene B, Soen B, Taminau J, Vermassen P, Goossens S, Haigh K, Cazzola S, Lambert J, Huylebroeck D, Haigh JJ, Berx G. ZEB2-transgene expression in the epidermis compromises the integrity of the epidermal barrier through the repression of different tight junction proteins. *Cell Mol Life Sci* **71**:3599-3609 (2014). doi: 10.1007/s00018-014-1589-0.

Tatematsu M, Yoshida R, Morioka Y, Ishii N, Funami K, Watanabe A, Saeki K, Seya T, Matsumoto M. Raftlin Controls Lipopolysaccharide-Induced TLR4 Internalization and TICAM-1 Signaling in a Cell Type–Specific Manner. *J Immunol* **196**: 3865–3876 (2016). https://doi.org/10.4049/jimmunol.1501734

Tian N, Hu L, Lu Y, Tong L, Feng M, Liu Q, Li Y, Zhu Y, Wu L, Ji Y, Zhang P, Xu T, Tong X. TKT maintains intestinal ATP production and inhibits apoptosis-induced colitis. *Cell Death Dis* **12**:853 (2021). doi: 10.1038/s41419-021-04142-4.

Tosi F, Sartori F, Guarini P, Olivieri O, Martinelli N. Delta-5 and delta-6 desaturases: crucial enzymes in polyunsaturated fatty acid-related pathways with pleiotropic influences in health and disease. *Adv Exp Med Biol* **824**:61-81 (2014). doi: 10.1007/978-3-319-07320-0_7.

Tu Q, Yu X, Xie W, Luo Y, Tang H, Chen K, Ruan Y, Li Y, Zhou J, Yin Y, Chen D, Song Z. Prokineticin 2 promotes macrophages-mediated antibacterial host defense against bacterial pneumonia. *Int J Inf Dis* **12**: 103-113 (2022). https://doi.org/10.1016/j.ijid.2022.10.003.

Tumbarello DA, Waxse BJ, Arden SD, Bright NA, Kendrick-Jones J, Buss F. Autophagy receptors link myosin VI to autophagosomes to mediate Tom1-dependent autophagosome maturation and fusion with the lysosome. *Nat Cell Biol* **14**:1024-1035 (2012). doi: 10.1038/ncb2589.

Uematsu A, Kido K, Takahashi H, Takahashi C, Yanagihara Y, Saeki N, Yoshida S, Maekawa M, Honda M, Kai T, Shimizu K, Higashiyama S, Imai Y, Tokunaga F, Sawasaki T. The E3 ubiquitin ligase MIB2 enhances inflammation by degrading the deubiquitinating enzyme CYLD. *J Biol Chem* **294**:14135-14148 (2019). doi: 10.1074/jbc.RA119.010119.

Valdez Y, Grassl GA, Guttman JA, Coburn B, Gros P, Vallance BA, Finlay BB. Nramp1 drives an accelerated inflammatory response during Salmonella-induced colitis in mice. *Cell Microbiol* **11**:351-362 (2009). doi: 10.1111/j.1462-5822.2008.01258.x.

van Driel B, Liao G, Romero X, O'Keeffe MS, Wang G, Faubion WA, Berger SB, Magelky EM, Manocha M, Azcutia V, Grisham M, Luscinskas FW, Mizoguchi E, de Waal Malefyt R, Reinecker HC, Bhan AK, Wang N, Terhorst C.

Signaling lymphocyte activation molecule regulates development of colitis in mice. *Gastroenterology* **143**:1544-1554.e7 (2012). doi: 10.1053/j.gastro.2012.08.042.

Varona R, Cadenas V, Flores J, Martínez-A C, Márquez G. CCR6 has a non-redundant role in the development of inflammatory bowel disease. *Eur J Immunol* **33**:2937-2946 (2003). doi: 10.1002/eji.200324347.

Verjans E., Ohl K., Reiss L. K., van Wijk F., Toncheva A. A., Wiener A., Yu Y., Rieg A. D., Gaertner V. D., Roth J., Knol E., Kabesch M., Wagner N., et al The cAMP response element modulator (CREM) regulates $T_H2$ mediated inflammation. *Oncotarget* **6**: 38538-38551 (2015). https://doi.org/10.18632/oncotarget.6041

Wang S, Yang YK, Chen T, Zhang H, Yang WW, Song SS, Zhai ZH, Chen DY. RNF123 has an E3 ligase-independent function in RIG-I-like receptor-mediated antiviral signaling. *EMBO Rep* **17**:1155-1168 (2016). doi: 10.15252/embr.201541703.

Wang, R., Li, H., Wu, J. *et al.* Gut stem cell necroptosis by genome instability triggers bowel inflammation. *Nature* **580**: 386–390 (2020). https://doi.org/10.1038/s41586-020-2127-x

Wang J, Wang W, Wang H, Tuo B. Physiological and Pathological Functions of SLC26A6. *Front Med (Lausanne)*. **7**:618256 (2021). doi: 10.3389/fmed.2020.618256.

Wang C, Li X, Xue B, Yu C, Wang L, Deng R, Liu H, Chen Z, Zhang Y, Fan S, Zuo C, Sun H, Zhu H, Wang J, Tang S. RasGRP1 promotes the acute inflammatory response and restricts inflammation-associated cancer cell growth. *Nat Commun* **13**:7001 (2022). doi: 10.1038/s41467-022-34659-x.

Watanabe R, Fujimoto M, Ishiura N, Kuwano Y, Nakashima H, Yazawa N, Okochi H, Sato S, Tedder TF, Tamaki K. CD19 expression in B cells is important for suppression of contact hypersensitivity. *Am J Pathol* **171**:560-570 (2007). doi: 10.2353/ajpath.2007.061279.

Wei Y, Ren X, Galbo PM Jr, Moerdler S, Wang H, Sica RA, Etemad-Gilbertson B, Shi L, Zhu L, Tang X, Lin Q, Peng M, Guan F, Zheng D, Chinai JM, Zang X. KIR3DL3-HHLA2 is a human immunosuppressive pathway and a therapeutic target. *Sci Immunol* **6**:eabf9792 (2021). doi: 10.1126/sciimmunol.abf9792.

Wei L, Zhang R, Zhang J, Li J, Kong D, Wang Q, Fang J, Wang L. PRKAR2A deficiency protects mice from experimental colitis by increasing IFN-stimulated gene expression and modulating the intestinal microbiota. *Mucosal Immunol* **14**:1282-1294 (2021). doi: 10.1038/s41385-021-00426-2.

Welz L, Kim NM, Harris D, Alsaadi A, Oumari M, Credidio G, Taubenheim J, Tran F, L Sievers, K Volk V, Koncina E, Verstockt B, Kaleta C, Letellier E, Feuerhake F, McReynolds M, Rosenstiel P, Schreiber S, Aden K. P224 JAK-STAT-Driven Tryptophan Degradation Fuels Mucosal Inflammation through QPRT Suppression-Induced Quinolinic Acid Overflow, *Journal of Crohn's and Colitis* **18**: 560 (2024) https://doi.org/10.1093/ecco-jcc/jjad212.0354

Wenzek C, Boelen A, Westendorf AM, Engel DR, Moeller LC, Führer D. The interplay of thyroid hormones and the immune system – where we stand and why we need to know about it,.*European Journal of Endocrinology* **186**: R65–R77 (2022), https://doi.org/10.1530/EJE-21-1171

West NR, Hegazy AN, Owens BMJ, Bullers SJ, Linggi B, Buonocore S, Coccia M, Görtz D, This S, Stockenhuber K, Pott J, Friedrich M, Ryzhakov G, Baribaud F, Brodmerkel C, Cieluch C, Rahman N, Müller-Newen G, Owens RJ, Kühl AA, Maloy KJ, Plevy SE; Oxford IBD Cohort Investigators; Keshav S, Travis SPL, Powrie F. Oncostatin M drives intestinal inflammation and predicts response to tumor necrosis factor-neutralizing therapy in patients with inflammatory bowel disease. *Nat Med* **23**:579-589 (2017). doi: 10.1038/nm.4307.

Wilkinson S, Croft DR, O'Prey J, Meedendorp A, O'Prey M, Dufès C, Ryan KM. The cyclin-dependent kinase PITSLRE/CDK11 is required for successful autophagy. *Autophagy* **7**:1295-1301 (2011). doi: 10.4161/auto.7.11.16646.

Wilson CH, Crombie C, van der Weyden L, Poulogiannis G, Rust AG, Pardo M, Gracia T, Yu L, Choudhary J, Poulin GB, McIntyre RE, Winton DJ, March HN, Arends MJ, Fraser AG, Adams DJ. Nuclear receptor binding protein 1 regulates intestinal progenitor cell homeostasis and tumour formation. *EMBO J* **31**:2486-2497 (2012). https://doi.org/10.1038/emboj.2012.91

Wolf CL, Pruett C, Lighter D, Jorcyk CL. The clinical relevance of OSM in inflammatory diseases: a comprehensive review. *Front Immunol* **14** (2023). https://doi.org/10.3389/fimmu.2023.1239732

Won M, Ro H, Dawid IB. Lnx2 ubiquitin ligase is essential for exocrine cell differentiation in the early zebrafish pancreas. *Proc Natl Acad Sci USA* **112**:12426-12431 (2015). doi: 10.1073/pnas.1517033112.

Worthington JJ, Reimann F, Gribble FM. Enteroendocrine cells-sensory sentinels of the intestinal environment and orchestrators of mucosal immunity. *Mucosal Immunol* **11**:3-20 (2018). doi: 10.1038/mi.2017.73.

Wright AP, Harris S, Madden S, Ramirez Reyes B, Mulamula E, Gibson A, Raucg I, Constant DA, Nice TJ. Interferon regulatory factor 6 (IRF6) determines intestinal epithelial cell development and immunity. *Mucosal Immunol* **17**: 633-650 (2024). https://doi.org/10.1016/j.mucimm.2024.03.013

Wu H, Handley TNG, Hoare BL, Hartono HA, Scott DJ, Chalmers DK, Bathgate RAD, Hossain MA. Developing insulin-like peptide 5-based antagonists for the G protein-coupled receptor, RXFP4. *Biochemical Pharmacology* **224**:116239 (2024). https://doi.org/10.1016/j.bcp.2024.116239.

Wu Q, Liu Y, Liang J, Dai A, Du B, Xi X, Jin L, Guo Y. Baricitinib relieves DSS-induced ulcerative colitis in mice by suppressing the NF-κB and JAK2/STAT3 signalling pathways. *Inflammopharmacology* **32**:849-861 (2024). doi: 10.1007/s10787-023-01396-6.

Xia M, Liu J, Wu X, Liu S, Li G, Han C, Song L, Li Z, Wang Q, Wang J, Xu T, Cao X. Histone methyltransferase Ash1l suppresses interleukin-6 production and inflammatory autoimmune diseases by inducing the ubiquitin-editing enzyme A20. *Immunity* **39**:470-481 (2013). doi: 10.1016/j.immuni.2013.08.016.

Xia, M., Liu, J., Liu, S. *et al.* Ash1l and lnc-Smad3 coordinate *Smad3* locus accessibility to modulate iTreg polarization and T cell autoimmunity. *Nat Commun* **8**: 15818 (2017). https://doi.org/10.1038/ncomms15818

Xia Q, Zheng H, Li Y, Xu W, Wu C, Xu J, Li S, Zhang L, Dong L. SMURF1 controls the PPP3/calcineurin complex and TFEB at a regulatory node for lysosomal biogenesis. Autophagy **20**:735-751 (2024). doi: 10.1080/15548627.2023.2267413.

Xu P, Roes J, Segal AW, Radulovic M. The role of grancalcin in adhesion of neutrophils. *Cell Immunol* **240**: 116-121 (2006).

Xu H, Shi J, Gao H, Liu Y, Yang Z, Shao F, Dong N. The N-end rule ubiquitin ligase UBR2 mediates NLRP1B inflammasome activation by anthrax lethal toxin. *EMBO J* **38**:e101996 (2019). doi: 10.15252/embj.2019101996.

Xu, S., He, Y., Lin, L. *et al.* The emerging role of ferroptosis in intestinal disease.*Cell Death Dis* **12**: 289 (2021). https://doi.org/10.1038/s41419-021-03559-1

Xu, C., Gu, L., Hu, L. *et al.* FADS1-arachidonic acid axis enhances arachidonic acid metabolism by altering intestinal microecology in colorectal cancer. *Nat Commun* **14**: 2042 (2023). https://doi.org/10.1038/s41467-023-37590-x

Xu ZY, Wang JC. LACC1 regulates changes in the intestinal flora in a mouse model of inflammatory bowel disease. *BMC Gastroenterol* **23**:358 (2023). doi: 10.1186/s12876-023-02971-5.

Xu X, Han Y, Deng J, Wang S, Zhuo S, Zhao K, Zhou W. Repurposing disulfiram with CuET nanocrystals: Enhancing anti-pyroptotic effect through NLRP3 inflammasome inhibition for treating inflammatory bowel diseases. *Acta Pharm Sin B* **14**:2698-2715 (2024). doi: 10.1016/j.apsb.2024.03.003.

Xue X, Miao Y, Wei Z. Nicotinamide adenine dinucleotide metabolism: driving or counterbalancing inflammatory bowel disease? *FEBS L* 597: 1179-1192 (2023). https://doi.org/10.1002/1873-3468.14528

Yamamoto Y, Nakase H, Matsuura M, Maruyama S, Masuda S. *CYP3A5* Genotype as a Potential Pharmacodynamic Biomarker for Tacrolimus Therapy in Ulcerative Colitis in Japanese Patients. *Int J Mol Sci* **21**:4347 (2020). doi: 10.3390/ijms21124347.

Yamaoka T, Yan F, Cao H, Polk DB. Transcativation of EGF receptor and ERBB2 protects intestinal epithelial cells from TNF-induced apoptosis. Proc Natl Acad Sci USA 105:11772-11777 (2008). https://doi.org/10.1073/pnas.0801463105

Yan J, Hedl M, Abraham C. An inflammatory bowel disease-risk variant in INAVA decreases pattern recognition receptor-induced outcomes. *J Clin Invest* **127**:2192-2205 (2017). doi: 10.1172/JCI86282.

Yang J, Xu P, Han L, Guo Z, Wang X, Chen Z, Nie J, Yin S, Piccioni M, Tsun A, Lv L, Ge S, Li B. Cutting edge: Ubiquitin-specific protease 4 promotes Th17 cell function under inflammation by deubiquitinating and stabilizing RORγt. *J Immunol* **194**:4094-4097 (2015). doi: 10.4049/jimmunol.1401451.

Yang S, He X, Zhao J, Wang D, Guo S, Gao T, Wang G, Jin C, Yan Z, Wang N, Wang Y, Zhao Y, Xing J, Huang Q. Mitochondrial transcription factor A plays opposite roles in the initiation and progression of colitis-associated cancer. *Cancer Commun (Lond)* **41**:695-714 (2021). doi: 10.1002/cac2.12184.

Ye B, Liu B, Yang L, Huang G, Hao L, Xia P, Wang S, Du Y, Qin X, Zhu P, Wu J, Sakaguchi N, Zhang J, Fan Z. Suppression of SRCAP chromatin remodelling complex and restriction of lymphoid lineage commitment by Pcid2. *Nat Commun* **8**:1518 (2017). doi: 10.1038/s41467-017-01788-7.

Ye B, Yang L, Qian G, Liu B, Zhu X, Zhu P, Ma J, Xie W, Li H, Lu T, Wang Y, Wang S, Du Y, Wang Z, Jiang J, Li J, Fan D, Meng S, Wu J, Tian Y, Fan Z. The chromatin remodeler SRCAP promotes self-renewal of intestinal stem cells. *EMBO J* **39**:e103786 (2020). doi: 10.15252/embj.2019103786.

Yıldız Ç, Gezgin Yıldırım D, Inci A, Tümer L, Cengiz Ergin FB, Sunar Yayla ENS, Esmeray Şenol P, Karaçayır N, Eğritaş Gürkan Ö, Okur I, Ezgü FS, Bakkaloğlu SA. A possibly new autoinflammatory disease due to compound heterozygous phosphomevalonate kinase gene mutation. *Joint Bone Spine* **90**:105490 (2023°. doi: 10.1016/j.jbspin.2022.105490.

Yim WWY, Mizushima N. Lysosome biology in autophagy. *Cell Discov* **6**: 6 (2020). https://doi.org/10.1038/s41421-020-0141-7.

Yoshida K, Murayama MA, Shimizu K, Tang C, Katagiri N, Matsuo K, Fukai F, Iwakura Y. IL-1R2 deficiency suppresses dextran sodium sulfate-induced colitis in mice via regulation of microbiota. *Biochem Biophys Res Commun* **496**:934-940 (2018). doi: 10.1016/j.bbrc.2018.01.116.

Yu J, Liu Z, Liang Y, Luo F, Zhang J, Tian C, Motzik A, Zheng M, Kang J, Zhong G, Liu C, Fang P, Guo M, Razin E, Wang J. Second messenger $Ap_4A$ polymerizes target protein HINT1 to transduce signals in FcεRI-activated mast cells. *Nat Commun* **10**:4664 (2019). doi: 10.1038/s41467-019-12710-8.

Yu H, Liu Z. GNA12 regulates C5a-induced migration by downregulating C5aR1-PLCβ2-PI3K-AKT-ERK1/2 signaling. *Biophys Rep* **9**:33-44 (2023). doi: 10.52601/bpr.2023.230001.

Yu M, Zhang Q, Yuan K, Sazonovs A, Stevens C, IIBDGC, Anderson CA, Daly MJ, Huang H. Cystic fibrosis risk variants confer protective effects against inflammatory bowel disease in large-scale exome sequencing analysis.

Yuan H, Li X, Zhang X, Kang R, Tang D. CISD1 inhibits ferroptosis by protection against mitochondrial lipid peroxidation. *Biochem Biophys Res Commun* **478**:838-844 (2016). doi: 10.1016/j.bbrc.2016.08.034.

Yue X, Izcue A, Borggrefe T. Essential role of mediator subunit Med1 in invariant natural killer T-cell development. *Proc Natl Acad Sci USA* **108**:17105-17110 (2011). https://doi.org/10.1073/pnas.1109095108

You K, Wang L, Chou CH, Liu K, Nakata T, Jaiswal A, Yao J, Lefkovith A, Omar A, Perrigoue JG, Towne JE, Regev A, Graham DB, Xavier RJ. QRICH1 dictates the outcome of ER stress through transcriptional control of proteostasis. *Science* **371**:eabb6896 (2021). doi: 10.1126/science.abb6896.

Zhan Z, Cao H, Xie X, Yang L, Zhang P, Chen Y, Fan H, Liu Z, Liu X. Phosphatase PP4 Negatively Regulates Type I IFN Production and Antiviral Innate Immunity by Dephosphorylating and Deactivating TBK1. *J Immunol* **195**:3849-57 (2015). doi: 10.4049/jimmunol.1403083.

Zhang J, Stirling B, Temmerman ST, Ma CA, Fuss IJ, Derry JM, Jain A. Impaired regulation of NF-kappaB and increased susceptibility to colitis-associated tumorigenesis in CYLD-deficient mice. *J Clin Invest* **116**:3042-3049 (2006). doi: 10.1172/JCI28746.

Zhang Y, Dubé PE, Washington MK, Yan F, Polk DB. ERBB2 and ERBB3 regulate recovery from dextran sulfate sodium-induced colitis by promoting mouse colon epithelial cell survival. Lab Invest **92**:437-450 (2012). 10.1038/labinvest.2011.192

Zhang M, Wang L, Zhao X, Zhao K, Meng H, Zhao W, Gao C. TRAF-interacting protein (TRIP) negatively regulates IFN-β production and antiviral response by promoting proteasomal degradation of TANK-binding kinase 1. *J Exp Med* **209**:1703-1711 (2012). doi: 10.1084/jem.20120024.

Zhang Y, Liu RB, Cao Q, Fan KQ, Huang LJ, Yu JS, Gao ZJ, Huang T, Zhong JY, Mao XT, Wang F, Xiao P, Zhao Y, Feng XH, Li YY, Jin J. USP16-mediated deubiquitination of calcineurin A controls peripheral T cell maintenance. *J Clin Invest* **129**:2856-2871 (2019). doi: 10.1172/JCI123801.

Zhang C, Han X, Yang L, Fu J, Sun C, Huang S, Xiao W, Gao Y, Liang Q, Wang X, Luo F, Lu W, Zhou Y. Circular RNA *circPPM1F* modulates M1 macrophage activation and pancreatic islet inflammation in type 1 diabetes mellitus. *Theranostics* **10**:10908-10924 (2020). doi: 10.7150/thno.48264.

Zhao J, Wei J, Mialki RK, Mallampalli DF, Chen BB, Coon T, Zou C, Mallampalli RK, Zhao Y. F-box protein FBXL19-mediated ubiquitination and degradation of the receptor for IL-33 limits pulmonary inflammation. *Nat Immunol* **13**:651-658 (2012). doi: 10.1038/ni.2341.

Zhao R, Liu Y, Wang H, Yang J, Niu W, Fan S, Xiong W, Ma J, Li X, Phillips JB, Tan M, Qiu Y, Li G, Zhou M. BRD7 plays an anti-inflammatory role during early acute inflammation by inhibiting activation of the NF-κB signaling pathway. *Cell Mol Immunol* **14**:830-841 (2017). doi: 10.1038/cmi.2016.31.

Zhao L, Hao Y, Song Z, Fan Y, Li S. TRIM37 negatively regulates inflammatory responses induced by virus infection via controlling TRAF6 ubiquitination. *Biochem Biophys Res Commun* 556:87-92 (2021). https://doi.org/10.1016/j.bbrc.2021.03.147

Zhao X, Di Q, Yu J, Quan J, Xiao Y, Zhu H, Li H, Ling J, Chen W. USP19 (ubiquitin specific peptidase 19) promotes TBK1 (TANK-binding kinase 1) degradation via chaperone-mediated autophagy. *Autophagy* **18**:891-908 (2022). doi: 10.1080/15548627.2021.1963155.

Zhao JH, Stacey D, Eriksson N, Macdonald-Dunlop E, Hedman ÅK, Kalnapenkis A, Enroth S, Cozzetto D, Digby-Bell J, Marten J, Folkersen L, Herder C, Jonsson L, Bergen SE, Gieger C, Needham EJ, Surendran P; Estonian Biobank Research Team; Paul DS, Polasek O, Thorand B, Grallert H, Roden M, Võsa U, Esko T, Hayward C, Johansson Å, Gyllensten U, Powell N, Hansson O, Mattsson-Carlgren N, Joshi PK, Danesh J, Padyukov L, Klareskog L, Landén M, Wilson JF, Siegbahn A, Wallentin L, Mälarstig A, Butterworth AS, Peters JE. Author Correction: Genetics of circulating inflammatory proteins identifies drivers of immune-mediated disease risk and therapeutic targets. *Nat Immunol* **24**:1960 (2023). doi: 10.1038/s41590-023-01635-6.

Zheng H, Gupta V, Patterson-Fortin J, Bhattacharya S, Katlinski K, Wu J, Varghese B, Carbone CJ, Aressy B, Fuchs SY, Greenberg RA. A BRISC-SHMT complex deubiquitinates IFNAR1 and regulates interferon responses. *Cell Rep* **5**:180-193 (2023). doi: 10.1016/j.celrep.2013.08.025.

Zhou Z, He H, Wang K, Shi X, Wang Y, Su Y, Wang Y, Li D, Liu W, Zhang Y, Shen L, Han W, Shen L, Ding J, Shao F. Granzyme A from cytotoxic lymphocytes cleaves GSDMB to trigger pyroptosis in target cells. *Science* **368**:eaaz7548 (2020). doi: 10.1126/science.aaz7548.

Zhu L, Li Y, Zhou L, Yang G, Wang Y, Han J, Li L, Zhang S. Role of RING-Type E3 Ubiquitin Ligases in Inflammatory Signalling and Inflammatory Bowel Disease. *Mediators Inflamm* **2020**:5310180 (2020). doi: 10.1155/2020/5310180.

Zou D, Zhou S, Wang H, Gou J, Wang S. Knee Joint Swelling at Presentation: A Case of Pediatric Crohn Disease With a TNFAIP3 Mutation. *Pediatrics* **146**:e20193416 (2020). doi: 10.1542/peds.2019-3416.
